# Reconstructing Dengue Exposure Dynamics and Assessing the Public Health Impact of Vaccination Scenarios Using Mathematical Models in Brazil

**DOI:** 10.64898/2026.10.15.26365586

**Authors:** Youngsuk Ko, Cleber D. Santos, Angkana T. Huang, Henrik Salje, Derek A.T. Cummings, Albert I. Ko

**Author notes:** Corresponding authors: Youngsuk Ko; Albert I. Ko.

## Abstract

Brazil experienced its worst dengue epidemic in 2024, with an increase in severe outcomes among elderly individuals, highlighting the need for vaccination strategies informed by population immunity. We reconstructed state- and age-specific immune profiles by fitting an age-structured catalytic model to dengue surveillance data from 2000–2025. This inferred immune profile was incorporated into an SIR-type transmission model to project disease burden and to evaluate the public health impact of vaccine allocation strategies. We observed substantial spatial heterogeneity in accumulated immunity, which was closely associated with projected burden and optimal allocation. Strategies prioritizing states with lower historical incidence and larger susceptible populations yielded greater benefits. Under limited vaccine supply, targeting children resulted in the greatest reductions in DALYs, whereas targeting elderly populations achieved smaller DALY reductions but larger reductions in mortality. We further showed that accelerated vaccine supply markedly increased impact, while allocation choices remained critical when availability was constrained.

## Introduction

Dengue virus remains a critical global public health challenge and is of particular concern in Brazil. In 2024, Brazil experienced its worst epidemic on record, reporting over six million cases and 5,000 deaths ^1–3^. The true disease burden is likely higher due to underreporting ^2–4^. Older adults have accounted for an increasing share of severe and fatal dengue outcomes since 2000, with individuals aged ≥60 comprising more than half of dengue-related deaths in 2024 ^3,5^. This shift is thought to arise from comorbidities and weakened immune responses ^3,6–10^. However, age-related severity may reflect accumulated immunity, which differs across regions and age groups according to cumulative infection history. Uncertainty about the causes of this increase in older individuals and underlying age patterns of burden limits the ability to design targeted vaccination strategies.

As of 2026, two dengue vaccines are licensed globally: Dengvaxia® (CYD-TDV), approved for individuals aged 9–45 in endemic areas and recommended only for seropositive individuals due to risk of antibody-dependent enhancement in seronegative persons, whose production was discontinued due to declining demand, and Qdenga® (TAK-003), approved for individuals aged 4–60 regardless of serostatus ^11–16^. With dengue threatening an estimated 3.9 billion people worldwide, vaccine supply remains far below demand; TAK-003 production is projected to reach a maximum of 100 million doses annually by 2030, insufficient for even a single large country such as Brazil with a population exceeding 200 million ^17,18^. In 2023, WHO’s Strategic Advisory Group of Experts recommended prioritizing Qdenga vaccination for children aged 6–16 years in high-transmission settings ^12,13^. While this strategy effectively targets secondary infections in younger populations, it leaves elderly largely untargeted with intervention. Evidence for vaccine efficacy in elderly remains limited, although experts increasingly advocate for their inclusion in future vaccination programs ^19^.

A major development occurred in late 2025, when ANVISA, the Brazilian health regulatory agency, approved a third dengue vaccine, the Butantan Institute’s single-dose tetravalent vaccine, for individuals aged 12–59 ^20^. More than one million doses were produced, and manufacturing expansion was projected to scale output to tens of millions of annual doses by 2026. However, in June 2026 the national rollout was temporarily suspended as a precautionary measure following reports of severe adverse events, including deaths, among vaccine recipients, pending investigation of a possible causal link—a development that frames the supply scenarios examined in this study ^21^.

Mathematical models are essential for informing vaccination policy, enabling integration of seroprevalence data, age-specific risks, and vaccine efficacy profiles. Previous studies estimated dengue’s force of infection (FOI) and case reporting rates across multiple countries, revealing substantial spatial and demographic heterogeneities ^22–27^. Other models evaluated optimal vaccine deployment strategies, focusing on serostatus, age, and transmission intensity, and highlighted the importance of spatial variation in shaping optimal allocation ^28–34^. However, no study has comprehensively reconstructed Brazil’s regional and age-specific immunity profiles or systematically assessed vaccine allocation strategies that incorporate these heterogeneities. The public health value of vaccinating older adults also remains underexplored.

Here, we evaluated optimal dengue vaccine allocation strategies for Brazil. We reconstructed state- and age-specific immunity profiles using SINAN case notification data and a catalytic modeling framework, estimating population distributions by number of previous dengue infections. We integrated these reconstructed immune landscapes into an SIR transmission model with vaccination, simulating the public health impact of multiple vaccine supply and targeting strategies over the next five years. Motivated by the approval and potential large-scale production of the Butantan vaccine, we also evaluated an accelerated supply scenario representing a rapidly expanding national stockpile. **Fig. 1** provides an overview of our analytical workflow.

**Fig. 1:**
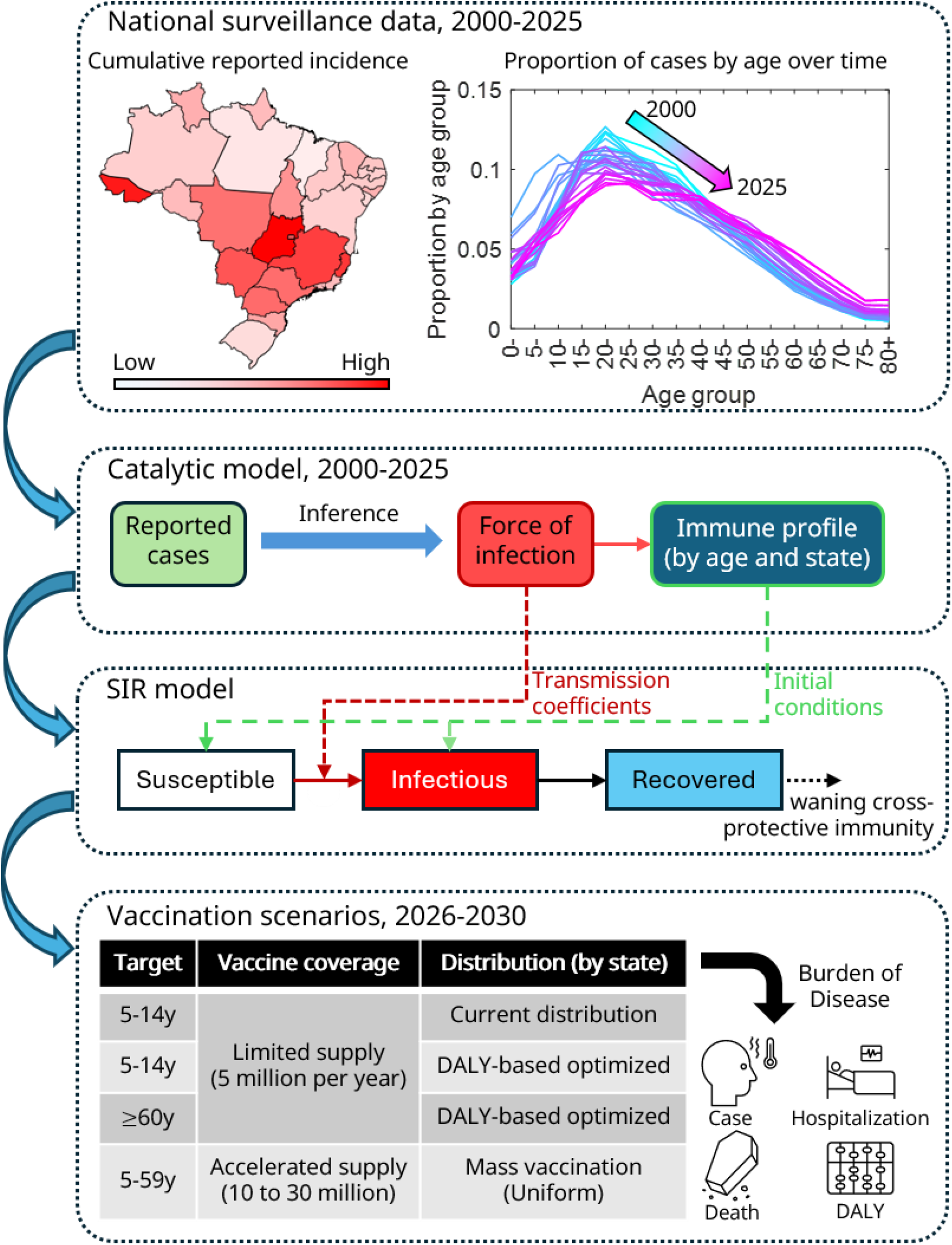
Conceptual workflow linking dengue surveillance, immunity inference, and vaccine allocation modeling. Reported case data from SINAN are used to estimate force of infection and derive age- and state-specific immune profiles. These inferred immune landscapes are then incorporated into a transmission model to assess the population-level health impact of alternative vaccine prioritization strategies, including limited and accelerated supply scenarios distributed by different targeting rules.

## Results

### Immune profile reconstruction using a catalytic modeling framework

Using catalytic models fit to case surveillance data from Brazilian national reporting system (SINAN), we estimated state-, age-, and time-specific FOI (**Fig. 2**). Panel A illustrates temporal variation in FOI using three states (Bahia, São Paulo, and Santa Catarina) as examples (full per-state, per-serotype FOI trajectories for all 27 states are shown in **Fig. S1**, with corresponding numerical estimates in **Table S2**). Panel B shows the age-specific FOI multipliers, which increased gradually from early childhood and peaked around ages 20–24y before steadily declining at older ages. Panels E and F display the reconstructed immune profiles by age for the years 2000 and 2026. In 2000, several northeastern states exhibited the highest prevalence of past infections, while many southern and southeastern states showed considerably lower immunity levels with large proportions of the population remaining naïve. Nationally, the population-weighted proportion of individuals with at least one prior DENV infection rose from 26.9% (95% CrI: 20.1–35.7) in 2000 to 83.3% (95% CrI: 78.8– 86.9) in 2025. By 2025, immunity levels had increased appreciably in nearly all states, with most regions showing higher proportions of individuals who had experienced multiple prior infections compared with 2000. Although southern states continued to have comparatively lower prevalence, they nonetheless experienced notable increases in accumulated immunity. Marked differences were also observed in the proportion of individuals with zero or one prior infection (dark curves in panels E and F), the group at risk of symptomatic disease upon subsequent exposure. The full temporal evolution of this proportion from 2000 to 2025 is shown in **Fig. S8**, and the corresponding state-level immune profile estimates for 2000 and 2026 are provided in **Table S7**. In 2026, this proportion varied substantially across states, ranging from approximately 28% in Goiás, a central-western state, to 68% in Rio Grande do Sul, a southern state. To assess the realism of our model-derived estimates, we compared the reconstructed immune profiles against age-stratified seroprevalence data from independent serosurveys conducted across multiple Brazilian states. The model-based estimates were broadly consistent with the observed serosurvey ranges: in 24 of 26 state-year comparisons, the serosurvey-reported values overlapped with the model’s 95% credible interval, providing external validation of the catalytic model outputs — noting that credible-interval overlap is a comparatively permissive validation criterion, and the two non-overlapping comparisons occurred in states and years with smaller serosurvey sample sizes and correspondingly wider empirical uncertainty (**Fig. S9**). The catalytic model also closely reproduced observed case-incidence trends across states (**Fig. S6**), and state-level case-to-infection ratios were consistent with expected patterns (**Fig. S7; Table S6**).

**Fig. 2:**
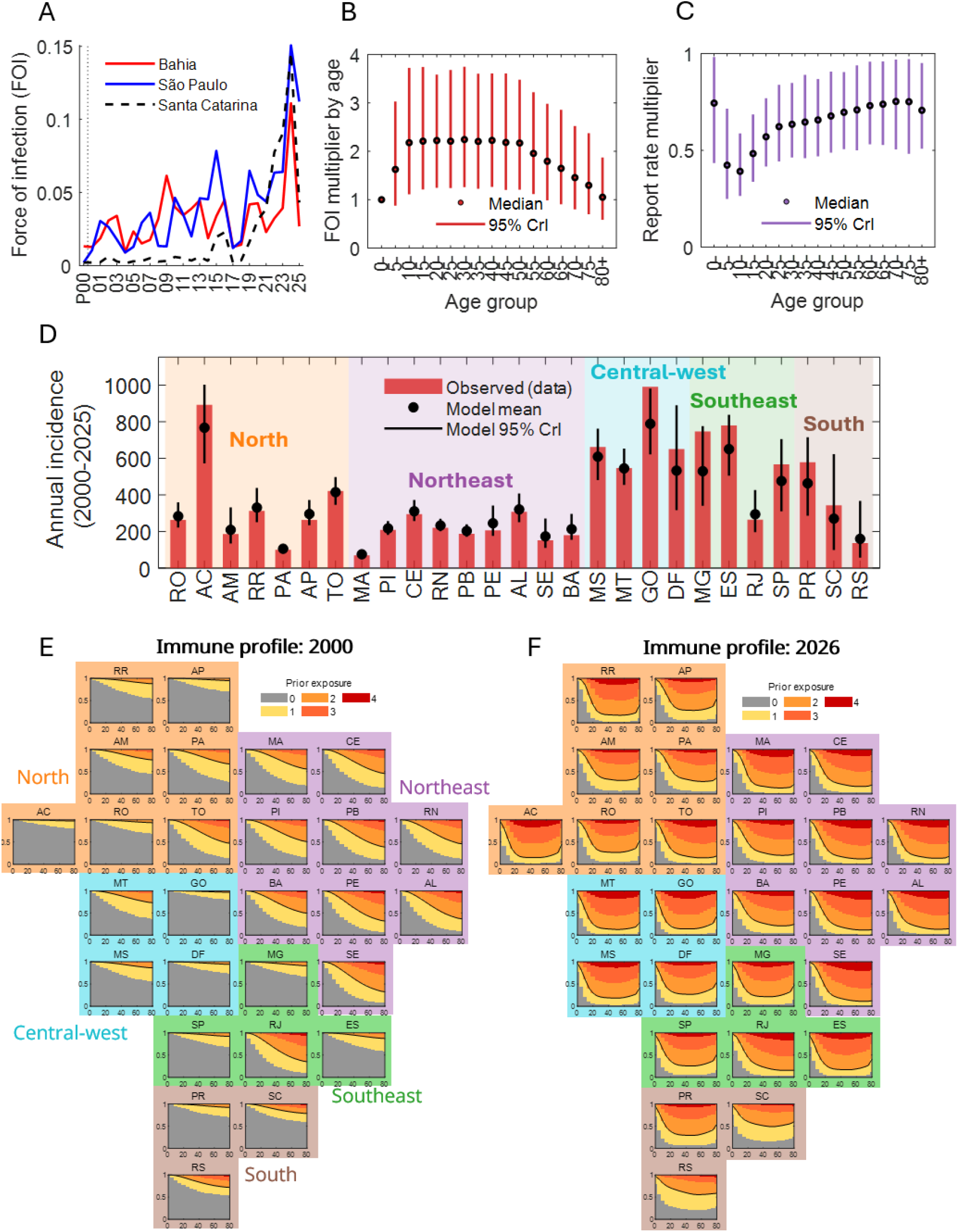
Parameter estimates, model fit, and reconstructed immune profiles from the catalytic modeling framework. (A) Temporal variation in the estimated median force of infection (FOI) illustrated for three example states: Bahia (Northeast), São Paulo (Southeast), and Santa Catarina (South). (B) Estimated age-specific FOI multipliers, shown as posterior medians (points) and 95% credible intervals (lines). (C) Estimated age-specific reporting rate multipliers, shown as posterior medians (points) and 95% credible intervals (lines). (D) Observed versus model-predicted average annual incidence (per 100,000) by state, 2000–2025; red bars show observed data, black points and lines show model means and 95% credible intervals; full state-level fit diagnostics are provided in the Supplementary Appendix. (E, F) Reconstructed age-specific immune profiles by state in 2000 (E) and 2026 (F). Colors indicate the proportion of the population with different numbers of prior dengue infections (from zero to four or more). The combined proportion of individuals with zero or one prior infection is highlighted by the black curve across panels. State panels are grouped and color-coded by geographic region.

### Projected Burden of Disease (BoD) in the absence of vaccination and associations with epidemiological indicators

The probability of symptomatic, reported dengue disease for a given age and infection history was derived by combining the age- and state-specific force of infection underlying the reconstructed immune profiles (**Fig. 2A, B, E, F**), the infection-order-dependent apparent illness ratio (**Fig. S4**), and the age-specific reporting-rate adjustment (**Fig. 2C**); overall model fit to observed incidence is shown in **Fig. 2D**. Using the reconstructed immune profiles, we projected the BoD over the next five years assuming no vaccination. In this study, BoD was assessed in terms of cases, hospitalizations, deaths, and Disability-Adjusted Life Years (DALYs). **Fig. 3A–D** shows state-level projections for annual cases, hospitalizations, deaths, and DALYs per 100,000 population. We observed that BoD tended to increase toward the southern states, with annual DALYs per 100,000 population ranging from roughly 150–230 in most northern states to over 300 in the southernmost states. Detailed BoD estimates for all states are provided in the **Supplementary Appendix** (**Table S12**). Corresponding age- and region-specific breakdowns of projected BoD are shown in **Fig. S12**.

**Fig. 3:**
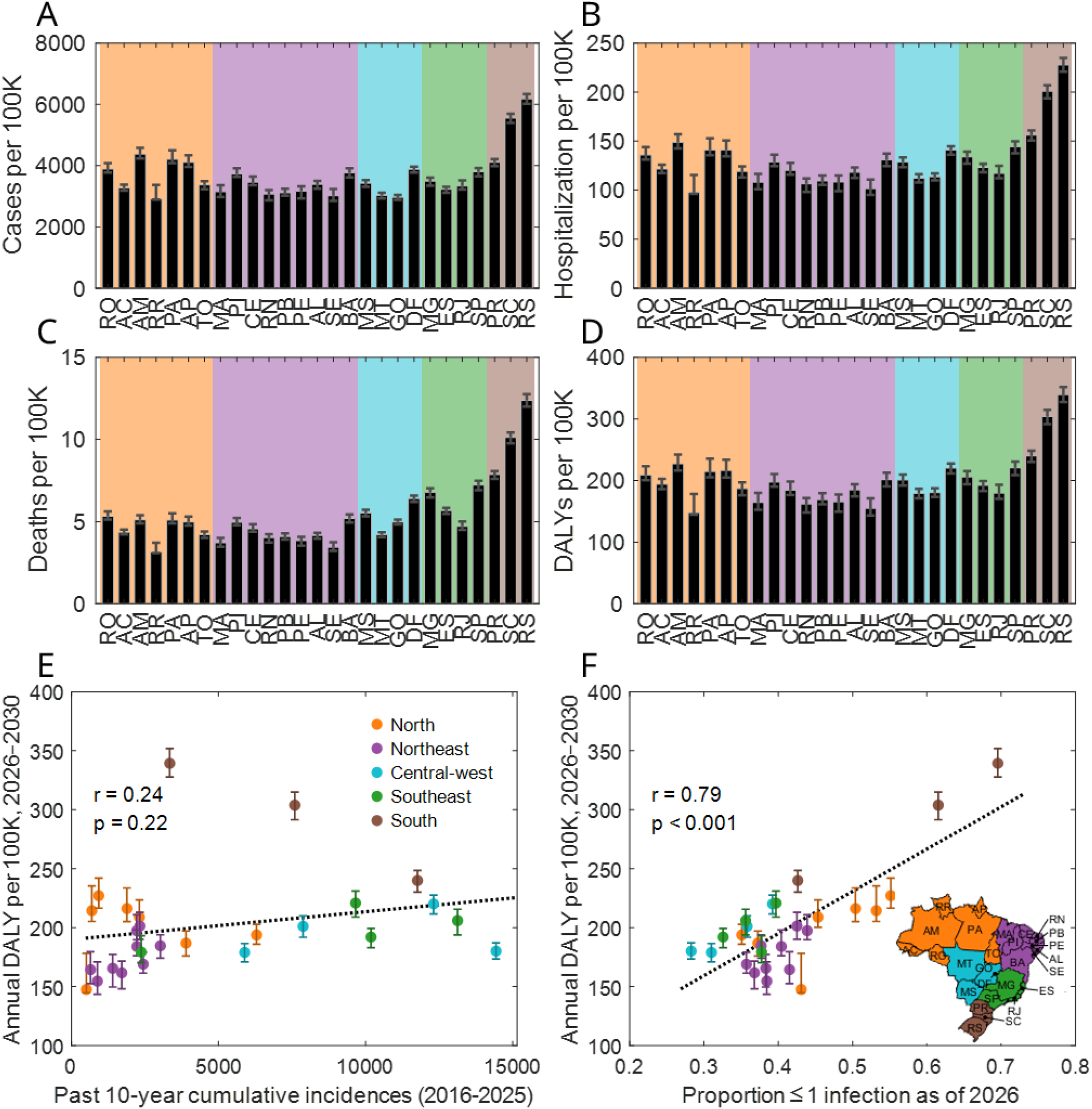
Projected annual burden of disease by state based on five-year simulations in the absence of vaccination. (A–D) State-level projections of cases (A), hospitalizations (B), deaths (C), and Disability-Adjusted Life Years (DALYs; D) per 100,000 population. Background colors indicate geographical regions, consistent with **Fig. 2**. (E, F). Associations between projected DALYs per 100,000 and two epidemiological indicators: cumulative dengue incidence over the past ten years, 2016 to 2025 (E) and the proportion of individuals with zero or one prior infection as of 2026 (F), corresponding to the area under the dark curves in **Fig. 2E–F**.

We next assessed which past epidemiological indicators were most strongly associated with future BoD. Correlations were evaluated for (i) cumulative dengue incidence over the past ten years (2016–2025), and (ii) the proportion of the population with zero or one prior infection as of 2026. Cumulative past incidence exhibited a weak, statistically non-significant association with future burden (r = 0.24; p = 0.22). These associations are descriptive and do not adjust for other state-level covariates, such as climate, urbanization, or socioeconomic factors, that may independently influence both past incidence and projected future burden. The stronger association was with the proportion of the population with zero or one prior infection (r = 0.79; p < 0.001). This association should be interpreted with caution: because future BoD is generated by a mechanistic model in which new cases arise predominantly among individuals with zero or one prior infection, the strong correlation with this indicator partly reflects the model’s structural design rather than an independent validation of it as an epidemiological predictor. Nonetheless, it remains a practically useful indicator for identifying states at greatest projected risk within this modeling framework.

### Projected public health benefits under vaccine allocation and supply scenarios

We first evaluated a baseline vaccination scenario reflecting Brazil’s current vaccine distribution pattern, in which vaccine supply is directed to children aged 5–14 years and distributed across states following the observed 2024–2025 allocation (**Table S10**), to provide a reference point for subsequent comparisons. Using this trend, we assumed five million individuals vaccinated annually and projected the resulting public health impact over a five-year horizon (**Fig. 4**). Panels A–D show BoD averted per 1,000 vaccinated individuals for cases, hospitalizations, deaths, and DALYs, stratified by age group.

**Fig. 4:**
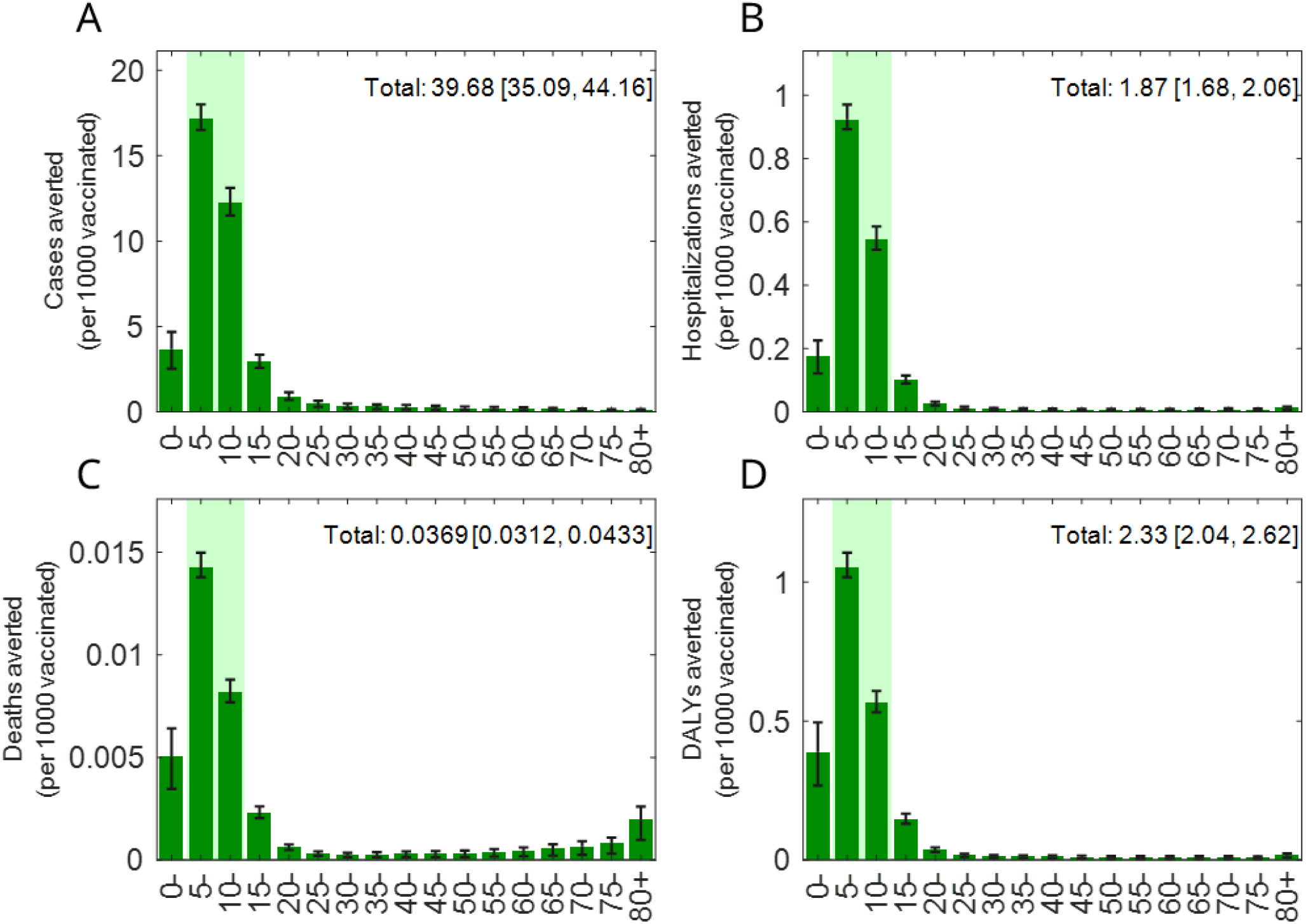
Estimated public health impact under the baseline vaccination scenario reflecting Brazil’s current distribution trend. Panels A through D present the burden of disease averted (cases, hospitalizations, deaths, and DALYs) expressed per 1,000 vaccinated individuals, stratified by age group as well as shown in total. The light green shading indicates the vaccine-targeted age group (5–14 years).

Marked reductions were observed within the target age group (5–14 years). Non-targeted age groups also benefited indirectly, even though the model includes no direct reduction in the infectiousness of vaccinated individuals who become infected. This benefit arose because vaccine-induced reductions in susceptibility to infection (0.02–0.48 depending on serostatus and time since vaccination; **Table S9**) lowered the number of infectious individuals and thus overall transmission. State-level results for this baseline scenario are provided in **Fig. S13**.

Building on the baseline vaccination results, we next compared multiple vaccine allocation strategies under limited and expanded supply conditions. **Table 1** summarizes the projected public health impacts across all vaccination scenarios considered in this study, including the baseline simulation reflecting Brazil’s current vaccine distribution pattern, optimized allocation strategies, and accelerated supply scenarios.

**Table 1:** Burden of disease averted under various vaccination scenarios. Projected reductions in cases, hospitalizations, deaths, and DALYs under different vaccination strategies and vaccine supply conditions. Values are reported as total numbers averted over the simulation period, with numbers in parentheses indicating outcomes averted per 1,000 vaccinated individuals.

| Vaccination target | Vaccine coverage per year | Vaccine allocation by state | Cases averted, total (per 1000 vaccinated) | Hospitalizations averted, total (per 1000 vaccinated) | Deaths averted, total (per 1000 vaccinated) | DALYs averted, total (per 1000 vaccinated) |
| --- | --- | --- | --- | --- | --- | --- |
| Children (5-14y) | 5m | Current distribution (baseline) | 992,073<br>[877,355, 1,103,948]<br>(39.68<br>[35.09, 44.16]) | 46,839<br>[41,946, 51,516]<br>(1.87<br>[1.68, 2.06]) | 922<br>[781, 1,082]<br>(0.04<br>[0.03, 0.04]) | 58,308<br>[51,116, 65,488]<br>(2.33<br>[2.04, 2.62]) |
| Children (5-14y) |  | DALY-based optimized | 1,144,569<br>[994,855, 1,255,178]<br>(45.78<br>[39.79, 50.21]) | 53,437<br>[47,526, 58,009]<br>(2.14<br>[1.90, 2.32]) | 1,051<br>[874, 1,208]<br>(0.04<br>[0.03, 0.05]) | 66,962<br>[57,902, 73,920]<br>(2.68<br>[2.32, 2.96]) |
| Elderly (≥60y) |  | DALY-based optimized | 409,062<br>[391,607, 463,957]<br>(16.36<br>[15.66, 18.56]) | 30,544<br>[29,516, 33,674]<br>(1.22<br>[1.18, 1.35]) | 3,001<br>[2,936, 3,227]<br>(0.12<br>[0.12, 0.13]) | 40,776<br>[39,202, 45,270]<br>(1.63<br>[1.57, 1.81]) |
| Mass vaccination (5-59y) | 10m | Uniform* | 1,063,593<br>[908,370, 1,202,443]<br>(21.27<br>[18.17, 24.05]) | 41,809<br>[35,659, 47,402]<br>(0.84<br>[0.71, 0.95]) | 1,220<br>[1,018, 1,427]<br>(0.02<br>[0.02, 0.03]) | 59,746<br>[50,136, 68,493]<br>(1.19<br>[1.00, 1.37]) |
|  | 20m |  | 2,129,416<br>[1,845,471, 2,432,254]<br>(21.29<br>[18.45, 24.32]) | 83,890<br>[72,646, 95,661]<br>(0.84<br>[0.73, 0.96]) | 2,448<br>[2,072, 2,876]<br>(0.02<br>[0.02, 0.03]) | 119,866<br>[102,282, 138,325]<br>(1.20<br>[1.02, 1.38]) |
|  | 30m |  | 3,234,549<br>[2,856,042, 3,663,980]<br>(21.56<br>[19.04, 24.43]) | 127,899<br>[112,716, 144,163]<br>(0.85<br>[0.75, 0.96]) | 3,734<br>[3,210, 4,328]<br>(0.02<br>[0.02, 0.03]) | 182,943<br>[159,345, 208,443]<br>(1.22<br>[1.06, 1.39]) |

\* Same coverage applied relative to the target population in each state.

Under a DALY-minimizing objective, the optimized strategy prioritizing children aged 5 to 14 years reduced the overall disease burden, averting 2.68 (range [2.32, 2.96]) DALYs per 1,000 vaccinated individuals. This corresponded to a 15% (range [13, 15]) improvement relative to the baseline scenario. In contrast, the optimized strategy targeting elderly individuals aged 60 years and above reduced DALYs less than the children-targeting baseline scenario, averting 1.63 (range [1.57, 1.81]) DALYs per 1,000 vaccinated individuals. This represented a 30% (range [23%, 31%]) decrease relative to the baseline scenario. However, when fatalities were considered, this elderly-targeting scenario substantially outperformed children-targeting scenarios, averting 0.12 (range [0.12, 0.13]) deaths per 1,000 vaccinated individuals. This corresponded to a 225% (range [198%, 276%]) improvement relative to the baseline scenario. Notably, even with limited supply, the elderly-targeting scenario was projected to avert more deaths (3,001 [2,936, 3,227]) than the 20-million-dose mass vaccination scenario targeting individuals aged 5–59 years (2,448 [2,072, 2,876]).

To examine intermediate allocation strategies under limited vaccine supply, we constructed linear combinations of the optimized allocations targeting children and elderly individuals, varying the relative contribution of each strategy. **Fig. 5** shows how BoD averting ratios change as the allocation shifts between children-targeting and elderly-targeting strategies. The response to mixed allocation strategies exhibited distinct regional heterogeneity. In the north, northeast, and central-west regions, shifting the allocation toward the elderly-targeting strategy was associated with monotonic declines in BoD averting ratios across cases, hospitalizations, fatalities, and DALYs. In contrast, the southeast and south regions showed distinct patterns, with greater emphasis on elderly-targeting consistently improving fatality reduction. In both regions, DALY-averting ratios also increased as the allocation shifted toward the elderly-targeting strategy, whereas case-averting ratios showed a non-monotonic response, with modest increases under elderly-dominant allocation (approximately 80% of vaccines allocated to the elderly), followed by declines at higher levels. At the nationwide level, when the share of vaccines allocated to children decreased from 100% to 60%, the DALY-averting ratio declined only slightly, from 2.89% (range [2.37%, 3.36%]) to 2.57% (range [2.20%, 2.97%]). In contrast, over the same range, the fatality-averting ratio increased substantially, from 1.54% (range [1.23%, 1.85%]) to 2.89% (range [2.64%, 3.34%]).

**Fig. 5:**
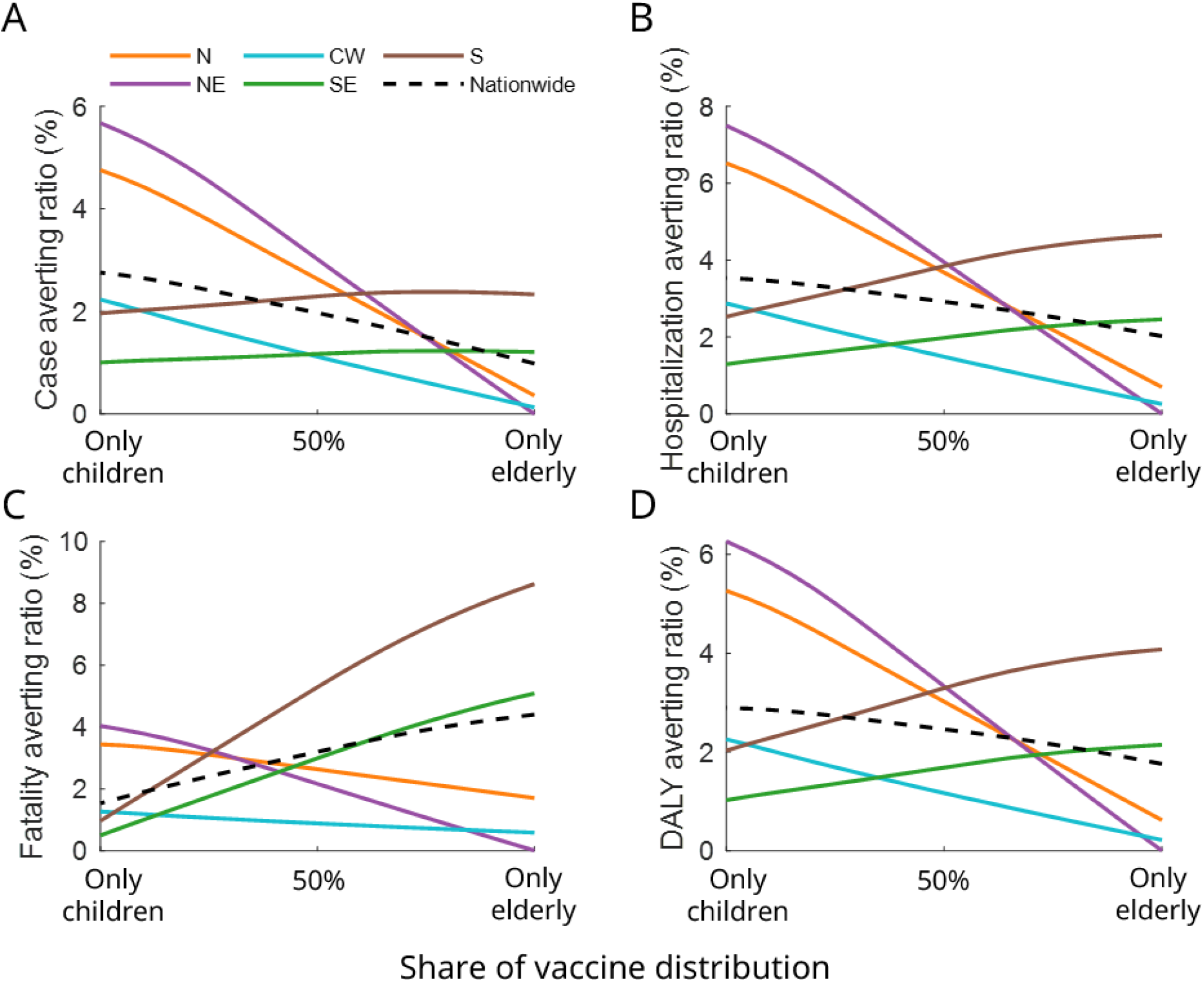
Burden-of-disease averting ratios under mixed children- and elderly-targeting vaccination strategies. Each ratio is the number of cases, hospitalizations, fatalities, or DALYs averted by vaccination over the five-year simulation period, divided by the corresponding number projected in the absence of vaccination (**Fig. 3**) for the same region or for Brazil as a whole. Results are shown as a function of the proportion of vaccine supply allocated to children or elderly individuals, obtained by linearly combining the optimized allocation strategies targeting children and elderly individuals. (A–D) Panels show averting ratios for cases (A), hospitalizations (B), fatalities (C), and DALYs (D). Curves are colored by geographical region (N: north, NE: northeast, CW: central-west, SE: southeast, S: south), and the dark dashed curve indicates nationwide scale.

In contrast to the limited-supply setting, expanding vaccine availability resulted in markedly larger absolute reductions in all BoD metrics (**Table 1**). Under the mass vaccination scenario targeting individuals aged 5–59 years, increasing annual vaccination capacity from 10 to 30 million individuals increased the total number of averted cases from approximately 1.06 million (range [0.91, 1.20 million]) to 3.23 million (range [2.86, 3.66 million]) and the total number of averted DALYs from about 59,700 (range [50,100, 68,500]) to 182,900 (range [159,300, 208,400]). Over the same range, the number of averted fatalities increased from approximately 1,220 (range [1,018, 1,427]) to 3,730 (range [3,210, 4,328]). Note that under the 30-million-dose scenario, vaccination coverage could reach approximately 90% of individuals aged 5–59 years during the simulation period (5 years).

## Discussion

In this study, we first applied a catalytic model to reconstruct age- and state-specific immune profiles from surveillance data and then incorporated these reconstructed immune landscapes into an SIR-type transmission model to project future BoD and quantify impacts relative to a predefined baseline, across multiple vaccine allocation scenarios and vaccine supply conditions. Notably, the unprecedented 2024 epidemic substantially elevated immunity levels—particularly in southern states previously characterized by lower seroprevalence—and this shift is captured in the 2026 initial immune profiles used as the starting point for our vaccination simulations.

Although dengue surveillance data in Brazil are based on passive reporting systems and are therefore subject to substantial underreporting and regional heterogeneity, our approach leverages a nationwide, simultaneous parameter estimation framework, which helped mitigate these challenges ^4,35^. By leveraging age-specific patterns in reported incidence across states, the catalytic model was able to jointly estimate state-specific FOI, reporting ratios (summarized in the **Supplementary Appendix**), and the average pre-2000 FOI corresponding to the period prior to systematic case aggregation (**Fig. S2**).

The reconstructed immune profile in 2000 (**Fig. 2E**) revealed markedly higher seroprevalence in the north and northeast regions, reflecting the earlier establishment of dengue transmission in those areas, predating systematic national surveillance ^36–40^. Over subsequent decades, continued circulation has expanded population exposure and led to the high nationwide seroprevalence as of 2026 (**Fig. 2F**). However, the proportion of individuals with zero or one prior dengue infection remains non-negligible in certain regions, and this susceptible or partially immune population is especially pronounced in southern states, where historical dengue incidence was lower. This pattern may be interpreted as an indirect signal of heightened vulnerability in southern states and underscores the need for increased attention to these areas in vaccine allocation and broader public health resource planning.

The SIR-type model made the implications of the reconstructed immune profiles for future BoD more explicit. When we simulated the system without vaccination, projected BoD (**Fig. 3**) tended to be larger in the southern states, consistent with the relative lack of naturally accumulated immunity highlighted above. The baseline scenario, implemented using Brazil’s vaccine allocation trends, showed a reduction in BoD (**Fig. 4**). In addition, consistent with the catalytic model results, the age groups considered for vaccination in Brazil are those with relatively lower cumulative infection exposure than older adult age groups (**Fig. 2F**), yet comparatively higher infection risk (**Fig. 2B**). Taken together, these findings support the rationale for Brazil’s children-targeting vaccination approach. Ferguson et al. highlighted that the population-level impact of dengue vaccination is shaped primarily by the immune composition of the targeted age groups and baseline seroprevalence, rather than age alone, based on analyses of a first-generation dengue vaccine with serostatus-dependent effects ^28^. More recently, Daniels et al. integrated trial-informed vaccine protection profiles into transmission modeling across settings defined by seroprevalence and showed that projected public health impact depends strongly on preexisting immunity in the population ^34^. Lastly, Shen et al. applied transmission models informed by age-specific exposure patterns in Thailand and showed that prioritizing younger age groups with lower cumulative exposure, but higher future infection risk can yield greater long-term reductions in BoD ^33^. Collectively, these studies reinforce the central role of heterogeneity in accumulated immunity in shaping vaccination impact, consistent with the patterns observed in Brazil. Optimized allocation vectors derived under the Shen and Daniels VE assumptions were highly correlated across most scenarios **(Fig. S18**), supporting the robustness of our qualitative conclusions to the choice of VE source.

In addition to the baseline scenario using real-world vaccine distribution trend, we evaluated optimized allocation strategies that minimize DALYs, considering two age group-targeting options: vaccination targeted to children aged 5 to 14 years and vaccination targeted to individuals aged 60 years and older. Across these analyses, underlying immune structure appears to play a central role in shaping future BoD and vaccine allocation outcomes, as evidenced by the negative association between allocation weight and cumulative incidence under the children-targeting optimized strategy (Supplementary **Figs. S14 and S15**). When allocation was restricted to elderly-targeting vaccination, the optimal strategy consistently placed greater weight on southern states (under both VE assumptions; **Figs. S16** and **S17**). This pattern suggests that incorporating elderly populations as vaccination targets favors prioritization of southern states, where dengue transmission has emerged more recently and accumulated immunity among older age groups remains relatively limited.

When combining the two optimized allocation strategies (**Fig. 5**), the mixed allocation analysis highlighted clear differences in how vaccination priorities affect BoD. From a nationwide perspective, vaccine allocation targeting children achieved the largest reductions in overall DALYs, whereas reallocating doses toward older adults yielded greater reduction in fatalities. This divergence arises because DALYs integrate both morbidity and mortality across the full lifespan, favoring strategies that prevent infections and severe disease in younger individuals who have more remaining life-years. In contrast, elderly-targeting allocation has a smaller effect on lifetime disease burden but a stronger impact on dengue-related death. Under limited vaccine supply, these objectives cannot be achieved simultaneously, and policymakers must decide whether to prioritize minimizing overall disease burden or averting the largest number of deaths. By reporting DALY- and mortality-based outcomes separately rather than collapsing them into a single composite score, our analysis makes explicit how the choice of metric embeds a normative weighting: DALYs privilege life-years and therefore favor younger recipients, whereas raw mortality counts weight each death equally regardless of age. This study focuses on epidemiological impact and implicitly assumes equal cost per dose delivered across allocation strategies; it does not incorporate differences in delivery cost, cold-chain requirements, or coverage logistics between school-based child vaccination and outreach to older adults, which would need to be integrated in a subsequent cost-effectiveness analysis to fully inform resource allocation decisions. Our results also depend on the waning of vaccine-induced protection, which was taken from published estimates and remains uncertain ^33,34^.

The regional contrasts under mixed strategies (**Fig. 5**) reflect differences in age-specific immunity. Southern regions have lower accumulated immunity even among older adults, who have experienced fewer past infections. Strategies prioritizing vaccination of elderly individuals are therefore likely to yield the greatest mortality benefits when focused on southern states, where immunological susceptibility remains high despite lower historical incidence.

Under accelerated supply scenarios, expanding vaccination capacity led to large absolute reductions in cases, hospitalizations, fatalities, and DALYs, even when vaccination was restricted to individuals aged 5 to 59 years. This finding highlights the dominant role of vaccine availability in determining population-level impact and underscores the potential public health value of large-scale vaccine production efforts, such as those anticipated for the Butantan single-dose vaccine. From a policy perspective, these results suggest that investments in expanding supply may yield greater returns than fine-tuning allocation strategies once coverage reaches sufficiently high levels.

The policy relevance of allocation under constrained supply has been underscored by events that unfolded as this work was completed. Butantan-DV, the first single-dose dengue vaccine, was approved in late 2025 and was expected to enable rapid, large-scale immunization—motivating the accelerated-supply scenarios examined here. In June 2026, however, Brazil’s Ministry of Health precautionarily suspended the Butantan-DV rollout pending investigation of rare serious adverse events identified through routine pharmacovigilance, for which a causal association with the vaccine has not been established ^21^. Irrespective of the outcome of that review, the immediate consequence is that Brazil’s near-term vaccine supply remains constrained and dependent on the two-dose Qdenga product with limited age eligibility. Our results speak directly to this setting: when supply is limited, allocation is consequential, and the objective chosen—minimizing overall disease burden versus averting deaths—materially alters which ages and regions should be prioritized. Because the framework is modular, it can be updated as the safety review concludes and as product-specific efficacy and eligibility are clarified, offering a transparent basis for revising allocation as the supply landscape evolves.

Several limitations should be acknowledged. Our catalytic model does not distinguish between dengue serotypes, despite the fact that dengue transmission in Brazil re-emerged gradually from the 1980s onward with different serotypes introduced at different times across regions, rather than through simultaneous circulation of all four serotypes nationwide. Accordingly, immunity is represented by the number of prior infections rather than by serotype-specific infection histories. The model also omits antibody-dependent enhancement (ADE) as an explicit serotype-resolved mechanism, although differential severity by infection order is captured empirically through the infection-order-dependent apparent illness ratios (**Table S3**). While ADE is epidemiologically important, its explicit incorporation requires serotype-resolved infection tracking, which is beyond the scope of the present framework; the empirically derived serostatus-dependent vaccine effectiveness parameters used in the SIR model partially capture the net immunological consequences of prior infection history. Notably, the infection-order-dependent apparent illness ratios estimated by the catalytic model (higher for secondary than primary infection, declining thereafter; see **Table S3**) already reproduce the population-level phenomenology commonly attributed to antibody-dependent enhancement following natural infection, even without explicit serotype tracking. For vaccine-induced immunity, TAK-003 was licensed irrespective of serostatus because clinical trials did not detect the enhancement signal previously observed for a first-generation dengue vaccine restricted to seropositive recipients; nonetheless, trial follow-up periods are finite, and any longer-term or age-specific enhancement risk outside the observed trial window would not be captured here. Similarly, vaccine effectiveness was modeled as depending only on prior infection status (seronegative or seropositive) and waning over time, and did not incorporate potential differences in effectiveness across dengue serotypes. Behavioral responses to vaccination, including hesitancy and logistical barriers, were also not modeled. Because most scenarios considered here assume vaccine supply far below full population coverage, differential uptake due to hesitancy would primarily rescale the effective number vaccinated rather than alter the relative ranking of allocation strategies or geographic priorities, which are the primary policy-relevant outputs of this study. Additionally, vaccine effectiveness was assumed to be age-independent; however, immunosenescence may reduce vaccine-induced protection in older adults, potentially leading to an overestimation of the mortality benefits of elderly-targeting strategies. Future work could extend this framework to incorporate serotype-specific transmission dynamics, ADE-mediated severity, and serotype-dependent vaccine effectiveness. The conversion of catalytic model-derived FOI estimates to SIR transmission parameters relied on an endemic equilibrium assumption, which may not fully capture the episodic, epidemic-driven nature of dengue transmission in Brazil, particularly in light of the unprecedented 2024 outbreak. This endemic-equilibrium approximation was adopted deliberately: directly fitting a fully time-varying, non-equilibrium transmission model across 27 states and four serotypes would introduce severe parameter identifiability problems given currently available surveillance data, whereas any alternative conversion from surveillance-derived force of infection to a transmission rate must still draw on the same long-run incidence trends already embedded in our catalytic model estimates. Similarly, the pre-2000 constant-FOI assumption was applied retrospectively across each individual’s full lifetime for tractability, without bounding it to a specific historical onset year (e.g., Brazil’s dengue re-emergence in the early 1980s), and may therefore overstate cumulative exposure among the oldest cohorts.

Beyond Brazil, these findings may also be informative for settings where dengue transmission has not yet become widespread or where future expansion of dengue risk is anticipated. In such contexts, reconstructing or projecting population immune profiles could motivate further investigation into vaccination design ahead of large-scale outbreaks, though such extrapolation beyond the Brazilian context studied here remains speculative. For example, in countries that have not experienced sustained dengue transmission over long periods, strategies emphasizing more evenly distributed vaccination across age groups, or prioritizing older adults rather than focusing exclusively on younger populations, may warrant consideration depending on underlying immunity patterns. These findings do not contradict WHO’s recommendation of prioritizing children in high-transmission settings, which remains effective for reducing overall dengue burden. Rather, they suggest that when the policy objective explicitly includes mortality reduction—particularly in settings where older adults bear a disproportionate share of fatal outcomes—complementary elderly-targeting strategies warrant consideration, especially in regions with lower historical exposure.

## Methods

We employed two mathematical modeling frameworks in this study: a catalytic model and an SIR-type model. A schematic overview of the modeling workflow, detailed parameter tables, and additional simulation results beyond those presented in the main text are provided in **the Supplementary Appendix**.

## Data sources

Dengue is a disease of mandatory notification in Brazil. All suspected cases must be reported to the Epidemiological Surveillance Service of the Municipal Health Department, which investigates each notification and assigns a final case classification (e.g., confirmed by laboratory criteria, confirmed by clinical-epidemiological criteria, or discarded) according to the Brazilian Ministry of Health case definitions. These notifications are consolidated in the National Notifiable Diseases Information System (SINAN, *Sistema de Informação de Agravos de Notificação*), which was the primary data source for case counts used in this study ^41^. Cases classified as confirmed by laboratory criteria or by clinical-epidemiological criteria were included in this analysis; cases classified as discarded were excluded. Life expectancy was obtained from the Brazilian Institute of Geography and Statistics (IBGE, *Instituto Brasileiro de Geografia e Estatística*) ^42^.

### Assumptions for catalytic model framework

We developed a Bayesian age-structured catalytic model to estimate the cumulative DENV infection history across 27 Brazilian states from 2000 to 2025. The model accounts for sequential infection by up to four distinct DENV serotypes, incorporates age- and state-specific heterogeneity in transmission intensity and case reporting, and infers annual force of infection (FOI) jointly across all states. The following assumptions underlie the model framework.

1. **Serotype co-circulation**: All four DENV serotypes (DENV-1 through DENV-4) are assumed to have co-circulated throughout the entire study period. This simplifying assumption removes temporal complexity associated with the historical introduction of individual serotypes and allows the model to focus on overall transmission intensity, acknowledging that the sequential introduction of serotypes in Brazil has been documented since the 1980s ^36–38,43–47^.
2. **Homogeneous mixing and per-serotype FOI**: Individuals within the same age group and state are assumed to mix homogeneously and share equal exposure to infection risk. Reinfection with the same serotype is not permitted, and lifelong immunity is assumed upon infection. The model does not track serotype identities explicitly but instead follows the cumulative number of distinct infections per individual. No temporary cross-protection or antibody-dependent enhancement is modeled; all infections are assumed to occur independently, conditional on previous exposure. For example, let the per-serotype FOI be λ; an individual with 2 prior infections has a total infection hazard of (4 ™ 2) × λ = 2λ, corresponding to the number of serotypes they have not yet encountered.
3. **FOI before and after year 2000**: Due to data limitations and for simplicity, the per-serotype FOI prior to the year 2000 is set as a state-specific constant, reflecting the average transmission intensity, applied retrospectively from each individual’s birth year through 1999 for tractability; this constant-FOI assumption is not bounded to a specific historical onset year (e.g., Brazil’s dengue re-emergence in the early 1980s), and may therefore overstate cumulative exposure among the oldest cohorts. From 2000 onward, the per-serotype FOI is assumed to be piecewise constant over time, defined within discrete, pre-specified intervals (i.e., annually). Within each interval, FOI may vary by age group, time, and state, but remains constant throughout the time step. The age-dependent weights (multiplier) for FOI are assumed to be consistent nationwide.
4. **Apparent illness and case report ratio**: Similar to the structure of FOI, these parameters vary by age group, time, and state, and the age-dependent weights are assumed to be consistent across the country. In addition, the apparent illness ratio differs by prior infection status, assumed to be highest for secondary infections, followed by primary, tertiary, and quaternary infections ^48–50^.

## Model notations

From the year 2000 onward, the population is grouped into 5-year age intervals (0–4, 5–9, …, and 80+ years; 17 groups total, indexed *a*). For the purpose of accumulating lifetime FOI exposure, the model additionally tracks single-year age cohorts *a*^∗^= 1, …, 81 (representing ages 0 through 80+), each mapped to its corresponding 5-year group (*a*). The following variables and parameters are used.

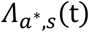: Cumulative per-serotype force of infection experienced by an individual of single-year age *a*^∗^in state *s*, up to time *t* . Prior to year 2000, this is accumulated linearly across age under the state-specific constant FOI: 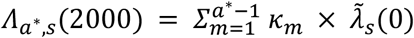. From 2000 onward, each cohort ages forward one year at a time, accumulating that year’s FOI: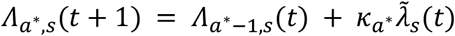

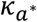: Age-specific FOI multiplier at single-year age *a*^∗^, mapped from the corresponding 5-year group *a*; assumed consistent nationwide.

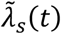: Time-dependent per-serotype FOI by state, with 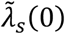denoting the state-specific constant value applied prior to year 2000.

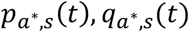: The probability that an individual of age *a*^∗^in state s has been infected by a given single serotype by time *t*, and its complement, obtained from the cumulative per-serotype force of infection:

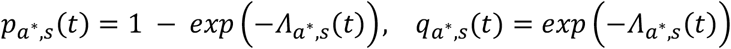

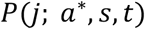: The probability that an individual of age *a*^∗^in state *s* has accumulated exactly *j* prior infections (*j* = 0, 1, 2, 3) by time t, obtained from the binomial expansion across the four independent serotypes:

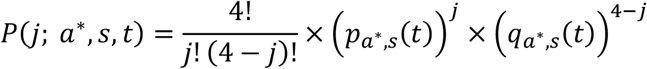

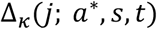: The within-year hazard of acquiring at least one new infection among the (4− *j*) serotypes not yet encountered, for an individual with j prior infections: 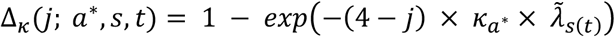

*δ*_*k*_: The apparent illness ratio associated with the k-th infection order (*k* = 1: primary, *k* = 2: secondary, *k* = 3: tertiary, *k* = 4: quaternary), parameterized as: *δ*_1_, *δ*_2_, *δ*_3_ = *δ*_1_ × *c*_*tert*_, *δ*_4_ = *δ*_3_ × *c*_*quat*_ where *δ*_1_ and *δ*_2_ are estimated directly, and *c*_*tert*_, *c*_*quat*_ are fractional multipliers governing the apparent illness ratio of tertiary and quaternary infections relative to the preceding infection order.

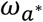: Age group-specific multiplier on reporting probability at single-year age *a*^∗^, mapped from the corresponding 5-year group *a*; assumed consistent nationwide.

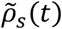 : State- and time-specific reporting rate, capturing spatiotemporal heterogeneity in surveillance and case ascertainment.

## Model formulation

The expected number of newly apparent (symptomatic, reported) infections at single-year age *a*^∗^in state *s* at time *t* is obtained by summing, over all possible prior infection counts *j* = 0, 1, 2, 3, the probability of having exactly *j* prior infections, weighted by the probability of newly acquiring at least one additional infection within the current year and the corresponding apparent illness ratio for that infection order:

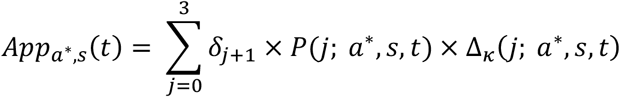

This single-year quantity is averaged across the five single-year ages comprising each 5-year age group *a* to obtain the age group-level apparent infection probability, *App*_*a,s*_(*t*). The expected number of reported cases (*µ*_*a,s*_(*t*)) is then obtained by scaling this probability by the age-, state-, and time-specific reporting components and the observed population size:

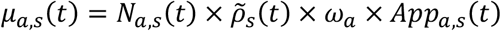

where *N*_*a,s*_(*t*)is the observed population size for age group *a*, state *s*, and year *t*.

## Likelihood and case weighting

Let *Y*_*a,s*_(*t*)denote the observed number of reported SINAN dengue cases for age group *a*, state *s*, and year *t*. Cases were modeled using a negative binomial likelihood to account for overdispersion in surveillance data:

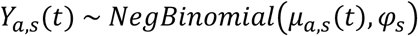

where φ_*s*_ is a state-specific overdispersion parameter shared across age groups within each state. Since the model already captures temporal variation through the annual state-specific FOI and reporting rate, freely estimating φ_*s*_ would result in upward bias driven by residual variance already accounted for by these temporal parameters, potentially inducing overfitting in other model parameters. To avoid this, φ_*s*_was fixed at values derived empirically from the observed data using the method of moments:

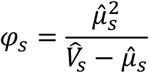

where 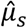 and 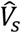 are the empirical mean and variance of annual total case counts in each state *s* across 2000–2025. To prevent large states with disproportionately high case counts from dominating the joint likelihood across all 27 states, the log-likelihood contribution of each state was weighted by the inverse square root of its total observed case count. This weighting scheme is motivated by variance stabilization under the negative binomial model: since case count variance scales with the mean, states with larger case volumes have proportionally larger absolute variability, and down-weighting equalizes the effective contribution of each state to the posterior.

## Parameter estimation

Parameters were estimated using Hamiltonian Monte Carlo (HMC) via Stan (cmdstanr interface in R), with computation parallelized across states using the reduce_sum construct ^51^. Four independent chains were run with 500 warm-up and 500 sampling iterations each, yielding 2,000 posterior draws in total, with a maximum treedepth of 12. Convergence was assessed using R-hat statistics, bulk and tail effective sample sizes (ESS), the number of divergent transitions, and the E-BFMI. All diagnostics indicated satisfactory convergence; detailed results are provided in the **Supplementary Appendix**.

Age-specific FOI multiplier. The multiplier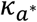(mapped to 17 five-year age groups) was constrained to be non-negative but was not assigned an explicit prior distribution, allowing its age profile to be fully determined by the data. Posterior estimates of this age-specific multiplier are provided in **Table S1**.

Force of infection. The annual per-serotype FOI 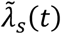was assigned an *Exponential*(*(4/mean*(*k*_*a*_))prior, while the pre-2000 constant FOI 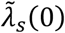was assigned a tighter *Exponential*(*(4/mean*(*κ*_*a*_))prior, where *mean*(*k*_*a*_)is the mean of the estimated age-specific multiplier across all age groups. Because *κ*_*a*_ was left unconstrained in scale, this formulation ties the prior mean of the baseline FOI inversely to the average magnitude of the age multiplier, preventing the product 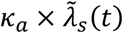 — which determines the effective per-serotype FOI — from drifting to an implausible joint scale, without imposing an explicit normalization constraint on *κ*_*a*_ that could otherwise distort its estimated age profile.

Age-specific reporting multiplier. The multiplier 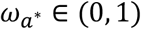 was assigned a first-order random walk prior on the truncated normal scale for age groups *a* = 2, …, 17: ω_*a*_ | ω_*a*−1_ ∼ *TruncatedNormal*(ω_*a*−1_, 1, 0, 1) with the prior log-likelihood scaled by the number of states *n*_*s*_ = 27 to reflect the pooled evidence across states for this nationally shared parameter. The first age group, ω_1_ (ages 0–4), was instead assigned a *Beta*(1.*5*,1)prior, also scaled by *n*_*s*_, reflecting the epidemiological expectation that infants and young children tend to have relatively higher healthcare-seeking and reporting rates. The random walk prior to subsequent age groups penalizes large abrupt changes between adjacent groups while remaining agnostic about the overall direction of the age profile beyond the first group.

Apparent illness ratios. Prior distributions for *δ*_1_ and *δ*_2_ were specified as *Beta*(4,16)(mean 0.20) and *Beta*(5, 5)(mean 0.50), respectively, consistent with the expectation that secondary infections are associated with a higher apparent illness ratio than primary infections. Posterior estimates of infection-order-dependent apparent illness ratios are shown in **Fig. S4** and **Table S3**. Posterior estimates of age-specific reporting-rate adjustment multipliers are shown in **Fig. 2C** and **Table S4**. The fractional multipliers *c*_*tert*_ and *c*_*quat*_ were not assigned explicit priors beyond their [0, 1] bound. Prior log-likelihoods for *δ*_1_ and *δ*_2_ were scaled by the number of states *n*_*s*_ = 27, reflecting the pooled evidence across all states for these nationally shared parameters. State- and time-specific reporting rate. 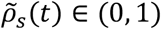was assigned a *Beta*(1, 1.5)prior, weakly favoring values below 0.5, consistent with the expectation that under-reporting is prevalent in dengue surveillance in Brazil. Estimated state- and year-specific reporting rates, and the corresponding apparent illness-to-case report ratios, are provided in **Fig. S5** and **Table S5**. Other parameters were not assigned explicit prior distributions but were constrained to be non-negative, or bounded between 0 and 1 for ratio parameters.

## SIR-type model framework

### Assumptions for model framework

Building upon the catalytic model that estimated the force of infection and case reporting ratios from 2000 to 2025, we extended the framework into a dynamic transmission model to simulate dengue spread and evaluate the potential impact of future vaccination strategies. A schematic flow diagram of the model structure is provided in **Fig. S10**, and the full parameter set is summarized in **Table S8**. The following assumptions were adopted.

1. Vector population and infection process: We assumed that mosquito dynamics adjust instantaneously relative to human infection dynamics, following a quasi–steady-state (fast reaction) approximation. As a result, vector compartments were not explicitly modeled; instead, their effects were implicitly represented through the transmission rate. Given the model’s annual time step, both intrinsic and extrinsic incubation periods (approximately 6–12 days) were considered negligible and were therefore omitted.
2. Population mixing and spatial structure: Individuals within the same age group and state were assumed to mix homogeneously and share equal exposure risk. Inter-state transmission was neglected, under the assumption that local transmission dominates over cross-state infection. Consequently, each state was modeled independently with its own demography and transmission parameters.
3. Force of infection (FOI) and transmission dynamics: The FOI in the SIR-type model is proportional to the number of infectious individuals, consistent with standard transmission formulations. To convert the catalytic model outputs into transmission parameters, we applied the endemic equilibrium (Anderson and May assumption) ^52^, allowing FOI estimates to be translated into transmission rates under equilibrium conditions. Temporary cross-protection following infection was incorporated, assuming that individuals acquire one year of short-term immunity to heterologous serotypes, after which the protection fully wanes, and subsequent infections proceed independently. This design difference is intentional: the catalytic model is a simplified inference tool focused on reconstructing cumulative infection histories from surveillance data, and cross-protection was omitted to reduce model complexity during parameter estimation; the SIR-type model, by contrast, is a transmission model intended to capture realistic dengue dynamics and therefore incorporates the short-term cross-protection that has been empirically documented following heterotypic infection.
4. Vaccine effectiveness in the model: Vaccine effectiveness (VE) was modeled on transmission, disease, hospitalization, and mortality ^33,34^. Because the model operates on an annual time scale, vaccination was assumed to induce protection immediately after administration, with no delay in onset. VE was assumed to be independent of age but dependent on serostatus, with higher effectiveness among seropositive individuals. Vaccine-derived protection wanes over time, and breakthrough infections reset an individual’s protection to its maximum value before it declines again ^33,34^.
5. VE setting: As the model does not explicitly distinguish serotypes, average VE values across the four serotypes were applied. These values were extracted separately from the serotype-specific efficacy estimates reported by Shen et al. and Daniels et al. ^33,34^. We did not differentiate VE by multitypic prior exposure, assuming that individuals with 1 to 4 prior infections have the same VE. VE against mortality was assumed to be identical to VE against hospitalization. Lastly, vaccine uptake was assumed to occur without hesitancy or logistical constraints, resulting in uniform coverage across eligible populations.

## Model notations

The *X* compartments introduced in the catalytic model were further subdivided into the *S, I*, and *R* classes. The placement and meaning of all indices followed the same conventions described earlier. Because all variables were time-dependent, the explicit time argument (*t*) was omitted for simplicity. We first define the compartments for unvaccinated individuals as follows:

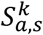: Number of susceptible individuals with *k* prior dengue infections in age group *a* in state *s*

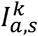: Number of infectious individuals with *k* prior dengue infections in age group *a* in state *s*

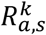 : Number of recovered individuals with *k* prior dengue infections in age group *a* in state *s*

The following parameters are defined in relation to disease transmission:

*k*_*a*_: Age group-specific multiplier (identical with those are contained in catalytic model)

β_*s*_(*t*): Time dependent transmission rate

α: Recovery rate

ω: Waning rate of cross-immunity induced by natural infection

In addition, demographic effects are incorporated into the model and are defined as follows:

*surv*_*a*_: Probability that individuals in age group *a* survive and proceed into the next age group *a* + 1

*µ*_*n*_: Natural birth rate

*µ*: Aging rate between 5-year age intervals

*µ*_*f*_: Natural death rate in the final age group (17, age over 80 years)

To incorporate the effects of vaccination and waning immunity into the state variables, we defined the notation as follows: variables with a bar (-) represent individuals within one year after vaccination (strongest); those with a dot (·) indicate individuals between one and five years after vaccination; and those with two dots (··) indicate individuals more than five years after vaccination. For example,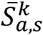indicates susceptible individuals with strong vaccine effectiveness. Following is vaccine effectiveness and waning rate. These parameters are also stratified using the same notation—bar, one dot, and two dots—to reflect different levels of vaccine-derived protection.

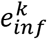 : Vaccine effectiveness against infection (acquisition) for those who have *k* prior dengue infections; values by serostatus and time since vaccination are provided in **Table S9**.”

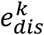 : Vaccine effectiveness against disease for those who have *k* prior dengue infections

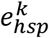: Vaccine effectiveness against hospitalization (or fatality) for those who have *k* prior dengue infections

ξ: Vaccine-induced immunity waning rate

## Model formulation

Our SIR-type model extends the catalytic modeling framework. The immune profile evolves dynamically throughout the simulation as individuals transition between compartments through natural infection and vaccination, rather than being held fixed at the 2026 initial condition. The model equations are:

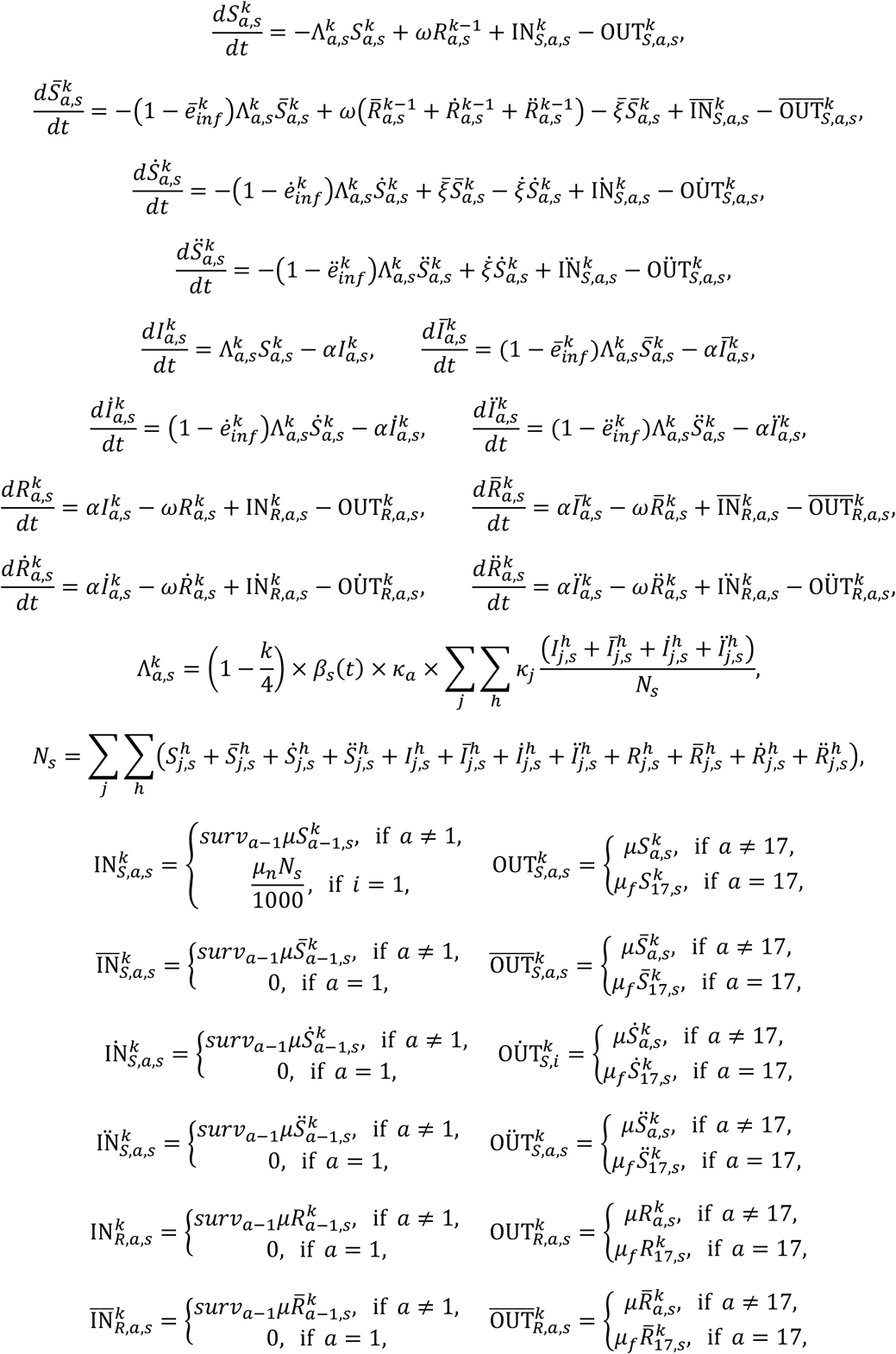

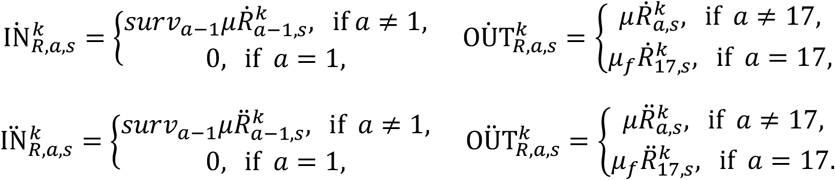

Note that demographic transition in infected individuals was not considered, as their infectious period is relatively short (a week) compared to the unit time (a year).

In this study, vaccine effectiveness estimates reported by Shen and Daniels were aggregated and incorporated into our model ^33,34^. Their studies provide serotype-specific vaccine effectiveness values; however, for the purposes of this model, we used the mean across serotypes. Aggregated vaccine effectiveness and waning dynamics are shown in **Fig. S11**, and the underlying VE estimates from both studies are summarized in **Table S9**. To map their estimates onto our modeling framework, we assigned representative point estimates to the three vaccine-derived immunity states used in this study, bar (–), one dot (·), and two dots (··), corresponding to strong immunity (<1 year post-vaccination), intermediate immunity (1–5 years), and waned immunity (>5 years), respectively. The main results presented in this study are based on vaccine effectiveness estimates from Shen, while results obtained using estimates from Daniels, along with additional details on vaccine effectiveness assumptions, are provided in the **Supplementary Appendix**.

## Initial condition and transmission rate conversion from catalytic model

To convert the catalytic model outputs into parameters for the SIR-type model, we applied the endemic equilibrium assumption following Anderson and May ^52^. The resulting state-level reproduction numbers under this assumption are shown in **Fig. S3**. Under a simplified SIR model, the endemic equilibrium yields the following relationship:

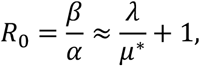

where

α is the recovery rate used in our model,

λ corresponds to the catalytic model FOI 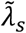(*t*),

β is matched to the transmission rate β_*s*_(*t*), and

*µ*^∗^is the inverse of the average life expectancy in Brazil (76.4 years) ^42^.

Using this relationship, we converted the catalytic-model FOI into transmission rates for SIR-type model. Let 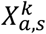 (2026)is the number of individuals in age group *a* who experienced *k* infections as of year 2026, aggregated from the final value of the catalytic model. In addition, under the same assumption, the fraction of infectious individuals (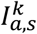 (2026)) among 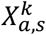 (2026)as of year 2025 is given by:

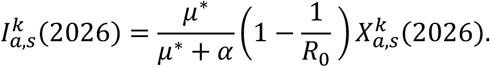

Recovered individuals were assumed to be zero at initialization, and the remaining population was assigned to the susceptible class. In this study, the SIR-type model incorporated the parameters sampled from the catalytic model as well as the median values of the corresponding state variables.

## Burden of Disease (BoD)

BoD derived from this model included cases, hospitalizations, fatalities, and DALYs. The number of cases is defined as follows (Note that the state- and time-varying reporting ratio, 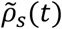, was not applied here):

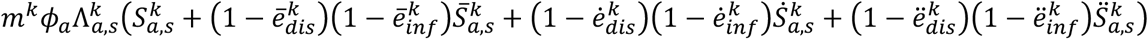

Let the parameters *p*_*h,a*_ and *p*_*f,a*_ denote the age-specific case hospitalization ratio and age-specific case fatality ratio, respectively. Hospitalizations and fatalities are defined as follows:

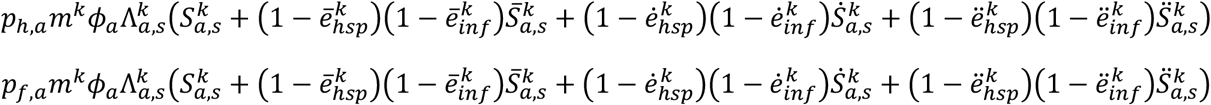

DALYs are defined as the sum of years of life lost (YLL) and years lived with disability (YLD) as follows:

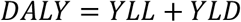

In this study, YLL was calculated by applying the life expectancy corresponding to the age group at death. Age-specific YLL values, along with case hospitalization and fatality ratios by age group, are provided in **Table S11**. YLD was computed using a per-case value of 0.00084 for general cases and a per-case value of 0.0051 for hospitalized cases. These values were derived from the Global Burden of Disease study by applying a disability weight of 0.051 for moderate infectious disease for a duration of six days for general cases, and a disability weight of 0.133, taken from the severe infectious disease category, for a duration of fourteen days for hospitalized cases ^53,54^.

## Vaccination scenarios

We used an SIR-type model to evaluate BoD over the next five years and to quantify the impact of vaccination interventions. The vaccination scenarios were organized into five sequential components. Firstly, we projected the BoD in the absence of vaccination. We then estimated the BoD averted under future vaccination coverage extrapolated from recent distribution patterns in Brazil. Next, we investigated an alternative scenario that applies an optimal vaccine allocation strategy under limited supply, followed by a scenario that targets the elderly population. Finally, we assessed the potential reductions in BoD under an accelerated vaccine production scenario. Scenarios are listed as follows:

- Baseline: Vaccine allocation followed Brazil’s observed vaccination patterns during 2024–2025, targeting individuals aged 5–14 years. The corresponding vaccine distribution data by state are summarized in **Table S10**.
- Children-targeting optimized: Vaccination targeted individuals aged 5–14 years, with state-level allocation optimized to minimize DALYs.
- Elderly-targeting optimized: Vaccination targeted individuals aged 60 years and older, with state-level allocation optimized to minimize DALYs.
- Mixed optimized strategies: A linear combination of the children-targeting and elderly-targeting optimized strategies.
- Accelerated vaccine supply: Vaccination targeted individuals aged 5–59 years, with annual vaccination capacity expanded to 10, 20, and 30 million individuals.

The accelerated supply scenarios evaluate the impact of expanded vaccination capacity per se, independent of which specific product supplies that capacity; the Butantan-DV approval and subsequent supply developments motivate the plausibility of such capacity expansion but are not used to parameterize vaccine effectiveness anywhere in this study. Vaccine effectiveness throughout this study is based on TAK-003, the only dengue vaccine with published serotype-specific efficacy estimates suitable for this modeling framework.

All scenarios from the baseline through the mixed optimized strategies assumed a limited vaccine supply of 5 million individuals per year, with vaccination randomly assigned within each target age group. Under this constrained supply, cumulative coverage over the five-year simulation period corresponded to approximately 87% of individuals aged 5–14 years or 70% of individuals aged 60 years and older, depending on the target population. In contrast, accelerated vaccine supply scenarios assumed annual vaccination capacities of 10, 20, and 30 million individuals, corresponding to coverage of up to approximately 90% of individuals aged 5–59 years.

Optimization was performed independently for each combination of projected reproduction number scenario (Q25, Q50, Q75) and vaccine effectiveness assumption (Shen, Daniels ^33,34^), yielding the ranges reported throughout the Results and **Table 1**; other catalytic-model parameters (e.g., infection-order-dependent apparent illness ratios, age-specific reporting multipliers) were fixed at their posterior median values across all optimizations, as re-optimizing across the full joint posterior was computationally prohibitive given the number of states and scenarios considered. Optimization was performed using MATLAB’s *fmincon* function in Optimization Toolbox with the interior-point algorithm, with the objective of minimizing total DALYs ^55^. Gradients were evaluated using a forward finite-difference scheme, and the Hessian matrix was approximated using the Broyden–Fletcher–Goldfarb–Shanno algorithm ^56^.

## Data Availability

All data produced are available online at https://github.com/youngsukko/denv_cat_sir_2025-2026

## Data availability

This study used only publicly available data. Dengue case notifications by state, age group and year for 2000–2025 were obtained from the National Notifiable Diseases Information System (SINAN) through the Brazilian Ministry of Health ^41^. Life tables were obtained from the Brazilian Institute of Geography and Statistics (IBGE) ^42^. Population estimates by state, five-year age group and year were obtained from the municipal population estimates by age and sex of the Brazilian Ministry of Health (DATASUS) for 2000–2021, accessed through the brpop R package, and aggregated to the state level; values for 2022–2025 were linearly extrapolated from the 2019–2021 trend ^57^. Scripts that retrieve the input data from the original sources are included in the code repository described below. Posterior parameter estimates and model outputs underlying the main figures are provided in the **Supplementary Appendix**.

## Code availability

All code used in this study is publicly available on GitHub. The catalytic model was implemented in Stan (CmdStan version 2.38.0) and fitted in R (version 4.5.3) using cmdstanr (version 0.9.0.9001) ^51^ . The SIR-type transmission model and the vaccine allocation optimization were implemented in MATLAB (version R2024b) with the Optimization Toolbox. The repository (https://github.com/youngsukko/denv_cat_sir_2025-2026) contains scripts to retrieve the input data, fit the catalytic model, run the vaccination scenarios and reproduce the figures in the main text and **Supplementary Appendix**.

## Author contributions

Y.K. and A.I.K. conceived and designed the study. Y.K. curated the data, developed the models, wrote the code, performed the analyses, prepared the figures and wrote the original draft. A.I.K. supervised the study and acquired funding. C.D.S., A.T.H., H.S. and D.A.T.C. provided methodological and epidemiological input and contributed to the interpretation of the results. All authors reviewed and revised the manuscript and approved the final version.

