## supplementary appendix for "Reconstructing Dengue Exposure Dynamics and Assessing the Public Health Impact of Vaccination Scenarios Using Mathematical Models in Brazil"

**S1. List of figures**

**S2. List of tables**

**S3. Supplementary results from the catalytic model**

**S3-1. MCMC Convergence Diagnostics**

**S3-2. Force of infection and conversion to reproductive number and projection**

**S3-3. Infection to case report ratio**

**S3-4. Immune profile and comparison with real-world surveillances**

**S4. Supplementary figures and tables for the SIR-type model framework**

**S5. Supplementary SIR-type model outputs**

**S5-1. Projection of BoD without vaccination**

**S5-2. Projection of BoD under different vaccine effectiveness setting (baseline scenario)**

**S5-3. Optimal state-level vaccine allocation targeting children (or elderly)**

**S6. References**

**S1. List of figures**

**Fig. S1**: Time-dependent, per-serotype force of infection (FOI) by Brazilian state

**Fig. S2**: Pre-2000 force of infection by Brazilian state. Colors of bar indicate the geographical regions in Brazil.

**Fig. S3**: Reproductive number under the endemic equilibrium assumption.

**Fig. S4**: Infection-to-apparent illness ratios by infection order (primary, secondary, tertiary, quaternary).

**Fig. S5**: Apparent illness-to-case report ratio by state and year.

**Fig. S6**: Model-fitted case incidence across states in Brazil using the catalytic modeling framework.

**Fig. S7**: Case/Infection across states in Brazil. Color of box chart indicates the geographical region.

**Fig. S8**: Temporal evolution of the age-specific proportion of individuals with fewer than two prior dengue infections.

**Fig. S9**: Comparison of model-based immune profile estimates with real-world serosurvey data across Brazilian states.

**Fig. S10**: Flow diagram of the SIR-type mathematical model.

**Fig. S11:** Aggregated vaccine effectiveness (markers) and waning dynamics from the model (solid curve).

**Fig. S12**: Proportion of BoD by age group in different geographical regions.

**Fig. S13**: Burden of disease (BoD) averted per 1,000 vaccinated individuals under the projected vaccine distribution.

**Fig. S14**: Optimized vaccine allocation results under Shen's vaccine effectiveness assumptions.

**Fig. S15**: Optimized vaccine allocation results under Daniels' vaccine effectiveness assumptions.

**Fig. S16**: Choropleth maps of optimized vaccine allocation under Shen's vaccine effectiveness assumptions.

**Fig. S17**: Choropleth maps of optimized vaccine allocation under Daniels' vaccine effectiveness assumptions.

**Fig. S18**: Optimal vaccine allocation under Daniels- and Shen-derived vaccine efficacy (VE) assumptions.

**S2. List of tables**


**Table S1**: Age group–specific multipliers for the force of infection.

**Table S2**: Force of infection (median and 95% CrI).

**Table S3**: Posterior medians and 95% credible intervals for sequence-dependent infection-to-apparent illness ratios.

**Table S4**: Posterior medians and 95% credible intervals for age group–specific adjustment multipliers for apparent illness.

**Table S5**: Apparent illness-to-case report ratio (%) by state and year (median and 95% CrI).

**Table S6**: Case/Infection ratio across states in Brazil.

**Table S7**: Immune profiles showing the proportion of the population exposed to DENV (%) in 2000 and 2026.

**Table S8**: SIR-type model parameters.

**Table S9**: Vaccine effectiveness setting from two studies.

**Table S10**: The quantity of vaccine distributed over the 2024/25 period in Brazil.

**Table S11**: Years of life lost (YLL) and case hospitalization/fatality ratio by age group.

**Table S12**: Projected annual burden of disease (BoD) per 100,000 population by state without vaccine administration.

### S3. Supplementary results from the catalytic model

#### S3-1. MCMC Convergence Diagnostics

Posterior sampling was performed using four independent chains. Convergence was evaluated using R-hat statistics (threshold < 1.05), bulk and tail effective sample sizes (ESS; threshold > 400), the number of divergent transitions, and the E-BFMI (threshold > 0.3). No divergent transitions were observed across any chain, and no transitions exceeded the maximum tree depth. E-BFMI values ranged from 0.848 to 1.028 across the four chains, all well above the recommended threshold. R-hat values were ≤ 1.01 for all parameters, indicating satisfactory mixing. Bulk ESS values were generally well above 400; two exceptions were delta[3] and delta[4] (apparent illness ratios for tertiary and quaternary infections; bulk ESS 523 and 619, respectively), reflecting the limited number of individuals with three or more prior infections in the data. The tail ESS for the pre-2000 FOI in Bahia (BA) was 304, marginally below threshold, consistent with greater posterior uncertainty in a state with complex early transmission history. Neither issue indicated systematic sampling failure.

#### S3-2. Force of infection and conversion to reproductive number and projection

**Table S1** lists the age group–specific FOI multipliers ($\kappa_a$) relative to the 0–4 age group. The time-dependent FOI by state varied substantially across both state and year. **Fig. S1** visualizes these estimates using a panel layout arranged in the geographic shape of Brazil. Background colors indicate the geographical regions (north, northeast, central-west, southeast, and south) each states belong. Also, dotted dark curves in each panel indicate the incidence per 100 population. **Fig. S2** presents the pre-2000 FOI values as box plots, with colors indicating the corresponding geographical regions. The northeast and several states in the north and southeast had high pre-2000 FOI, whereas the southern region had low pre-2000 FOI. These values are summarized in **Table S2**.

**Table S1**: Age group–specific multipliers for the force of infection.

| Age group | Median, 95% CrI |
|---|---|
| **5-9y** | 1.63 [0.87, 3.03] |
| **10-14y** | 2.18 [1.11, 3.73] |
| **15-19y** | 2.21 [1.21, 3.74] |
| **20-24y** | 2.22 [1.24, 3.59] |
| **25-29y** | 2.21 [1.24, 3.68] |
| **30-34y** | 2.24 [1.26, 3.75] |
| **35-39y** | 2.20 [1.22, 3.61] |
| **40-44y** | 2.23 [1.26, 3.61] |
| **45-49y** | 2.18 [1.20, 3.61] |
| **50-54y** | 2.17 [1.21, 3.48] |
| **55-59y** | 1.96 [1.12, 3.22] |
| **60-64y** | 1.79 [0.99, 2.98] |
| **65-69y** | 1.65 [0.90, 2.86] |
| **70-74y** | 1.46 [0.80, 2.52] |
| **75-79y** | 1.30 [0.70, 2.37] |
| **≥80y** | 1.05 [0.58, 1.87] |

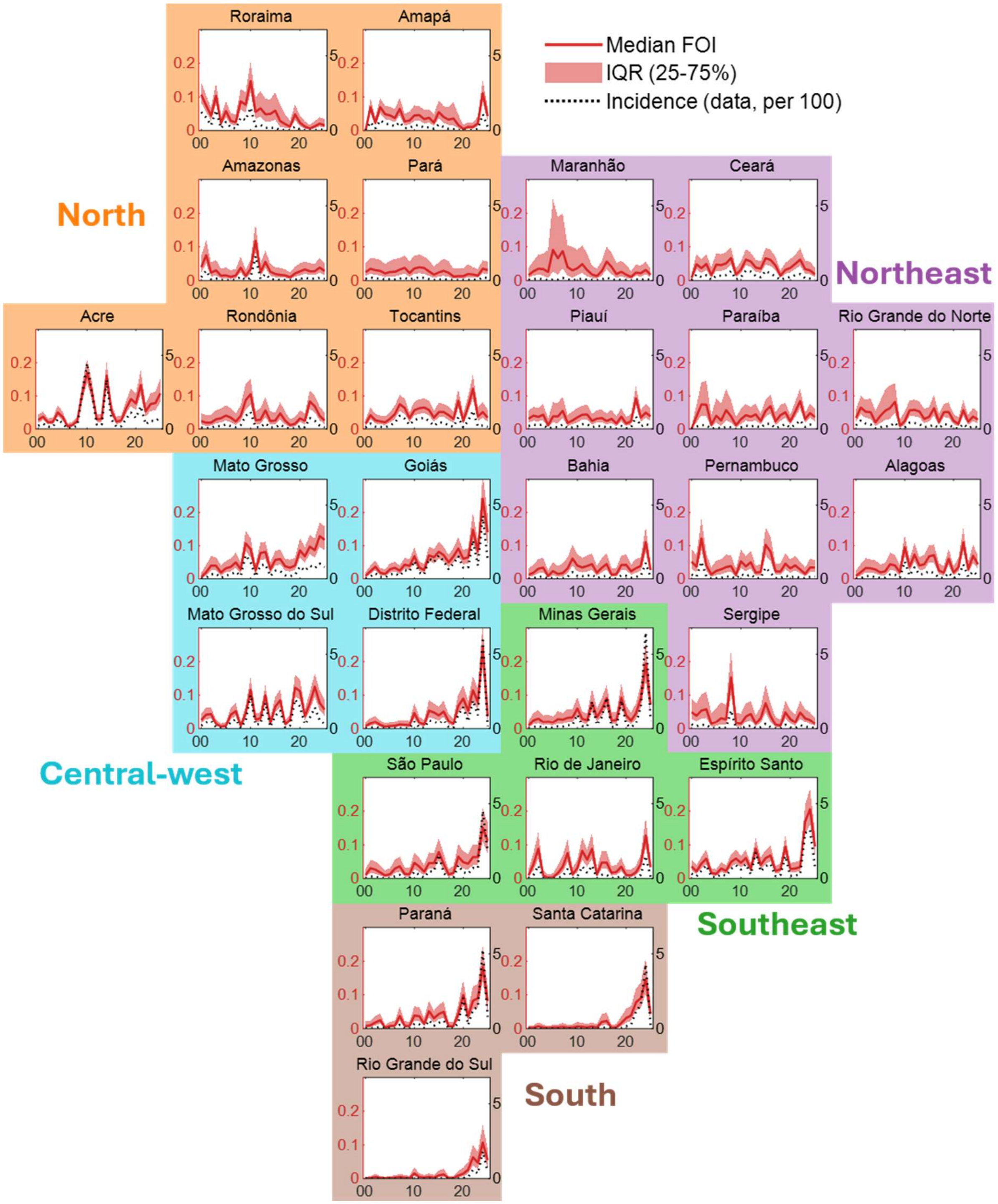


**Fig. S1**: Time-dependent, per-serotype force of infection (FOI) by Brazilian state. Solid red curves indicate the median estimated FOI, and shaded regions represent the 95% credible intervals. Black dashed lines denote reported dengue incidence per 100 individuals. In each panel, the left y-axis corresponds to the per-serotype FOI, while the right y-axis shows dengue incidence per 100 individuals. Panels are arranged by geographical region, and the x-axis represents calendar year. Note that FOI in each panel represents FOI causing primary infection, i.e. $4\tilde{\lambda}_s(t)$.

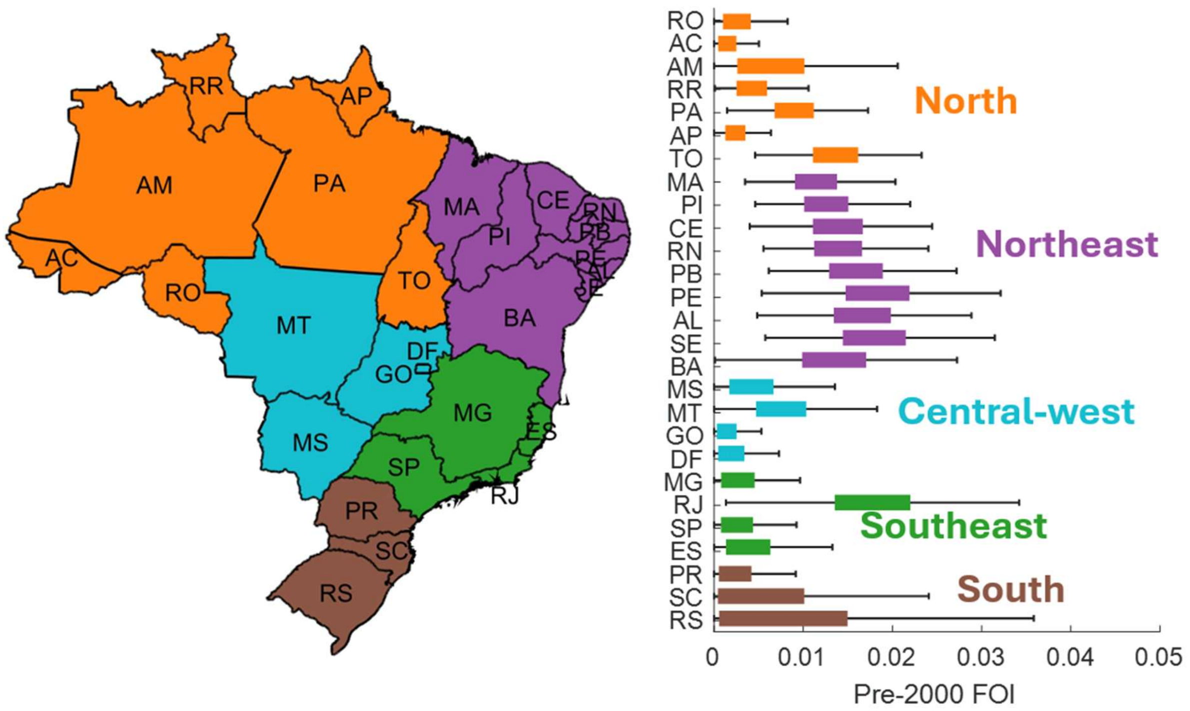


**Fig. S2**: Pre-2000 force of infection by Brazilian state. Colors of bar indicate the geographical regions in Brazil.

**Table S2**: Force of infection (median and 95% CrI). Values are scaled by a factor of 100 (e.g., 0.02 is shown as 2 in the table).

| Year | Pre-00 | 00 | 01 | 02 | 03 | 04 | 05 | 06 | 07 | 08 | 09 | 10 | 11 | 12 | 13 | 14 | 15 | 16 | 17 | 18 | 19 | 20 | 21 | 22 | 23 | 24 | 25 |
|---|---|---|---|---|---|---|---|---|---|---|---|---|---|---|---|---|---|---|---|---|---|---|---|---|---|---|---|
| **RO** | 0.24 [0, 0] | 2.36 [1, 5] | 1.88 [1, 4] | 1.93 [1, 4] | 2.92 [2, 6] | 3.24 [2, 6] | 4.05 [3, 7] | 3.42 [2, 6] | 2.26 [1, 5] | 3.95 [2, 7] | 8.67 [6, 14] | 10.50 [7, 15] | 2.65 [1, 5] | 2.78 [1, 6] | 5.62 [4, 10] | 2.40 [1, 5] | 2.54 [1, 5] | 4.71 [3, 8] | 2.54 [1, 5] | 1.19 [1, 3] | 1.58 [1, 4] | 3.17 [2, 6] | 2.47 [1, 5] | 8.31 [6, 12] | 6.83 [5, 10] | 3.96 [3, 7] | 2.54 [1, 5] |
| **AC** | 0.13 [0, 0] | 2.73 [2, 4] | 3.69 [3, 5] | 2.00 [1, 3] | 2.13 [1, 3] | 4.98 [4, 7] | 3.61 [3, 5] | 0.80 [0, 2] | 1.16 [1, 2] | 2.44 [2, 4] | 10.65 [8, 14] | 16.98 [14, 21] | 11.13 [9, 14] | 2.88 [2, 4] | 3.04 [2, 5] | 16.08 [13, 20] | 5.41 [4, 8] | 2.24 [1, 4] | 2.55 [2, 4] | 6.17 [5, 8] | 9.04 [7, 12] | 7.70 [6, 11] | 13.26 [10, 18] | 5.29 [4, 8] | 7.51 [5, 11] | 7.82 [6, 11] | 10.75 [8, 15] |
| **AM** | 0.61 [0, 1] | 3.96 [2, 7] | 7.51 [5, 12] | 2.34 [1, 5] | 3.00 [1, 6] | 1.42 [1, 4] | 1.51 [1, 4] | 1.10 [0, 3] | 1.48 [1, 3] | 3.81 [2, 7] | 1.37 [1, 3] | 3.43 [2, 6] | 11.75 [9, 17] | 2.45 [1, 5] | 5.52 [4, 9] | 2.59 [1, 5] | 1.93 [1, 4] | 1.67 [1, 4] | 1.51 [1, 3] | 0.95 [0, 2] | 2.09 [1, 4] | 2.71 [1, 5] | 3.28 [2, 6] | 2.75 [2, 5] | 2.76 [2, 5] | 3.84 [2, 7] | 2.82 [2, 5] |
| **RR** | 0.42 [0, 1] | 10.70 [8, 14] | 7.42 [5, 10] | 4.39 [3, 7] | 10.31 [8, 14] | 2.31 [1, 4] | 5.80 [4, 9] | 2.87 [2, 5] | 2.51 [1, 5] | 8.68 [6, 12] | 7.68 [5, 11] | 14.70 [11, 20] | 5.54 [3, 10] | 6.61 [4, 12] | 4.85 [3, 9] | 4.92 [3, 9] | 5.92 [3, 11] | 2.81 [1, 8] | 1.93 [1, 5] | 1.16 [0, 3] | 4.80 [3, 8] | 2.32 [1, 4] | 1.18 [1, 3] | 0.61 [0, 2] | 1.26 [1, 3] | 2.06 [1, 4] | 1.54 [1, 3] |
| **PA** | 0.88 [1, 1] | 2.48 [1, 5] | 3.47 [2, 6] | 3.07 [2, 6] | 2.85 [1, 6] | 2.08 [1, 5] | 2.54 [1, 6] | 2.56 [1, 6] | 3.22 [2, 7] | 3.77 [2, 7] | 2.23 [1, 6] | 3.57 [2, 7] | 3.92 [2, 8] | 3.72 [2, 7] | 2.68 [1, 6] | 1.73 [1, 4] | 2.39 [1, 5] | 2.87 [1, 6] | 2.29 [1, 6] | 1.21 [0, 4] | 1.26 [1, 3] | 1.28 [0, 3] | 1.93 [1, 5] | 1.77 [1, 4] | 1.46 [1, 3] | 3.49 [2, 6] | 3.10 [2, 6] |
| **AP** | 0.23 [0, 0] | 0.03 [0, 0] | 6.92 [5, 10] | 2.29 [1, 4] | 6.95 [5, 10] | 4.87 [3, 7] | 4.50 [3, 7] | 3.72 [2, 6] | 6.46 [4, 10] | 2.72 [2, 5] | 2.98 [2, 6] | 4.50 [3, 7] | 4.57 [3, 7] | 3.40 [2, 6] | 3.78 [2, 6] | 2.37 [1, 5] | 5.50 [4, 9] | 4.01 [2, 7] | 3.25 [2, 7] | 3.59 [2, 8] | 1.64 [1, 5] | 0.49 [0, 2] | 1.06 [1, 2] | 1.08 [0, 2] | 2.12 [1, 4] | 11.10 [9, 15] | 4.29 [3, 7] |
| **TO** | 1.33 [1, 2] | 1.60 [1, 3] | 4.16 [3, 6] | 2.61 [2, 5] | 2.29 [1, 4] | 2.02 [1, 4] | 3.08 [2, 5] | 4.50 [3, 7] | 7.47 [5, 10] | 6.35 [4, 9] | 3.24 [2, 5] | 5.66 [4, 8] | 6.25 [5, 9] | 6.48 [5, 9] | 5.72 [4, 9] | 3.40 [2, 5] | 5.10 [3, 8] | 5.11 [4, 8] | 4.18 [3, 7] | 3.02 [2, 5] | 8.52 [6, 12] | 2.41 [1, 5] | 6.81 [5, 10] | 12.17 [9, 16] | 3.78 [2, 6] | 5.10 [3, 8] | 3.41 [2, 6] |
| **MA** | 1.12 [1, 1] | 1.60 [1, 4] | 2.74 [1, 6] | 3.48 [2, 8] | 3.39 [1, 8] | 2.53 [1, 9] | 9.10 [3, 24] | 6.62 [2, 19] | 8.72 [4, 20] | 3.94 [1, 10] | 3.01 [1, 10] | 3.64 [1, 9] | 4.86 [2, 10] | 3.08 [1, 8] | 1.77 [1, 4] | 1.31 [1, 3] | 2.82 [1, 6] | 5.63 [3, 10] | 4.07 [2, 8] | 1.91 [1, 5] | 2.59 [1, 6] | 1.73 [1, 5] | 1.06 [0, 3] | 2.36 [1, 5] | 2.18 [1, 4] | 3.13 [2, 6] | 1.73 [1, 3] |
| **PI** | 1.24 [1, 1] | 3.07 [2, 5] | 4.03 [2, 7] | 3.63 [2, 6] | 4.32 [3, 8] | 1.13 [0, 3] | 3.76 [2, 8] | 3.76 [2, 8] | 5.50 [3, 10] | 1.84 [1, 4] | 2.54 [1, 5] | 3.50 [2, 6] | 4.29 [3, 7] | 4.38 [3, 7] | 2.43 [1, 5] | 3.11 [2, 6] | 3.30 [2, 6] | 2.43 [1, 5] | 3.03 [2, 6] | 1.66 [1, 4] | 3.71 [2, 6] | 1.65 [1, 4] | 2.33 [1, 4] | 9.23 [7, 13] | 3.42 [2, 6] | 4.94 [3, 8] | 3.82 [2, 6] |
| **CE** | 1.36 [1, 2] | 0.77 [0, 2] | 4.82 [3, 8] | 3.67 [2, 6] | 4.91 [3, 8] | 1.60 [1, 4] | 4.71 [3, 8] | 4.58 [3, 8] | 5.09 [3, 8] | 6.67 [5, 10] | 1.78 [1, 4] | 3.22 [2, 6] | 6.22 [4, 9] | 5.65 [4, 8] | 3.85 [3, 6] | 3.51 [2, 6] | 6.61 [5, 10] | 6.11 [4, 9] | 4.45 [3, 7] | 1.43 [1, 3] | 3.15 [2, 5] | 3.74 [2, 6] | 4.95 [3, 8] | 6.21 [4, 10] | 3.39 [2, 6] | 2.99 [2, 6] | 1.83 [1, 4] |
| **RN** | 1.36 [1, 2] | 3.34 [2, 6] | 6.46 [4, 10] | 5.20 [3, 9] | 5.06 [3, 9] | 2.01 [1, 6] | 3.97 [2, 9] | 5.49 [3, 12] | 6.32 [3, 13] | 8.18 [5, 14] | 0.97 [0, 3] | 2.37 [1, 5] | 5.63 [4, 9] | 5.74 [4, 9] | 5.30 [3, 8] | 3.02 [2, 6] | 3.67 [2, 7] | 6.27 [4, 10] | 2.13 [1, 5] | 5.06 [4, 8] | 5.13 [3, 8] | 2.46 [1, 5] | 1.50 [1, 3] | 5.66 [4, 9] | 2.18 [1, 4] | 3.75 [2, 6] | 2.82 [2, 5] |
| **PB** | 1.55 [1, 2] | 0.06 [0, 1] | 3.76 [2, 7] | 7.39 [4, 14] | 7.32 [4, 14] | 1.29 [0, 4] | 3.99 [2, 9] | 2.57 [1, 8] | 5.69 [3, 12] | 3.80 [2, 9] | 1.16 [0, 4] | 2.92 [1, 6] | 4.18 [2, 7] | 3.68 [2, 7] | 4.40 [3, 8] | 2.42 [1, 5] | 5.50 [4, 9] | 6.87 [5, 10] | 2.31 [1, 5] | 4.14 [2, 7] | 5.05 [3, 8] | 2.32 [1, 4] | 5.24 [4, 8] | 8.41 [6, 12] | 2.96 [2, 5] | 4.80 [3, 8] | 3.57 [2, 6] |
| **PE** | 1.79 [1, 2] | 5.19 [3, 9] | 3.69 [2, 7] | 12.20 [9, 18] | 4.04 [2, 8] | 1.34 [1, 3] | 2.41 [1, 5] | 2.91 [1, 6] | 3.56 [2, 7] | 3.87 [2, 7] | 1.16 [0, 3] | 5.33 [3, 9] | 3.79 [2, 7] | 5.03 [3, 9] | 1.91 [1, 4] | 2.06 [1, 4] | 10.28 [7, 15] | 8.19 [5, 13] | 2.31 [1, 5] | 2.22 [1, 5] | 4.59 [3, 8] | 3.20 [2, 6] | 4.10 [3, 7] | 2.82 [2, 5] | 1.52 [1, 3] | 3.43 [2, 6] | 3.48 [2, 6] |
| **AL** | 1.63 [1, 2] | 0.92 [0, 3] | 2.01 [1, 4] | 3.30 [2, 6] | 3.20 [2, 6] | 2.85 [2, 6] | 1.96 [1, 4] | 2.30 [1, 5] | 4.43 [3, 8] | 4.07 [2, 7] | 1.99 [1, 4] | 9.55 [7, 13] | 3.68 [2, 6] | 7.34 [5, 11] | 4.05 [3, 7] | 4.42 [3, 7] | 6.81 [5, 10] | 7.16 [5, 10] | 2.49 [1, 5] | 1.89 [1, 4] | 6.06 [4, 9] | 1.92 [1, 4] | 3.51 [2, 6] | 11.01 [8, 15] | 2.91 [2, 5] | 6.96 [5, 10] | 4.41 [3, 7] |
| **SE** | 1.76 [1, 2] | 4.96 [3, 9] | 3.91 [2, 8] | 5.42 [3, 10] | 5.84 [3, 11] | 1.44 [1, 5] | 2.00 [1, 6] | 3.17 [1, 8] | 2.75 [1, 7] | 15.29 [10, 23] | 1.72 [1, 5] | 1.47 [1, 4] | 3.65 [2, 8] | 4.42 [2, 8] | 1.52 [1, 4] | 3.18 [2, 6] | 7.64 [5, 13] | 3.82 [2, 8] | 1.17 [0, 3] | 0.86 [0, 3] | 4.90 [3, 8] | 2.90 [2, 6] | 1.85 [1, 4] | 4.84 [3, 8] | 3.13 [2, 6] | 2.59 [1, 5] | 1.42 [1, 3] |
| **BA** | 1.32 [1, 2] | 1.27 [0, 3] | 1.86 [1, 4] | 3.09 [2, 6] | 3.41 [2, 6] | 0.91 [0, 3] | 2.34 [1, 5] | 1.52 [1, 4] | 1.76 [1, 5] | 3.20 [2, 6] | 6.15 [4, 10] | 4.01 [2, 8] | 3.45 [2, 6] | 3.90 [2, 7] | 4.47 [3, 8] | 1.87 [1, 4] | 3.38 [2, 6] | 4.50 [3, 8] | 1.28 [1, 3] | 1.43 [1, 3] | 4.19 [3, 7] | 4.26 [3, 7] | 2.30 [1, 4] | 3.23 [2, 6] | 3.93 [2, 7] | 11.11 [8, 16] | 2.69 [2, 5] |
| **MS** | 0.40 [0, 1] | 2.60 [2, 4] | 4.22 [3, 6] | 4.39 [3, 6] | 1.42 [1, 3] | 0.56 [0, 1] | 0.76 [0, 2] | 4.22 [3, 6] | 5.26 [4, 8] | 0.81 [0, 2] | 4.61 [3, 7] | 11.57 [9, 15] | 2.73 [2, 4] | 3.13 [2, 5] | 9.96 [8, 13] | 1.64 [1, 3] | 6.61 [5, 10] | 8.48 [6, 12] | 1.30 [1, 3] | 2.66 [2, 4] | 12.24 [10, 16] | 11.20 [9, 15] | 3.80 [2, 6] | 7.62 [5, 11] | 12.53 [10, 16] | 7.71 [5, 11] | 5.73 [4, 9] |
| **MT** | 0.75 [0, 1] | 0.53 [0, 2] | 2.02 [1, 4] | 4.05 [3, 7] | 3.96 [3, 7] | 1.94 [1, 4] | 3.21 [2, 5] | 3.66 [2, 6] | 4.99 [3, 8] | 2.89 [2, 5] | 10.72 [8, 14] | 8.96 [7, 12] | 2.52 [1, 4] | 7.80 [6, 11] | 8.16 [6, 11] | 2.90 [2, 5] | 5.74 [4, 8] | 5.99 [4, 9] | 3.81 [2, 6] | 3.39 [2, 6] | 4.04 [3, 6] | 8.62 [6, 12] | 6.82 [5, 10] | 9.76 [7, 13] | 8.65 [6, 12] | 12.90 [10, 17] | 11.80 [9, 16] |
| **GO** | 0.11 [0, 0] | 0.93 [0, 2] | 2.34 [1, 4] | 3.68 [3, 5] | 1.86 [1, 3] | 1.49 [1, 3] | 2.92 [2, 5] | 3.36 [2, 5] | 2.48 [2, 4] | 4.30 [3, 6] | 3.95 [3, 6] | 6.84 [5, 9] | 3.41 [2, 5] | 2.71 [2, 4] | 7.09 [5, 10] | 6.55 [5, 9] | 8.20 [6, 11] | 7.15 [5, 10] | 4.99 [3, 7] | 6.82 [5, 9] | 9.16 [7, 13] | 6.09 [4, 9] | 6.49 [5, 9] | 14.82 [11, 20] | 8.30 [6, 12] | 24.18 [19, 31] | 14.09 [10, 20] |
| **DF** | 0.16 [0, 0] | 0.73 [0, 2] | 1.56 [1, 3] | 2.05 [1, 4] | 1.07 [0, 2] | 0.52 [0, 2] | 0.72 [0, 2] | 0.82 [0, 2] | 1.19 [1, 3] | 1.08 [0, 3] | 0.96 [0, 2] | 4.66 [3, 8] | 1.78 [1, 4] | 1.18 [1, 3] | 5.39 [3, 9] | 4.89 [3, 8] | 4.00 [2, 7] | 5.16 [3, 8] | 2.27 [1, 4] | 1.48 [1, 3] | 6.85 [5, 10] | 9.00 [6, 13] | 5.42 [3, 8] | 11.48 [8, 16] | 7.91 [5, 12] | 24.17 [19, 31] | 3.75 [2, 7] |
| **MG** | 0.22 [0, 0] | 0.88 [0, 2] | 2.48 [1, 5] | 3.04 [2, 6] | 1.97 [1, 4] | 2.09 [1, 4] | 1.85 [1, 4] | 2.87 [1, 5] | 2.70 [1, 5] | 3.27 [2, 6] | 3.39 [2, 6] | 6.05 [4, 9] | 2.96 [2, 5] | 2.56 [1, 5] | 7.78 [5, 12] | 3.48 [2, 6] | 5.82 [4, 9] | 9.15 [6, 13] | 2.47 [1, 5] | 2.62 [1, 5] | 9.27 [6, 13] | 4.03 [2, 7] | 2.43 [1, 5] | 4.63 [3, 8] | 9.15 [6, 13] | 19.36 [14, 26] | 7.23 [4, 11] |
| **ES** | 0.34 [0, 1] | 3.57 [2, 6] | 2.14 [1, 4] | 4.43 [3, 6] | 5.88 [4, 9] | 1.62 [1, 3] | 1.38 [1, 3] | 2.95 [2, 5] | 2.26 [1, 4] | 4.92 [3, 7] | 5.93 [4, 9] | 4.66 [3, 7] | 6.09 [4, 8] | 2.86 [2, 4] | 8.86 [7, 12] | 4.00 [3, 6] | 6.05 [4, 8] | 6.84 [5, 10] | 2.25 [1, 4] | 2.70 [2, 4] | 9.44 [7, 13] | 2.81 [2, 5] | 2.95 [2, 5] | 3.41 [2, 5] | 16.91 [13, 22] | 20.58 [16, 27] | 9.51 [7, 14] |
| **RJ** | 1.74 [1, 2] | 0.93 [0, 3] | 4.25 [2, 8] | 8.86 [6, 14] | 0.81 [0, 3] | 0.20 [0, 1] | 0.32 [0, 1] | 1.63 [1, 4] | 3.50 [2, 7] | 7.52 [5, 12] | 1.23 [0, 3] | 3.12 [2, 6] | 8.22 [5, 12] | 5.58 [3, 9] | 8.82 [6, 13] | 1.13 [0, 3] | 4.75 [3, 8] | 4.71 [3, 8] | 1.69 [1, 4] | 1.96 [1, 4] | 3.50 [2, 6] | 1.01 [0, 3] | 0.93 [0, 3] | 1.85 [1, 4] | 4.32 [3, 7] | 12.75 [9, 18] | 3.36 [2, 6] |
| **SP** | 0.21 [0, 0] | 1.04 [0, 3] | 3.05 [2, 6] | 2.64 [1, 5] | 1.79 [1, 4] | 0.90 [0, 2] | 1.28 [0, 3] | 2.92 [2, 6] | 3.61 [2, 6] | 1.34 [1, 3] | 1.31 [1, 3] | 4.64 [3, 8] | 3.51 [2, 6] | 2.00 [1, 4] | 4.61 [3, 8] | 4.53 [3, 8] | 7.85 [5, 12] | 4.33 [3, 7] | 1.20 [0, 3] | 1.69 [1, 4] | 6.49 [4, 10] | 4.84 [3, 8] | 4.38 [3, 7] | 6.36 [4, 10] | 6.41 [4, 10] | 15.07 [11, 21] | 11.22 [7, 17] |
| **PR** | 0.18 [0, 0] | 0.72 [0, 2] | 0.97 [0, 2] | 1.81 [1, 4] | 2.47 [1, 4] | 0.24 [0, 1] | 0.74 [0, 2] | 0.85 [0, 2] | 3.73 [2, 6] | 0.78 [0, 2] | 0.76 [0, 2] | 3.84 [2, 7] | 3.66 [2, 6] | 1.36 [1, 3] | 5.17 [3, 8] | 3.03 [2, 5] | 4.32 [3, 7] | 4.93 [3, 8] | 0.69 [0, 2] | 0.69 [0, 2] | 4.42 [3, 7] | 9.93 [7, 14] | 3.60 [2, 6] | 8.26 [6, 12] | 9.00 [6, 13] | 18.36 [14, 24] | 8.15 [5, 12] |
| **SC** | 0.20 [0, 1] | 0.21 [0, 1] | 0.23 [0, 1] | 0.63 [0, 2] | 0.29 [0, 1] | 0.15 [0, 1] | 0.30 [0, 1] | 0.31 [0, 1] | 0.51 [0, 2] | 0.27 [0, 1] | 0.32 [0, 1] | 0.59 [0, 2] | 0.51 [0, 2] | 0.35 [0, 1] | 0.60 [0, 2] | 0.32 [0, 1] | 1.95 [1, 4] | 2.28 [1, 5] | 0.21 [0, 1] | 0.36 [0, 1] | 1.63 [1, 4] | 3.13 [2, 6] | 3.86 [2, 7] | 7.76 [5, 12] | 9.03 [6, 14] | 14.56 [10, 21] | 4.31 [2, 7] |
| **RS** | 0.28 [0, 1] | 0.09 [0, 1] | 0.14 [0, 1] | 0.44 [0, 1] | 0.18 [0, 1] | 0.09 [0, 1] | 0.13 [0, 1] | 0.23 [0, 1] | 0.44 [0, 1] | 0.23 [0, 1] | 0.15 [0, 1] | 1.51 [1, 3] | 0.45 [0, 1] | 0.25 [0, 1] | 0.47 [0, 1] | 0.23 [0, 1] | 0.93 [0, 2] | 1.22 [1, 3] | 0.11 [0, 1] | 0.13 [0, 1] | 0.90 [0, 2] | 1.43 [1, 3] | 2.64 [1, 5] | 6.33 [4, 10] | 4.69 [3, 8] | 10.70 [7, 16] | 5.48 [3, 9] |

**Fig. S3** presents the estimated and projected reproduction numbers with median values and IQRs, based on the previously described FOI and endemic equilibrium assumption. In this process, Gaussian process regression was utilized. These projected values are subsequently used to inform the transmission rate ($\beta_S(t)$) in the SIR-type model.

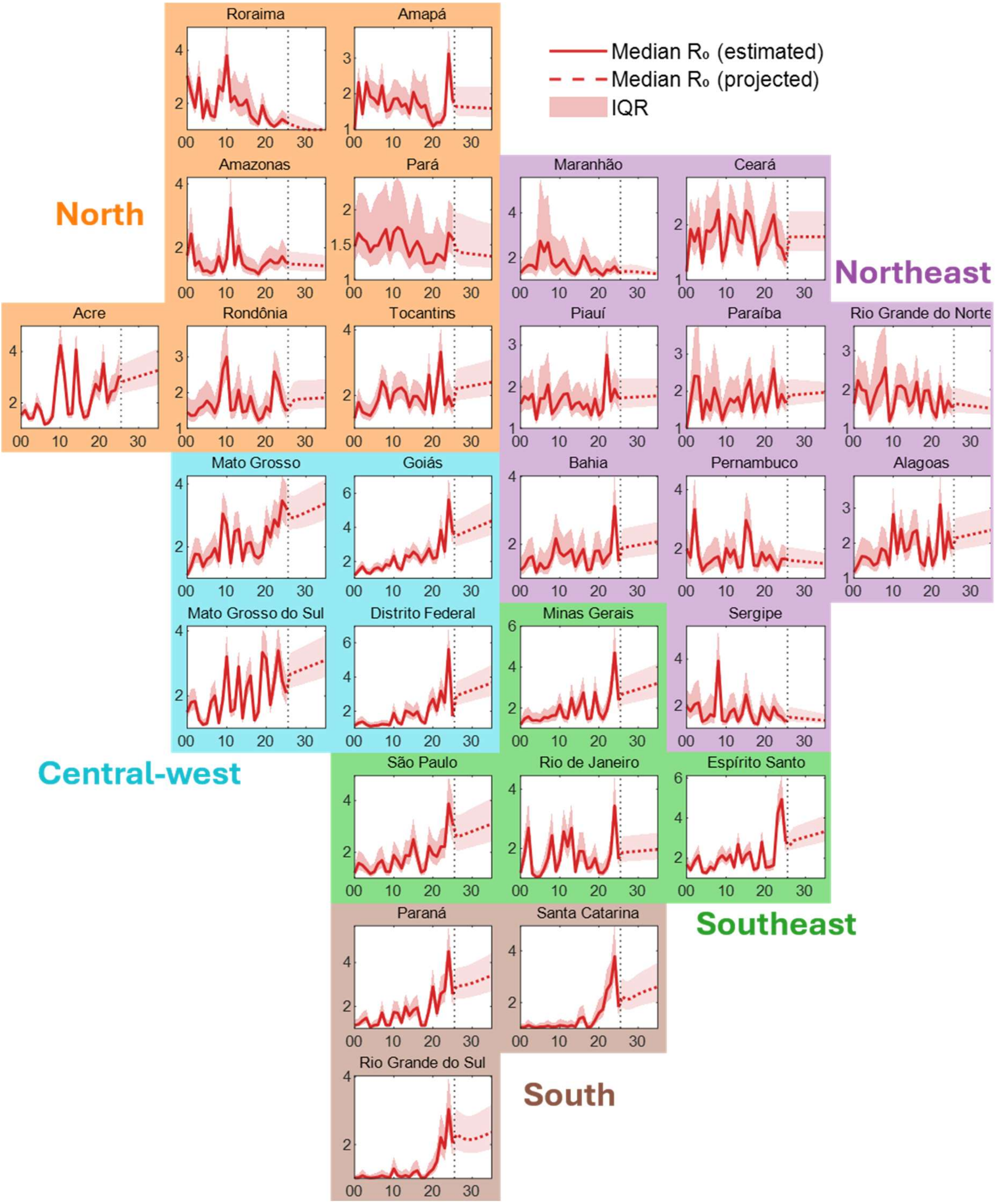


**Fig. S3**: Reproductive number under the endemic equilibrium assumption, based on estimated force of infection, and projections for the next five years using Gaussian process regression.

**S3-3. Infection to case report ratio**

**Fig. S4** visualizes the estimated sequence-dependent infection-to-apparent illness ratios. For primary infections, the posterior estimates were slightly lower than the prior mean of 0.2, with a median 0.15 (95% CrI [0.11, 0.18]). For secondary infections, the estimates were close to the prior mean of 0.5, with a median 0.49 (95% CrI [0.43, 0.56]). **Table S3** lists these values in detail. Age-specific reporting-rate adjustment multipliers are now shown in the main text (**Fig. 2C**), and are lowest in age group 10–14y with a characteristic V-shaped pattern across ages; detailed values are provided in **Table S4**.

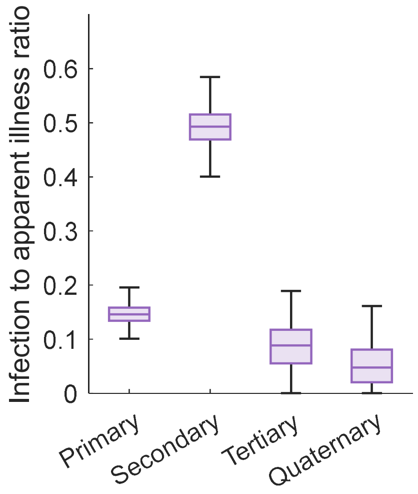


**Fig. S4**: Infection-to-apparent illness ratio by infection order (primary, secondary, tertiary, quaternary).

**Table S3**: Posterior medians and 95% credible intervals for sequence-dependent infection-to-apparent illness ratios (primary, secondary, tertiary, and quaternary infections).

| Sequence | Median, 95% CrI |
|---|---|
| **Primary** | 0.15 [0.11, 0.18] |
| **Secondary** | 0.49 [0.43, 0.56] |
| **Tertiary** | 0.09 [0.01, 0.16] |
| **Quaternary** | 0.05 [0.00, 0.13] |

**Table S4**: Posterior medians and 95% credible intervals for age group–specific adjustment multipliers for apparent illness and healthcare-seeking behavior.

| Age group | Median, 95% CrI |
|---|---|
| **0-4y** | 0.74 [0.43, 0.98] |
| **5-9y** | 0.42 [0.25, 0.72] |
| **10-14y** | 0.39 [0.26, 0.59] |
| **15-19y** | 0.48 [0.34, 0.69] |
| **20-24y** | 0.57 [0.42, 0.77] |
| **25-29y** | 0.62 [0.44, 0.84] |
| **30-34y** | 0.63 [0.46, 0.85] |
| **35-39y** | 0.65 [0.46, 0.89] |
| **40-44y** | 0.66 [0.47, 0.87] |
| **45-49y** | 0.68 [0.49, 0.91] |
| **50-54y** | 0.70 [0.51, 0.90] |
| **55-59y** | 0.71 [0.50, 0.94] |
| **60-64y** | 0.73 [0.53, 0.96] |
| **65-69y** | 0.74 [0.53, 0.96] |
| **70-74y** | 0.75 [0.50, 0.97] |
| **75-79y** | 0.75 [0.48, 0.97] |
| **≥80y** | 0.71 [0.51, 0.95] |

**Fig. S5** presents the time-dependent apparent illness–to–case report ratio by state, using the same panel layout as shown previously. Due to the influence of the prior, many estimates remained below 50%, while certain states exhibited relatively higher values (e.g., Acre or Distrito Federal after late 2010s). Across the central-western to southern states, the ratio showed a gradual increasing trend over time. These values are listed in **Table S5**. **Fig. S6** presents the state-specific incidence fitted with the population, FOI, and reporting ratios.

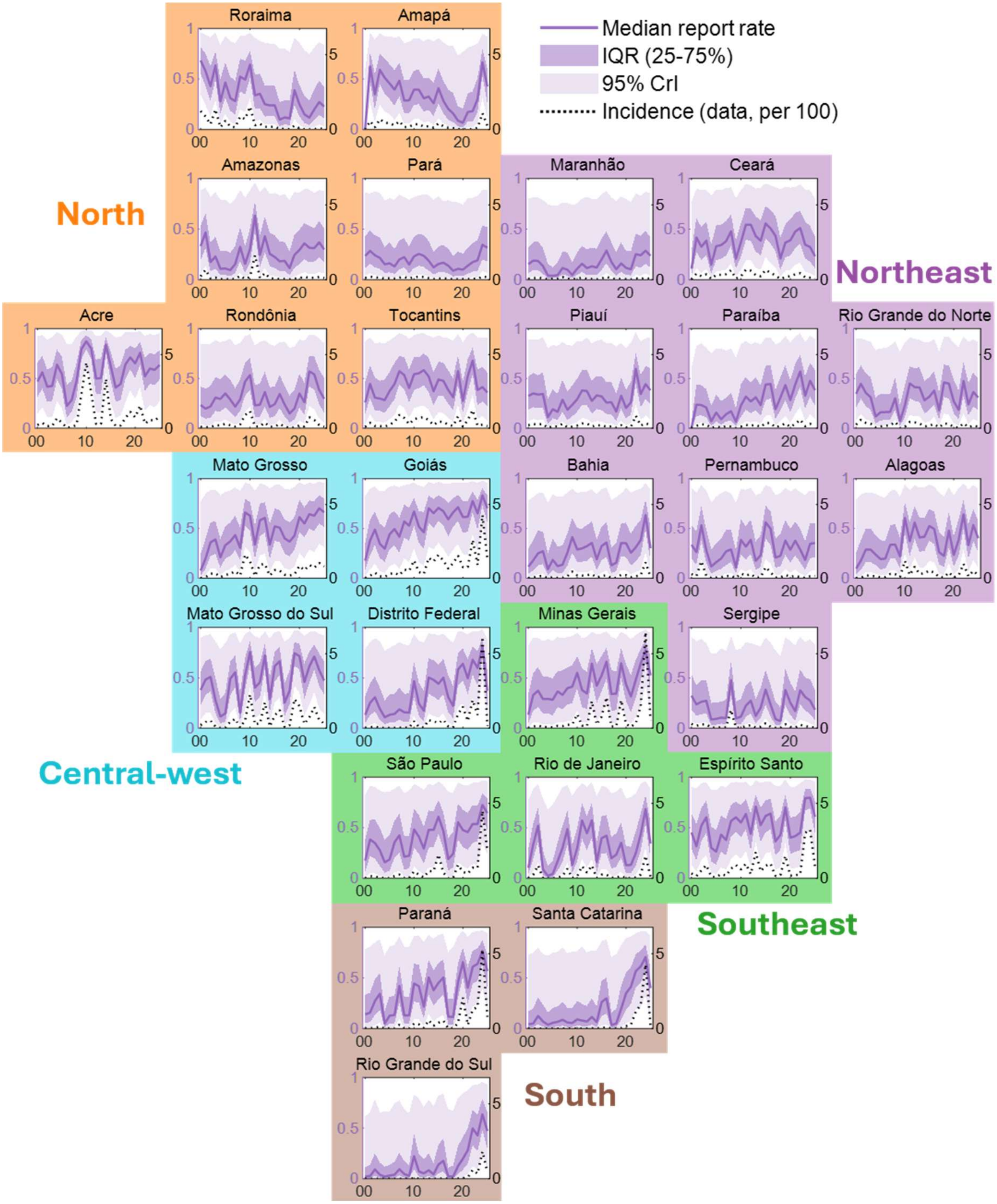


**Fig. S5**: Apparent illness-to-case report ratio by state and year. Dotted curves in each panel indicate the real-world incidence per 100 individuals.

**Table S5**: Apparent illness-to-case report ratio (%) by state and year (median and 95% CrI).

| Year | 00 | 01 | 02 | 03 | 04 | 05 | 06 | 07 | 08 | 09 | 10 | 11 | 12 | 13 | 14 | 15 | 16 | 17 | 18 | 19 | 20 | 21 | 22 | 23 | 24 | |
|---|---|---|---|---|---|---|---|---|---|---|---|---|---|---|---|---|---|---|---|---|---|---|---|---|---|---|
| RO | 23.37 [4, 84] | 19.61 [3, 85] | 21.91 [3, 85] | 30.77 [7, 88] | 29.40 [6, 86] | 34.98 [9, 88] | 28.81 [6, 90] | 21.74 [3, 83] | 28.90 [6, 86] | 46.92 [17, 92] | 53.39 [21, 94] | 22.96 [4, 83] | 23.67 [4, 87] | 36.31 [10, 88] | 21.41 [3, 84] | 24.18 [4, 82] | 34.36 [8, 86] | 23.84 [4, 87] | 14.85 [1, 81] | 19.77 [2, 82] | 34.85 [7, 89] | 26.13 [5, 88] | 57.16 [22, 94] | 54.14 [19, 94] | 38.33 [10, 90] | 29.54 [5, 86] |
| AC | 46.90 [14, 92] | 56.18 [21, 94] | 41.69 [12, 89] | 42.47 [11, 92] | 63.82 [27, 96] | 54.49 [20, 94] | 22.46 [3, 84] | 28.35 [6, 84] | 47.51 [13, 92] | 79.49 [48, 98] | 87.39 [61, 99] | 79.98 [48, 98] | 49.89 [15, 93] | 50.41 [16, 93] | 83.94 [55, 98] | 62.45 [25, 95] | 40.83 [10, 91] | 44.42 [12, 92] | 64.16 [26, 96] | 71.13 [38, 96] | 65.19 [30, 96] | 75.76 [41, 97] | 52.21 [18, 93] | 59.90 [23, 95] | 58.68 [21, 94] | 63.68 [27, 95] |
| AM | 33.10 [6, 88] | 46.58 [13, 91] | 17.33 [2, 82] | 22.84 [3, 86] | 10.70 [1, 79] | 11.05 [1, 79] | 9.39 [1, 75] | 15.39 [1, 79] | 32.37 [7, 89] | 15.56 [1, 81] | 30.43 [6, 88] | 63.28 [25, 96] | 24.53 [4, 87] | 42.89 [11, 92] | 25.46 [4, 85] | 22.40 [3, 86] | 18.05 [2, 83] | 18.59 [2, 84] | 11.30 [1, 79] | 24.55 [3, 87] | 29.38 [5, 85] | 35.86 [7, 90] | 31.46 [6, 89] | 31.15 [6, 86] | 36.91 [7, 89] | 29.77 [6, 88] |
| RR | 68.44 [35, 96] | 57.78 [24, 94] | 42.90 [13, 91] | 63.39 [29, 96] | 27.81 [6, 83] | 46.54 [15, 93] | 31.12 [7, 87] | 28.11 [5, 86] | 52.31 [20, 93] | 49.33 [17, 93] | 64.26 [31, 95] | 33.89 [8, 91] | 35.33 [10, 88] | 25.03 [5, 86] | 23.83 [5, 85] | 23.84 [5, 85] | 9.77 [1, 80] | 11.98 [1, 80] | 10.99 [1, 77] | 38.86 [10, 91] | 24.80 [4, 86] | 15.06 [2, 79] | 11.73 [1, 74] | 19.82 [3, 81] | 27.20 [5, 86] | 22.67 [3, 83] |
| PA | 23.44 [4, 86] | 28.44 [6, 85] | 24.08 [4, 87] | 21.51 [4, 86] | 15.14 [2, 78] | 16.14 [2, 80] | 13.04 [2, 79] | 19.93 [3, 84] | 22.23 [4, 82] | 14.85 [2, 79] | 23.03 [4, 82] | 25.51 [5, 83] | 23.55 [4, 84] | 18.66 [3, 84] | 12.16 [1, 79] | 15.44 [2, 82] | 16.74 [2, 78] | 13.85 [2, 78] | 8.80 [1, 77] | 10.66 [1, 76] | 9.87 [1, 74] | 13.75 [2, 81] | 17.30 [2, 81] | 19.07 [2, 83] | 34.53 [8, 88] | 31.53 [7, 89] |
| AP | 0.70 [0, 59] | 62.34 [26, 96] | 34.88 [8, 89] | 59.09 [25, 95] | 51.20 [18, 92] | 45.55 [15, 92] | 39.51 [10, 91] | 48.25 [16, 92] | 28.65 [6, 86] | 28.91 [6, 87] | 39.38 [10, 90] | 39.34 [11, 90] | 30.86 [7, 87] | 32.60 [8, 90] | 26.01 [4, 87] | 40.91 [11, 90] | 28.46 [6, 86] | 20.48 [3, 83] | 15.86 [2, 83] | 8.70 [1, 73] | 6.70 [0, 77] | 17.33 [2, 81] | 18.24 [2, 83] | 30.13 [6, 85] | 66.38 [31, 94] | 42.54 [12, 92] |
| TO | 25.85 [5, 87] | 44.42 [14, 91] | 30.71 [7, 85] | 29.84 [6, 86] | 28.26 [6, 85] | 37.36 [9, 88] | 44.49 [13, 92] | 57.31 [24, 95] | 53.44 [20, 94] | 40.39 [11, 90] | 54.31 [20, 93] | 55.22 [22, 95] | 58.38 [22, 95] | 51.99 [19, 93] | 37.15 [9, 91] | 48.88 [16, 93] | 47.61 [15, 91] | 38.50 [10, 91] | 31.13 [7, 88] | 57.37 [24, 95] | 27.42 [5, 86] | 52.50 [18, 93] | 67.86 [33, 96] | 38.37 [9, 91] | 41.33 [12, 91] | 35.81 [8, 90] |
| MA | 15.22 [2, 80] | 18.82 [3, 81] | 18.43 [3, 81] | 13.35 [2, 79] | 4.12 [0, 74] | 3.73 [1, 73] | 4.39 [1, 73] | 11.51 [2, 76] | 9.82 [1, 76] | 4.53 [0, 74] | 10.86 [1, 75] | 15.68 [3, 80] | 11.40 [1, 82] | 13.14 [1, 80] | 12.04 [1, 80] | 20.01 [3, 84] | 28.78 [6, 88] | 17.50 [3, 81] | 10.28 [1, 78] | 16.00 [2, 82] | 11.64 [1, 80] | 10.26 [1, 76] | 25.22 [5, 85] | 24.10 [4, 83] | 28.63 [6, 87] | 23.60 [4, 86] |
| PI | 32.38 [7, 89] | 34.91 [9, 89] | 33.83 [8, 89] | 34.37 [9, 91] | 11.15 [1, 80] | 19.08 [4, 83] | 16.47 [3, 83] | 28.69 [7, 86] | 18.59 [3, 83] | 23.55 [4, 83] | 29.14 [6, 87] | 37.94 [10, 87] | 37.42 [10, 91] | 27.49 [5, 85] | 32.97 [7, 88] | 31.97 [7, 89] | 25.78 [5, 85] | 26.14 [5, 85] | 16.69 [2, 82] | 32.62 [8, 86] | 20.38 [3, 80] | 26.04 [5, 86] | 59.69 [23, 96] | 34.30 [8, 87] | 44.29 [13, 90] | 38.09 [9, 91] |
| CE | 10.78 [1, 76] | 41.70 [12, 90] | 32.50 [7, 88] | 38.56 [11, 89] | 14.86 [2, 83] | 33.09 [8, 89] | 34.16 [9, 88] | 37.96 [10, 91] | 47.77 [16, 91] | 20.64 [3, 81] | 36.98 [9, 88] | 54.37 [19, 95] | 54.46 [19, 93] | 43.06 [12, 90] | 38.50 [10, 91] | 55.59 [22, 95] | 50.59 [16, 93] | 43.32 [12, 91] | 20.89 [3, 89] | 36.62 [9, 92] | 38.89 [9, 91] | 47.53 [14, 91] | 51.08 [17, 94] | 34.53 [8, 89] | 32.26 [6, 89] | 23.14 [3, 80] |
| RN | 34.62 [8, 90] | 44.58 [14, 90] | 35.52 [9, 88] | 31.04 [7, 87] | 11.87 [1, 78] | 15.99 [3, 80] | 16.25 [3, 80] | 16.52 [3, 83] | 30.08 [8, 87] | 7.30 [1, 77] | 23.04 [4, 86] | 45.02 [13, 91] | 41.28 [12, 90] | 40.96 [12, 89] | 28.15 [5, 86] | 31.69 [7, 87] | 37.83 [11, 89] | 18.09 [3, 77] | 46.03 [15, 92] | 44.84 [14, 92] | 27.01 [5, 86] | 18.52 [2, 81] | 47.43 [15, 92] | 23.96 [4, 87] | 36.39 [9, 89] | 30.85 [6, 86] |
| PB | 0.49 [0, 60] | 23.73 [4, 82] | 23.35 [5, 85] | 20.74 [5, 81] | 7.01 [0, 75] | 15.81 [2, 80] | 9.03 [1, 80] | 14.12 [3, 78] | 17.09 [3, 80] | 6.61 [1, 71] | 22.82 [4, 84] | 31.79 [7, 88] | 26.94 [6, 84] | 31.13 [7, 86] | 23.15 [4, 84] | 43.86 [12, 91] | 44.44 [13, 91] | 19.16 [3, 84] | 30.97 [7, 86] | 43.40 [12, 93] | 28.34 [5, 86] | 43.95 [14, 92] | 57.06 [21, 95] | 32.73 [7, 87] | 47.66 [14, 93] | 38.25 [9, 90] |
| PE | 33.68 [7, 91] | 26.27 [4, 86] | 53.25 [18, 94] | 28.06 [5, 88] | 11.91 [1, 77] | 16.22 [2, 82] | 19.87 [3, 86] | 27.06 [4, 88] | 29.06 [6, 85] | 11.05 [1, 76] | 38.66 [9, 90] | 31.85 [7, 89] | 38.72 [9, 91] | 17.65 [2, 83] | 23.30 [3, 85] | 56.24 [19, 93] | 49.89 [15, 92] | 20.81 [3, 84] | 22.70 [4, 85] | 37.95 [9, 91] | 28.71 [6, 87] | 37.82 [9, 90] | 28.94 [5, 86] | 18.46 [2, 84] | 34.64 [7, 88] | 35.49 [8, 87] |
| AL | 9.75 [1, 79] | 17.88 [2, 84] | 28.88 [6, 86] | 28.85 [6, 86] | 23.93 [4, 85] | 19.36 [3, 82] | 19.87 [3, 81] | 33.60 [7, 88] | 33.95 [8, 89] | 22.06 [4, 82] | 60.59 [25, 94] | 37.36 [9, 89] | 56.08 [21, 93] | 40.97 [11, 91] | 41.81 [11, 92] | 52.54 [18, 94] | 49.60 [16, 92] | 26.25 [5, 86] | 22.51 [3, 82] | 47.82 [15, 91] | 23.55 [4, 83] | 38.57 [8, 90] | 64.20 [29, 95] | 33.33 [7, 89] | 53.55 [18, 94] | 40.25 [11, 90] |
| SE | 32.42 [7, 90] | 23.83 [4, 86] | 26.42 [5, 88] | 27.42 [5, 84] | 8.86 [1, 74] | 9.49 [1, 74] | 10.71 [1, 79] | 10.27 [1, 77] | 47.76 [16, 91] | 9.91 [1, 78] | 11.69 [1, 79] | 22.54 [4, 85] | 27.59 [5, 84] | 12.39 [1, 83] | 20.53 [3, 81] | 38.40 [10, 90] | 23.69 [4, 85] | 9.28 [1, 77] | 7.17 [0, 72] | 38.02 [9, 90] | 25.43 [4, 87] | 17.18 [2, 82] | 37.78 [9, 91] | 31.13 [7, 86] | 27.62 [4, 88] | 18.48 [2, 83] |
| BA | 11.72 [1, 78] | 18.06 [2, 81] | 25.18 [4, 85] | 27.26 [5, 86] | 9.24 [1, 76] | 18.57 [2, 84] | 12.26 [1, 82] | 13.72 [1, 82] | 25.82 [4, 86] | 41.36 [10, 90] | 28.99 [6, 85] | 30.07 [6, 87] | 32.39 [6, 87] | 35.55 [8, 89] | 19.17 [2, 84] | 32.35 [6, 89] | 36.02 [8, 91] | 13.12 [1, 79] | 15.79 [2, 81] | 35.50 [8, 85] | 35.31 [8, 91] | 25.68 [4, 86] | 31.23 [6, 88] | 38.53 [8, 90] | 63.42 [26, 96] | 30.24 [5, 89] |
| MS | 38.05 [8, 90] | 48.31 [15, 92] | 51.07 [16, 93] | 28.00 [5, 86] | 11.87 [1, 78] | 14.79 [2, 84] | 49.10 [15, 93] | 55.56 [20, 95] | 15.45 [2, 84] | 52.71 [19, 94] | 75.64 [40, 97] | 40.52 [11, 91] | 45.72 [13, 90] | 72.20 [37, 96] | 29.14 [6, 89] | 61.63 [25, 95] | 66.54 [31, 96] | 25.62 [4, 86] | 37.27 [9, 89] | 74.48 [42, 97] | 71.63 [35, 97] | 45.96 [13, 91] | 61.37 [23, 95] | 71.20 [36, 97] | 57.76 [19, 94] | 47.39 [14, 92] |
| MT | 7.81 [0, 77] | 19.93 [3, 85] | 36.25 [10, 89] | 38.98 [10, 89] | 21.09 [3, 83] | 33.26 [7, 87] | 37.15 [9, 88] | 43.46 [13, 91] | 30.64 [7, 87] | 66.12 [31, 96] | 62.85 [26, 95] | 33.98 [7, 88] | 58.31 [23, 95] | 60.68 [25, 94] | 33.85 [7, 90] | 52.23 [17, 94] | 50.91 [17, 94] | 40.19 [10, 90] | 36.86 [8, 89] | 43.02 [12, 92] | 61.44 [25, 96] | 55.02 [20, 94] | 64.42 [29, 96] | 62.92 [26, 95] | 70.39 [35, 97] | 66.33 [31, 96] |
| GO | 17.77 [2, 83] | 38.11 [9, 90] | 48.95 [15, 92] | 34.62 [7, 87] | 29.80 [5, 88] | 44.25 [12, 92] | 49.52 [15, 93] | 41.87 [10, 91] | 57.15 [20, 94] | 52.89 [16, 94] | 67.11 [30, 96] | 51.91 [16, 93] | 46.23 [13, 92] | 70.44 [31, 97] | 65.43 [29, 95] | 70.78 [36, 97] | 67.40 [30, 95] | 59.22 [22, 95] | 66.19 [29, 96] | 71.47 [34, 96] | 61.59 [24, 96] | 61.78 [24, 96] | 77.63 [44, 98] | 65.27 [27, 96] | 83.28 [52, 98] | 70.38 [34, 96] |
| DF | 13.41 [1, 79] | 25.41 [3, 85] | 30.83 [5, 89] | 19.35 [2, 86] | 11.07 [1, 77] | 13.50 [1, 80] | 14.44 [1, 82] | 18.85 [2, 85] | 16.00 [1, 83] | 15.75 [1, 82] | 46.74 [12, 93] | 24.55 [3, 87] | 17.65 [2, 85] | 50.38 [14, 94] | 48.10 [12, 93] | 44.26 [11, 92] | 50.19 [15, 93] | 30.94 [5, 89] | 22.36 [2, 85] | 58.31 [21, 96] | 64.04 [25, 96] | 49.55 [14, 93] | 68.03 [30, 96] | 59.96 [20, 96] | 80.81 [48, 98] | 37.17 [8, 89] |
| MG | 13.51 [1, 82] | 33.15 [5, 90] | 37.32 [6, 90] | 29.09 [3, 86] | 29.22 [4, 88] | 28.45 [3, 86] | 36.61 [5, 90] | 33.27 [5, 89] | 39.82 [7, 91] | 41.40 [8, 91] | 55.10 [18, 95] | 38.08 [6, 89] | 34.48 [5, 90] | 64.11 [22, 96] | 42.31 [8, 93] | 52.39 [15, 94] | 66.16 [24, 96] | 34.00 [4, 90] | 34.52 [5, 89] | 64.48 [25, 96] | 43.63 [9, 93] | 31.69 [4, 88] | 48.34 [11, 92] | 64.26 [22, 96] | 77.47 [42, 98] | 52.48 [13, 94] |
| ES | 45.28 [14, 90] | 31.64 [6, 89] | 52.09 [18, 94] | 60.29 [23, 95] | 30.11 [5, 87] | 26.33 [4, 87] | 43.18 [11, 91] | 37.81 [9, 90] | 56.68 [19, 95] | 60.28 [24, 95] | 55.05 [19, 94] | 63.57 [27, 95] | 43.97 [12, 90] | 70.93 [35, 97] | 52.46 [15, 94] | 61.61 [25, 96] | 63.94 [26, 96] | 36.54 [8, 91] | 42.92 [11, 91] | 70.16 [34, 98] | 40.18 [9, 90] | 41.67 [10, 91] | 45.07 [11, 94] | 79.20 [45, 98] | 79.64 [47, 98] | 60.81 [23, 96] |
| RJ | 10.28 [1, 77] | 33.34 [7, 88] | 51.97 [16, 93] | 8.66 [0, 79] | 2.46 [0, 68] | 4.25 [0, 73] | 16.88 [2, 81] | 30.04 [5, 90] | 48.59 [15, 92] | 13.16 [1, 81] | 30.02 [5, 89] | 57.13 [19, 94] | 44.53 [11, 90] | 57.20 [19, 95] | 16.01 [1, 81] | 39.56 [9, 91] | 42.21 [9, 91] | 19.80 [2, 84] | 23.33 [3, 85] | 35.15 [6, 90] | 12.94 [1, 79] | 13.14 [1, 80] | 24.91 [4, 84] | 43.50 [11, 94] | 68.45 [30, 96] | 34.25 [7, 89] |
| SP | 17.48 [1, 83] | 38.37 [5, 91] | 34.91 [5, 90] | 27.67 [2, 87] | 15.72 [1, 85] | 18.62 [1, 86] | 35.59 [5, 88] | 42.51 [7, 91] | 21.34 [1, 84] | 22.31 [1, 86] | 48.29 [11, 93] | 41.15 [7, 91] | 29.29 [3, 87] | 47.96 [11, 93] | 47.85 [11, 92] | 60.77 [21, 96] | 46.81 [10, 92] | 18.98 [1, 85] | 24.19 [2, 85] | 55.44 [16, 95] | 47.68 [12, 92] | 45.64 [9, 92] | 53.75 [14, 94] | 53.72 [14, 94] | 72.52 [35, 97] | 64.46 [24, 96] |
| PR | 13.88 [1, 78] | 15.70 [1, 81] | 27.20 [3, 85] | 34.34 [5, 88] | 4.84 [0, 72] | 12.64 [1, 81] | 13.35 [1, 82] | 42.07 [9, 91] | 12.88 [1, 81] | 13.17 [1, 81] | 44.45 [9, 91] | 41.79 [9, 92] | 21.61 [2, 85] | 50.42 [14, 92] | 37.83 [7, 89] | 45.14 [11, 92] | 50.41 [13, 93] | 11.77 [1, 75] | 11.33 [1, 81] | 45.86 [11, 91] | 65.61 [27, 97] | 40.41 [8, 91] | 60.95 [22, 95] | 63.94 [23, 96] | 76.62 [39, 97] | 56.74 [19, 94] |
| SC | 4.21 [0, 73] | 4.83 [0, 75] | 12.43 [0, 80] | 6.95 [0, 73] | 3.70 [0, 72] | 5.60 [0, 75] | 6.52 [0, 76] | 8.62 [0, 80] | 5.93 [0, 74] | 5.40 [0, 76] | 8.93 [0, 76] | 7.84 [0, 81] | 6.90 [0, 75] | 11.41 [0, 77] | 6.31 [0, 77] | 26.01 [2, 86] | 29.93 [4, 88] | 3.38 [0, 70] | 4.96 [0, 73] | 19.49 [1, 82] | 34.83 [5, 89] | 41.30 [8, 91] | 56.75 [18, 94] | 61.36 [21, 96] | 70.59 [32, 97] | 39.92 [8, 92] |
| RS | 2.20 [0, 61] | 2.58 [0, 66] | 9.17 [0, 78] | 3.93 [0, 66] | 2.05 [0, 66] | 3.12 [0, 66] | 3.92 [0, 74] | 9.36 [0, 77] | 4.65 [0, 69] | 2.75 [0, 65] | 22.23 [2, 83] | 7.13 [0, 79] | 5.29 [0, 71] | 8.34 [0, 78] | 4.29 [0, 72] | 14.02 [1, 82] | 17.73 [1, 84] | 2.11 [0, 69] | 2.18 [0, 67] | 12.47 [1, 80] | 19.76 [2, 85] | 28.09 [4, 87] | 49.75 [12, 92] | 42.85 [9, 93] | 63.63 [24, 96] | 47.14 [11, 93] |

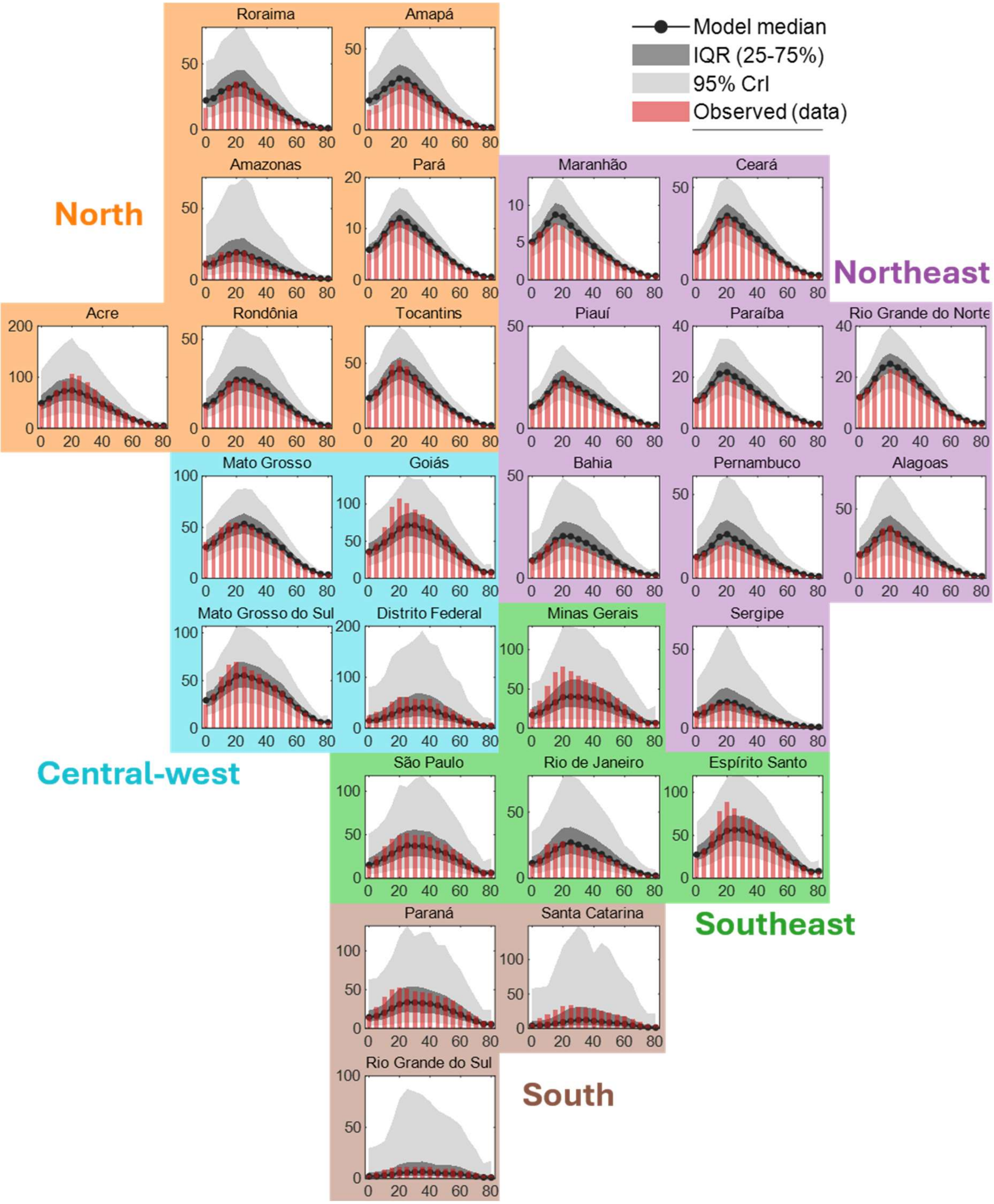


**Fig. S6**: Model-fitted case incidence across states in Brazil using the catalytic modeling framework. The curves represent the model medians, while the light grey shaded areas denote the 95% credible intervals, and the dark grey shaded areas indicate the IQRs. The red bars represent the observed incidence.

We also examined how the ratio of reported cases to infections varies across states from the model simulation. **Fig. S7** displays these state-specific ratios, showing substantial heterogeneity. The highest ratio was observed in Acre, with a median of 0.09 with CrI [0.06, 0.12], whereas the lowest was in Maranhão with a median 0.01 and narrow estimation interval. This means, there would be 100 actual infections if single case is reported. These results are summarized in **Table S6**.

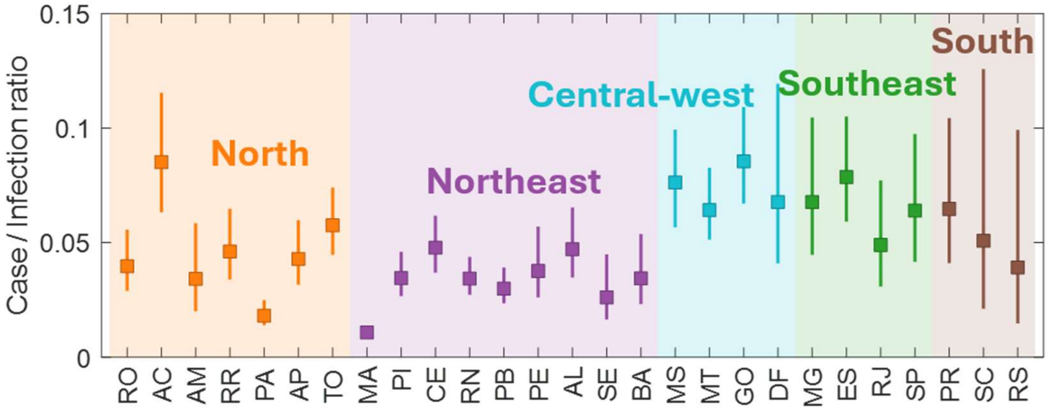


**Fig. S7**: Case/Infection across states in Brazil. Color of box chart indicates the geographical region.

**Table S6**: Case/Infection ratio across states in Brazil.

| State | Median, 95% CrI |
|---|---|
| **Rondônia (RO)** | 0.04 [0.03, 0.06] |
| **Acre (AC)** | 0.09 [0.06, 0.12] |
| **Amazonas (AM)** | 0.03 [0.02, 0.06] |
| **Roraima (RR)** | 0.05 [0.03, 0.06] |
| **Pará (PA)** | 0.02 [0.01, 0.02] |
| **Amapá (AP)** | 0.04 [0.03, 0.06] |
| **Tocantins (TO)** | 0.06 [0.04, 0.07] |
| **Maranhão (MA)** | 0.01 [0.01, 0.01] |
| **Piauí (PI)** | 0.03 [0.03, 0.05] |
| **Ceará (CE)** | 0.05 [0.04, 0.06] |
| **Rio Grande do Norte (RN)** | 0.03 [0.03, 0.04] |
| **Paraíba (PB)** | 0.03 [0.02, 0.04] |
| **Pernambuco (PE)** | 0.04 [0.03, 0.06] |
| **Alagoas (AL)** | 0.05 [0.03, 0.07] |
| **Sergipe (SE)** | 0.03 [0.02, 0.04] |
| **Bahia (BA)** | 0.03 [0.02, 0.05] |
| **Mato Grosso do Sul (MS)** | 0.08 [0.06, 0.10] |
| **Mato Grosso (MT)** | 0.06 [0.05, 0.08] |
| **Goiás (GO)** | 0.09 [0.07, 0.11] |
| **Distrito Federal (DF)** | 0.07 [0.04, 0.12] |
| **Minas Gerais (MG)** | 0.07 [0.04, 0.10] |
| **Espirito Santo (ES)** | 0.08 [0.06, 0.11] |
| **Rio de Janeiro (RJ)** | 0.05 [0.03, 0.08] |
| **São Paulo (SP)** | 0.06 [0.04, 0.10] |
| **Paraná (PR)** | 0.06 [0.04, 0.10] |
| **Santa Catarina (SC)** | 0.05 [0.02, 0.13] |
| **Rio Grande do Sul (RS)** | 0.04 [0.01, 0.10] |

#### S3-4. Immune profile and comparison with real-world surveillances

**Table S7** reports the model-derived immune profile for each state. Values are corrected for total population size and show the change between 2000 and 2026. As of 2000, the state with the lowest estimated prevalence was Acre (median 5.43%, 95% CrI [0.21, 17.30]), and the state with the highest was Rio de Janeiro (median 55.48%, 95% CrI [27.93, 66.36]). By 2026—profiled using data through 2025—prevalence was generally high across nearly all states (lowest region Rio Grande do Sul, median 69.85%, 95% CrI [50.37, 86.57]; highest region Goiás, median 90.58%, 95% CrI [85.77, 94.51]). **Fig. S8** shows, using color varying by year, the age-specific proportion of individuals with one or fewer prior infections (i.e., those still susceptible to disease), illustrating how the population's immune profile changed from 2000 to 2026.

**Table S7**: Immune profiles showing the proportion of the population exposed to DENV (%) in 2000 and 2026 across 27 Brazilian states, with median values and 95% credible intervals.

| **Year** | **2000** | | | | | **2026** | | | | |
|---|---|---|---|---|---|---|---|---|---|---|
| **Exposure** | **0** | **1** | **2** | **3** | **4** | **0** | **1** | **2** | **3** | **4** |
| **RO** | 89.72<br>[73.12, 99.49] | 9.61<br>[0.51, 21.73] | 0.66<br>[0.00, 4.54] | 0.03<br>[0.00, 0.55] | 0.00<br>[0.00, 0.03] | 19.31<br>[12.30, 27.29] | 26.00<br>[18.87, 32.77] | 29.75<br>[26.78, 31.59] | 19.48<br>[11.60, 27.54] | 5.11<br>[1.97, 11.12] |
| **AC** | 94.57<br>[82.70, 99.79] | 5.23<br>[0.21, 15.15] | 0.20<br>[0.00, 2.01] | 0.00<br>[0.00, 0.16] | 0.00<br>[0.00, 0.01] | 14.25<br>[8.94, 20.06] | 20.84<br>[14.69, 26.63] | 29.24<br>[25.28, 31.32] | 25.87<br>[19.19, 32.12] | 9.63<br>[5.02, 17.68] |
| **AM** | 78.09<br>[57.87, 99.00] | 18.30<br>[0.99, 27.42] | 3.21<br>[0.01, 11.21] | 0.33<br>[0.00, 3.07] | 0.02<br>[0.00, 0.39] | 26.59<br>[16.80, 39.01] | 28.39<br>[20.90, 34.64] | 26.48<br>[20.30, 29.75] | 14.87<br>[6.40, 23.97] | 3.41<br>[0.78, 9.07] |
| **RR** | 84.40<br>[72.61, 97.46] | 13.89<br>[2.50, 21.72] | 1.62<br>[0.04, 4.88] | 0.11<br>[0.00, 0.64] | 0.00<br>[0.00, 0.04] | 22.48<br>[16.60, 28.74] | 20.62<br>[15.75, 26.00] | 26.83<br>[24.27, 28.60] | 22.30<br>[15.78, 27.73] | 7.63<br>[3.84, 13.52] |
| **PA** | 70.38<br>[61.18, 84.47] | 22.85<br>[13.80, 26.78] | 5.76<br>[1.61, 9.67] | 0.88<br>[0.11, 2.22] | 0.06<br>[0.00, 0.23] | 25.04<br>[17.25, 33.88] | 28.06<br>[22.39, 33.14] | 27.56<br>[23.33, 30.19] | 15.54<br>[9.05, 22.77] | 3.60<br>[1.42, 7.50] |
| **AP** | 91.21<br>[81.21, 98.68] | 8.24<br>[1.31, 16.21] | 0.53<br>[0.01, 2.37] | 0.02<br>[0.00, 0.21] | 0.00<br>[0.00, 0.01] | 22.51<br>[14.48, 31.60] | 27.65<br>[21.19, 33.06] | 28.33<br>[24.40, 30.90] | 17.03<br>[10.20, 25.19] | 4.16<br>[1.68, 9.11] |
| **TO** | 59.68<br>[52.39, 68.92] | 27.26<br>[23.66, 29.27] | 10.31<br>[6.23, 13.75] | 2.45<br>[0.98, 4.33] | 0.27<br>[0.07, 0.64] | 15.77<br>[10.67, 21.49] | 21.50<br>[15.56, 26.99] | 29.24<br>[26.17, 31.08] | 24.55<br>[17.88, 30.78] | 8.67<br>[4.58, 15.71] |
| **MA** | 64.85<br>[57.15, 74.41] | 25.25<br>[20.56, 27.91] | 8.10<br>[4.41, 11.59] | 1.64<br>[0.57, 3.18] | 0.15<br>[0.03, 0.41] | 20.83<br>[14.24, 26.87] | 20.68<br>[16.46, 25.06] | 26.59<br>[24.04, 28.84] | 23.12<br>[17.80, 28.41] | 8.65<br>[4.99, 14.25] |
| **PI** | 60.10<br>[52.50, 69.97] | 27.27<br>[23.21, 29.49] | 10.01<br>[5.84, 13.62] | 2.29<br>[0.88, 4.23] | 0.24<br>[0.06, 0.61] | 18.70<br>[12.44, 25.69] | 25.14<br>[19.08, 30.73] | 29.38<br>[26.98, 31.25] | 20.51<br>[13.72, 27.68] | 5.92<br>[2.82, 11.35] |
| **CE** | 57.05<br>[48.31, 69.07] | 28.00<br>[23.76, 29.92] | 11.54<br>[6.22, 15.73] | 2.99<br>[0.99, 5.68] | 0.35<br>[0.07, 0.98] | 17.74<br>[11.91, 24.05] | 22.73<br>[16.52, 28.30] | 29.13<br>[26.64, 30.94] | 22.73<br>[16.09, 29.57] | 7.34<br>[3.72, 13.96] |
| **RN** | 56.26<br>[48.95, 65.49] | 28.57<br>[25.54, 30.36] | 11.72<br>[7.61, 15.34] | 2.99<br>[1.37, 5.14] | 0.35<br>[0.11, 0.82] | 16.36<br>[11.44, 21.87] | 20.43<br>[15.45, 25.86] | 28.47<br>[25.54, 30.47] | 25.12<br>[19.00, 30.61] | 9.33<br>[5.07, 16.00] |
| **PB** | 52.43<br>[45.48, 60.87] | 29.05<br>[27.02, 30.58] | 13.71<br>[9.70, 17.06] | 4.16<br>[2.11, 6.74] | 0.58<br>[0.20, 1.27] | 15.43<br>[10.57, 20.83] | 20.20<br>[15.19, 25.57] | 28.43<br>[25.21, 30.54] | 25.73<br>[19.69, 30.87] | 10.00<br>[5.55, 16.74] |
| **PE** | 48.97<br>[40.33, 61.20] | 29.33<br>[27.21, 30.79] | 15.41<br>[9.45, 19.40] | 5.25<br>[2.04, 9.13] | 0.84<br>[0.19, 2.13] | 17.42<br>[11.54, 24.31] | 20.82<br>[14.73, 27.11] | 27.73<br>[24.11, 29.97] | 24.39<br>[17.03, 30.60] | 9.21<br>[4.45, 17.42] |
| **AL** | 54.28<br>[46.97, 65.05] | 28.25<br>[25.29, 29.91] | 12.90<br>[7.86, 16.42] | 3.81<br>[1.48, 6.52] | 0.53<br>[0.12, 1.28] | 15.89<br>[10.18, 21.98] | 21.75<br>[15.55, 27.92] | 29.07<br>[25.82, 31.02] | 24.27<br>[17.41, 31.00] | 8.64<br>[4.37, 16.42] |
| **SE** | 51.43<br>[42.87, 61.58] | 28.89<br>[26.72, 30.47] | 14.21<br>[9.41, 18.20] | 4.56<br>[1.99, 8.04] | 0.69<br>[0.19, 1.78] | 18.19<br>[12.36, 25.05] | 20.26<br>[14.52, 26.60] | 27.16<br>[23.59, 29.51] | 24.55<br>[17.07, 30.44] | 9.60<br>[4.49, 17.66] |
| **BA** | 57.12<br>[45.40, 91.35] | 28.21<br>[8.18, 30.41] | 11.22<br>[0.45, 17.06] | 2.78<br>[0.01, 6.68] | 0.32<br>[0.00, 1.27] | 17.42<br>[10.73, 26.79] | 25.12<br>[17.17, 33.61] | 29.77<br>[26.01, 31.85] | 21.04<br>[11.23, 29.74] | 6.12<br>[1.92, 13.85] |
| **MS** | 82.00<br>[61.87, 99.21] | 15.83<br>[0.79, 27.53] | 1.99<br>[0.00, 8.91] | 0.14<br>[0.00, 1.72] | 0.00<br>[0.00, 0.15] | 13.75<br>[8.78, 19.68] | 22.10<br>[15.03, 28.49] | 30.71<br>[26.85, 32.37] | 25.00<br>[17.44, 31.93] | 8.21<br>[3.97, 16.25] |
| **MT** | 70.96<br>[56.83, 97.66] | 23.12<br>[2.30, 29.03] | 5.23<br>[0.03, 11.26] | 0.69<br>[0.00, 2.68] | 0.04<br>[0.00, 0.28] | 11.98<br>[7.16, 17.21] | 19.11<br>[13.32, 25.80] | 29.32<br>[24.49, 31.78] | 28.17<br>[20.94, 33.52] | 11.41<br>[5.73, 20.16] |
| **GO** | 94.53<br>[73.44, 99.75] | 5.29<br>[0.25, 21.72] | 0.18<br>[0.00, 4.34] | 0.00<br>[0.00, 0.49] | 0.00<br>[0.00, 0.02] | 9.42<br>[5.49, 14.23] | 18.90<br>[12.22, 25.70] | 30.83<br>[25.05, 33.22] | 29.36<br>[22.05, 35.27] | 11.46<br>[5.99, 21.33] |
| **DF** | 92.30<br>[67.44, 99.78] | 7.33<br>[0.22, 25.19] | 0.35<br>[0.00, 6.46] | 0.01<br>[0.00, 0.95] | 0.00<br>[0.00, 0.06] | 12.91<br>[7.67, 20.48] | 26.03<br>[17.28, 34.07] | 32.81<br>[29.62, 34.26] | 21.96<br>[13.13, 31.09] | 5.71<br>[2.23, 12.77] |
| **MG** | 88.68<br>[59.10, 99.52] | 10.51<br>[0.48, 28.52] | 0.77<br>[0.00, 10.04] | 0.03<br>[0.00, 2.11] | 0.00<br>[0.00, 0.19] | 12.14<br>[7.28, 19.29] | 23.25<br>[15.41, 31.65] | 32.06<br>[28.44, 33.70] | 24.64<br>[15.54, 32.56] | 7.50<br>[3.06, 15.44] |
| **ES** | 84.01<br>[58.93, 99.34] | 14.38<br>[0.66, 28.75] | 1.56<br>[0.00, 10.19] | 0.09<br>[0.00, 2.15] | 0.00<br>[0.00, 0.20] | 11.29<br>[6.81, 17.26] | 21.23<br>[14.13, 28.44] | 31.30<br>[26.65, 33.14] | 26.71<br>[18.54, 33.65] | 9.38<br>[4.39, 17.91] |
| **RJ** | 44.52<br>[33.64, 72.07] | 30.50<br>[22.84, 31.92] | 17.45<br>[4.59, 22.52] | 6.05<br>[0.50, 12.13] | 0.94<br>[0.02, 3.18] | 15.72<br>[9.80, 23.37] | 22.09<br>[14.95, 30.50] | 29.21<br>[25.14, 31.43] | 24.16<br>[14.93, 31.46] | 8.43<br>[3.15, 17.33] |
| **SP** | 89.26<br>[60.61, 99.63] | 10.05<br>[0.37, 28.39] | 0.67<br>[0.00, 9.13] | 0.02<br>[0.00, 1.70] | 0.00<br>[0.00, 0.14] | 13.72<br>[7.48, 21.95] | 25.82<br>[16.32, 33.96] | 32.07<br>[28.39, 33.79] | 21.90<br>[12.80, 31.75] | 5.88<br>[2.18, 14.24] |
| **PR** | 90.95<br>[58.92, 99.66] | 8.55<br>[0.34, 28.60] | 0.49<br>[0.00, 10.22] | 0.01<br>[0.00, 2.19] | 0.00<br>[0.00, 0.21] | 14.66<br>[8.04, 23.73] | 27.77<br>[17.79, 35.63] | 32.22<br>[27.83, 33.97] | 19.88<br>[11.04, 30.65] | 4.81<br>[1.70, 12.57] |
| **SC** | 89.74<br>[34.15, 99.77] | 9.61<br>[0.23, 30.98] | 0.63<br>[0.00, 22.02] | 0.02<br>[0.00, 12.33] | 0.00<br>[0.00, 3.50] | 24.19<br>[11.95, 40.09] | 36.45<br>[23.02, 39.98] | 27.40<br>[16.97, 33.08] | 10.32<br>[3.50, 24.84] | 1.52<br>[0.28, 8.92] |
| **RS** | 85.54<br>[30.98, 99.65] | 13.21<br>[0.35, 31.29] | 1.20<br>[0.00, 23.24] | 0.06<br>[0.00, 14.39] | 0.00<br>[0.00, 4.48] | 30.15<br>[13.43, 49.63] | 37.73<br>[24.48, 40.23] | 23.50<br>[11.80, 31.53] | 7.14<br>[1.77, 23.01] | 0.85<br>[0.10, 8.21] |

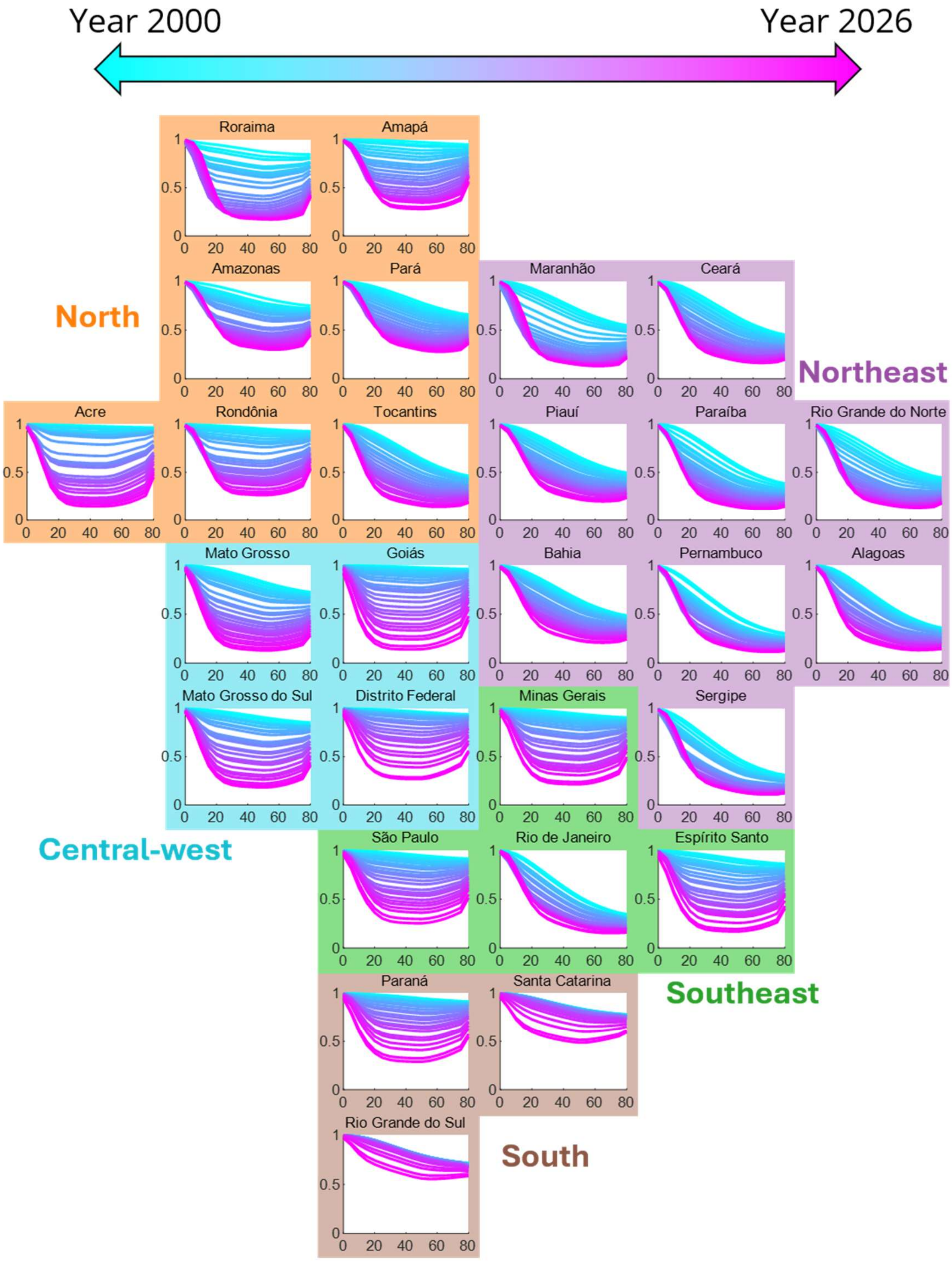


**Fig. S8**: Temporal evolution of the age-specific proportion of individuals with fewer than two prior dengue infections (corresponding to populations with a higher likelihood of progressing to reported dengue cases) between 2000 and 2026. Each panel represents a state-specific trajectory. Curves are colored according to calendar year, with earlier years shown in cyan and later years in magenta.

In this study, we aimed to infer the immune profile using only case surveillance data. To assess the realism of our estimates, we compiled and aggregated real-world serosurveys by state and compared their reported ranges with our model-derived estimates [1–22]. **Fig. S9** displays the simulation outputs (dark curve: simulation median; gray area: 95% CrI; green curve/area: real-world serosurvey) alongside the corresponding serosurvey values for each state, grouped by geographical region and plotted by survey year. Note that the title of each panel indicates the state name and survey year.

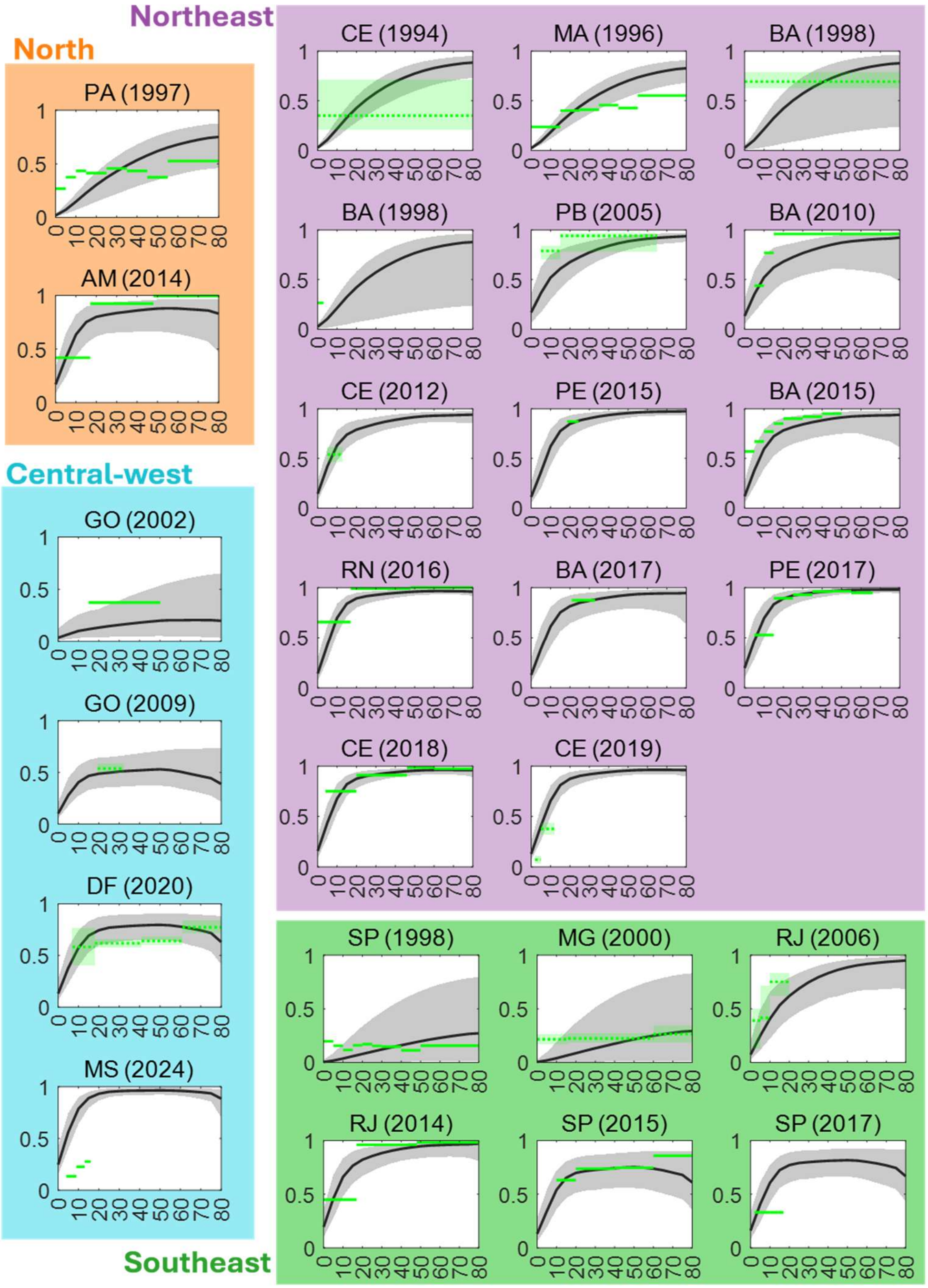


**Fig. S9**: Comparison of model-based immune profile estimates with real-world serosurvey data across Brazilian states. Dark curves denote the simulation medians, and shaded areas indicate the 95% credible intervals. Green curves and shaded regions represent serosurvey measurements. Panels are grouped by geographical region, with each panel title indicating the corresponding state and survey year. The x-axis represents age, and the y-axis represents seroprevalence.

**S4. Supplementary figures and tables for the SIR-type model framework**

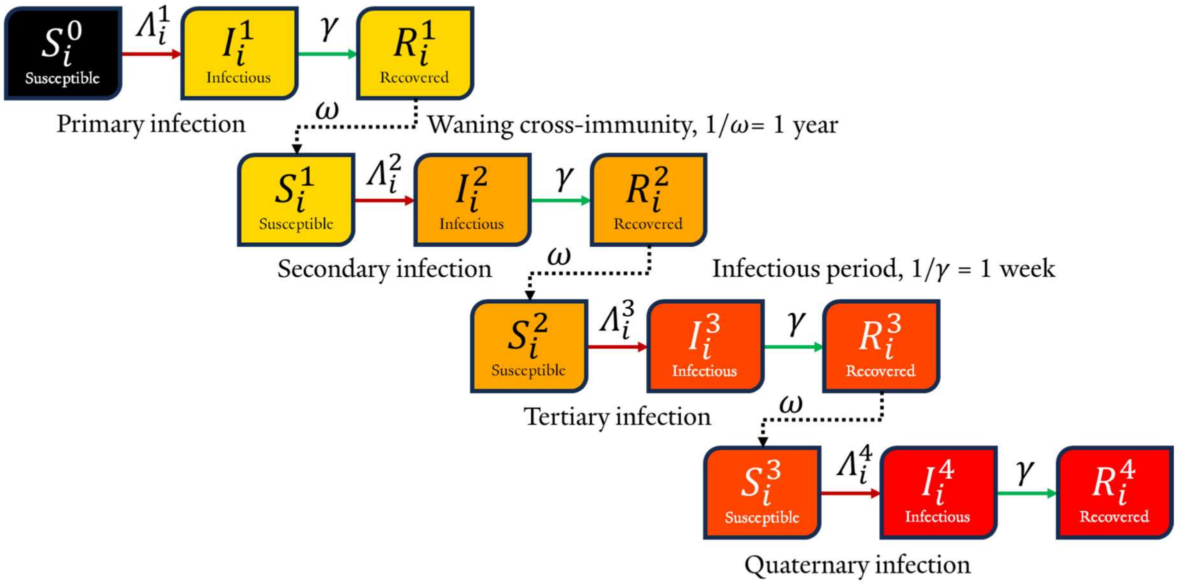


**Fig. S10**: Flow diagram of the SIR-type mathematical model. $S_{a,s}^k$, $I_{a,s}^k$, and $R_{a,s}^k$ are the number of susceptible, infectious, and recovered individuals with $k$ prior dengue infections in age group $a$ in state $s$, respectively. Detailed description of the model is in the main text (method section).

**Table S8**: SIR-type model parameters.

| Symbol | Description | Value | Reference |
|---|---|---|---|
| $\alpha$ | Recovery rate | 52 (1-week infectious period) | [23] |
| $\omega$ | Waning rate of immunity induced by natural infection | 1 (1-year) | [24] |
| $surv_a$ | Probability that individuals in age group '$a$' survive and proceed into the next age group '$a+1$' | 0.9851 (0–4y)<br>0.9987 (5–9y)<br>0.9984 (10–14y)<br>0.9948 (15–19y)<br>0.9919 (20–24y)<br>0.9911 (25–29y)<br>0.9905 (30–34y)<br>0.9885 (35–39y)<br>0.9847 (40–44y)<br>0.9792 (45–49y)<br>0.9700 (50–54y)<br>0.9567 (55–59y)<br>0.9364 (60–64y)<br>0.9053 (65–69y)<br>0.8673 (70–74y)<br>0.7911 (75–79y)<br>0.6733 (≥80y) | [25] |
| $\mu_n$ | Natural birth rate (per 1K) | 12.65 | [26] |
| $\mu$ | Aging rate between 5-year age intervals | 0.2 (5 years transition time) | |
| $\mu_f$ | Natural death rate in the final age group | 0.11 (8.9 years life expectancy) | [25] |

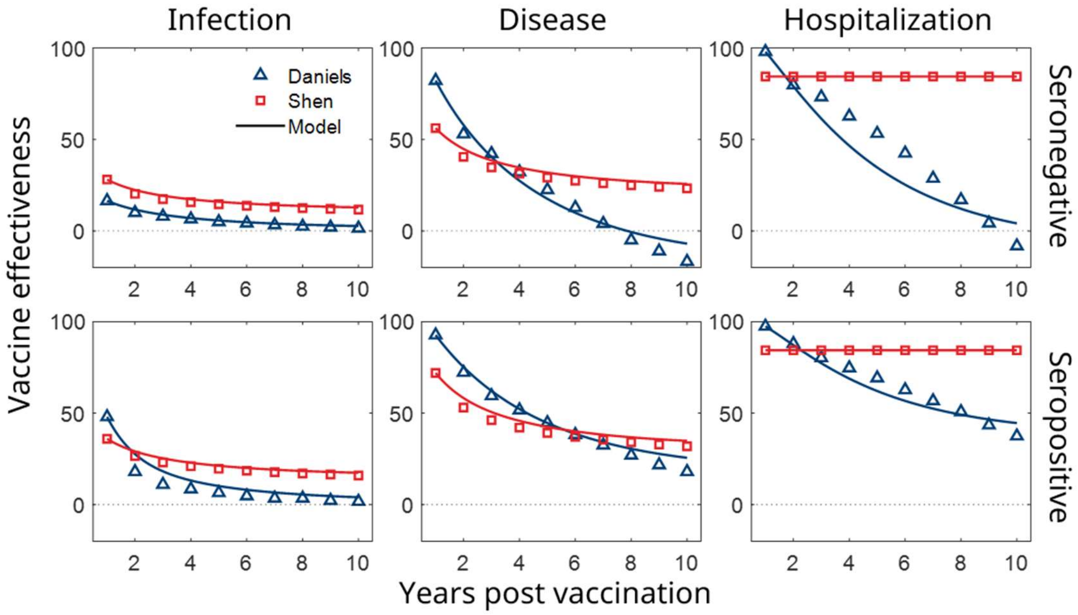


**Fig. S11:** Aggregated vaccine effectiveness (markers) and waning dynamics from the model (solid curve).

**Table S9**: Vaccine effectiveness setting from two studies [27,28].

| **Study** | **Shen** | | | | | | **Daniels** | | | | | |
|---|---|---|---|---|---|---|---|---|---|---|---|---|
| **Compartment** | **< 1 year (–)** | | **< 5 years (·)** | | **> 5 years (··)** | | **< 1 year (–)** | | **< 5 years (·)** | | **> 5 years (··)** | |
| **Serostatus** | **neg** | **pos** | **neg** | **pos** | **neg** | **pos** | **neg** | **pos** | **neg** | **pos** | **neg** | **pos** |
| **Infection** | 0.28 | 0.36 | 0.20 | 0.27 | 0.12 | 0.16 | 0.17 | 0.48 | 0.10 | 0.18 | 0.02 | 0.02 |
| **Disease** | 0.56 | 0.72 | 0.41 | 0.53 | 0.23 | 0.32 | 0.82 | 0.93 | 0.53 | 0.72 | -0.17 | 0.18 |
| **Hospitalization** | 0.84 | 0.84 | 0.84 | 0.84 | 0.84 | 0.84 | 0.98 | 0.98 | 0.80 | 0.88 | -0.08 | 0.38 |

**Table S10**: The quantity of vaccine distributed over the 2024/25 period in Brazil.

| State | Distributed doses | Adjusted, per 1000 target population |
|---|---|---|
| **Rondônia (RO)** | 16,526 | 61.37 |
| **Acre (AC)** | 41,432 | 252.2 |
| **Amazonas (AM)** | 163,096 | 198.48 |
| **Roraima (RR)** | 32,669 | 276.17 |
| **Pará (PA)** | 102,757 | 72.28 |
| **Amapá (AP)** | 22,893 | 138.18 |
| **Tocantins (TO)** | 31,300 | 128.19 |
| **Maranhão (MA)** | 82,731 | 74.4 |
| **Piauí (PI)** | 51,050 | 113.92 |
| **Ceará (CE)** | 58,115 | 45.71 |
| **Rio Grande do Norte (RN)** | 104,670 | 219.73 |
| **Paraíba (PB)** | 79,452 | 146.86 |
| **Pernambuco (PE)** | 112,562 | 82.1 |
| **Alagoas (AL)** | 45,836 | 93.36 |
| **Sergipe (SE)** | 22,446 | 66.69 |
| **Bahia (BA)** | 418,873 | 211.11 |
| **Mato Grosso do Sul (MS)** | 141,386 | 325.99 |
| **Mato Grosso (MT)** | 89,232 | 164.66 |
| **Goiás (GO)** | 325,051 | 305.92 |
| **Distrito Federal (DF)** | 150,609 | 401.2 |
| **Minas Gerais (MG)** | 433,816 | 167.91 |
| **Espirito Santo (ES)** | 166,358 | 290.79 |
| **Rio de Janeiro (RJ)** | 531,040 | 241.23 |
| **São Paulo (SP)** | 1,239,319 | 204.56 |
| **Paraná (PR)** | 275,071 | 179.69 |
| **Santa Catarina (SC)** | 160,096 | 168.09 |
| **Rio Grande do Sul (RS)** | 101,615 | 74.88 |

**Table S11**: Years of life lost (YLL) and case hospitalization/fatality ratio by age group.

| Age group | Years of life lost (YLL, year) | Case hospitalization ratio | Case fatality ratio |
|---|---|---|---|
| **0-4y** | 76.4 | 0.048226 | 0.001369 |
| **5-9y** | 72.5 | 0.042775 | 0.00066 |
| **10-14y** | 67.6 | 0.034394 | 0.000516 |
| **15-19y** | 62.7 | 0.025588 | 0.000582 |
| **20-24y** | 58 | 0.02291 | 0.000538 |
| **25-29y** | 53.4 | 0.02366 | 0.000611 |
| **30-34y** | 48.8 | 0.024754 | 0.000707 |
| **35-39y** | 44.2 | 0.025738 | 0.000858 |
| **40-44y** | 39.6 | 0.026329 | 0.001023 |
| **45-49y** | 35.1 | 0.027372 | 0.00119 |
| **50-54y** | 30.7 | 0.029407 | 0.001453 |
| **55-59y** | 26.5 | 0.032317 | 0.001825 |
| **60-64y** | 22.5 | 0.036942 | 0.002244 |
| **65-69y** | 18.7 | 0.044077 | 0.003219 |
| **70-74y** | 15.1 | 0.055893 | 0.004579 |
| **75-79y** | 11.8 | 0.072145 | 0.007225 |
| **≥80y** | 8.9 | 0.110941 | 0.01683 |

**S5. Supplementary SIR-type model outputs**

**S5-1. Projection of BoD without vaccination**

We first used the SIR model without vaccination to estimate the burden of disease (BoD) expected over the next five years. **Table S12** provides the projected annual BoD across states, showing an increasing trend from the northern to the southern states. **Fig. S12** further illustrates the age-group composition of BoD by geographical region. States in the South exhibited a larger contribution from older age groups, whereas states in the Northeast and Southeast showed a higher contribution from younger age groups.

**Table S12**: Projected annual burden of disease (BoD) per 100,000 population by state without vaccine administration.

| State \ BoD | Case | Hospitalization | Fatality | DALY |
|---|---|---|---|---|
| Rondônia (RO) | 3883.86 [3775.29, 4076.87] | 135.73 [131.05, 143.83] | 5.32 [5.13, 5.62] | 208.82 [200.32, 223.43] |
| Acre (AC) | 3271.88 [3180.28, 3374.47] | 121.37 [117.28, 125.96] | 4.36 [4.22, 4.52] | 193.88 [186.03, 202.72] |
| Amazonas (AM) | 4375.05 [4229.40, 4575.42] | 148.39 [142.37, 156.70] | 5.11 [4.90, 5.39] | 227.12 [216.43, 242.11] |
| Roraima (RR) | 2910.20 [2885.70, 3364.71] | 97.30 [95.93, 115.27] | 3.16 [3.09, 3.71] | 147.66 [144.84, 178.27] |
| Pará (PA) | 4208.81 [4083.75, 4500.03] | 140.84 [135.66, 152.71] | 5.10 [4.91, 5.51] | 214.34 [205.19, 235.47] |
| Amapá (AP) | 4104.34 [3956.45, 4336.83] | 140.70 [134.53, 150.40] | 4.98 [4.76, 5.32] | 216.09 [205.09, 233.67] |
| Tocantins (TO) | 3364.82 [3221.87, 3488.51] | 118.97 [112.95, 124.36] | 4.21 [4.01, 4.40] | 186.94 [175.98, 197.19] |
| Maranhão (MA) | 3147.09 [2969.69, 3357.64] | 107.84 [100.81, 116.56] | 3.71 [3.47, 4.01] | 164.35 [152.56, 179.62] |
| Piauí (PI) | 3730.87 [3632.23, 3909.03] | 128.70 [124.46, 136.11] | 4.96 [4.79, 5.24] | 197.48 [189.82, 210.77] |
| Ceará (CE) | 3455.31 [3354.56, 3642.25] | 119.95 [115.58, 127.77] | 4.57 [4.39, 4.86] | 184.10 [176.20, 198.09] |
| Rio Grande do Norte (RN) | 3069.30 [2864.62, 3196.77] | 106.17 [98.06, 111.69] | 4.02 [3.73, 4.25] | 161.66 [148.08, 171.59] |
| Paraíba (PB) | 3110.05 [3013.72, 3242.24] | 109.30 [105.11, 114.94] | 4.08 [3.92, 4.29] | 168.97 [161.37, 179.18] |
| Pernambuco (PE) | 3164.07 [2924.53, 3317.07] | 108.30 [98.89, 114.83] | 3.84 [3.53, 4.08] | 165.47 [149.63, 177.27] |
| Alagoas (AL) | 3385.44 [3244.39, 3498.46] | 118.24 [112.36, 123.14] | 4.15 [3.95, 4.33] | 184.66 [174.02, 193.93] |
| Sergipe (SE) | 3008.75 [2844.28, 3235.55] | 101.42 [94.91, 110.67] | 3.43 [3.21, 3.74] | 154.63 [143.58, 170.89] |
| Bahia (BA) | 3761.43 [3610.11, 3909.02] | 130.80 [124.48, 137.10] | 5.19 [4.95, 5.44] | 201.39 [190.20, 212.94] |
| Mato Grosso do Sul (MS) | 3417.08 [3302.47, 3520.45] | 128.55 [123.50, 133.20] | 5.53 [5.33, 5.72] | 201.14 [191.87, 209.88] |
| Mato Grosso (MT) | 3024.32 [2921.89, 3111.73] | 112.20 [107.79, 116.11] | 4.21 [4.06, 4.35] | 179.02 [170.88, 186.61] |
| Goiás (GO) | 2955.92 [2877.73, 3036.27] | 113.38 [109.81, 117.05] | 4.98 [4.83, 5.12] | 180.15 [173.34, 187.20] |
| Distrito Federal (DF) | 3875.22 [3769.43, 3963.41] | 140.83 [136.22, 144.75] | 6.41 [6.21, 6.57] | 220.09 [211.56, 227.52] |
| Minas Gerais (MG) | 3492.65 [3339.08, 3607.62] | 133.92 [127.08, 139.17] | 6.78 [6.46, 7.03] | 205.94 [193.84, 215.56] |
| Espirito Santo (ES) | 3223.68 [3108.66, 3306.15] | 123.04 [118.03, 126.85] | 5.68 [5.48, 5.84] | 192.12 [183.13, 199.30] |
| Rio de Janeiro (RJ) | 3330.37 [3211.03, 3514.55] | 117.14 [111.99, 124.91] | 4.72 [4.50, 5.02] | 179.33 [170.20, 193.19] |
| São Paulo (SP) | 3801.83 [3650.85, 3925.50] | 144.11 [137.38, 149.71] | 7.22 [6.90, 7.49] | 220.81 [208.90, 231.02] |
| Paraná (PR) | 4110.33 [3988.33, 4210.72] | 156.05 [150.58, 160.63] | 7.86 [7.60, 8.08] | 240.04 [230.18, 248.49] |
| Santa Catarina (SC) | 5551.89 [5386.66, 5690.62] | 200.76 [193.66, 206.86] | 10.11 [9.76, 10.42] | 303.85 [291.51, 314.77] |
| Rio Grande do Sul (RS) | 6173.39 [6014.42, 6337.13] | 227.51 [220.57, 234.69] | 12.38 [12.00, 12.76] | 339.37 [327.62, 351.77] |

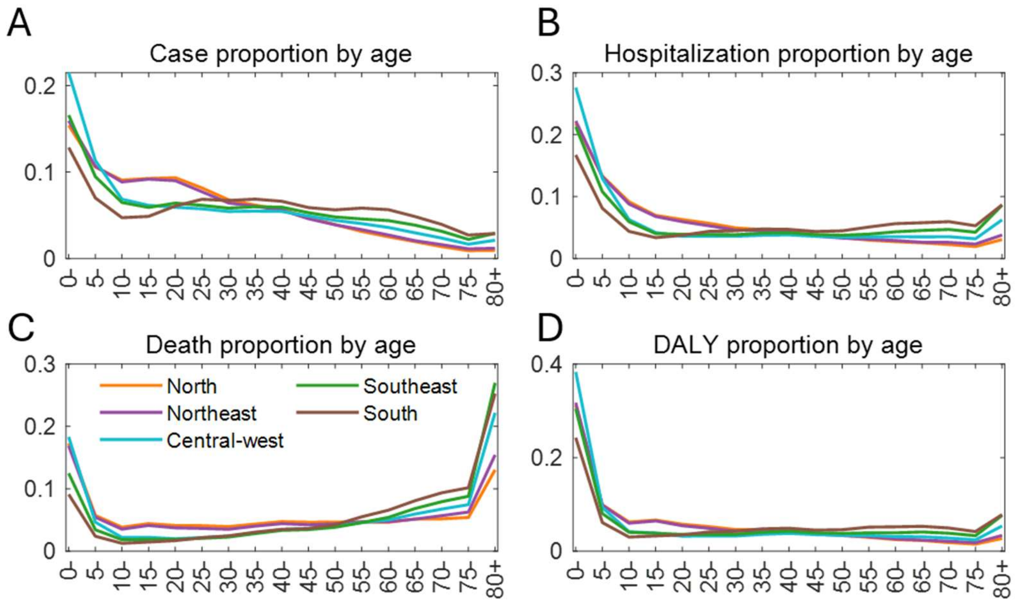


**Fig. S12**: Proportion of BoD by age group in different geographical regions. Panels A–D present the case, hospitalization, fatality, and DALY, respectively.

**S5-2. Projection of BoD under different vaccine effectiveness setting (baseline scenario)**

These supplementary results follow the baseline scenario introduced in the main text, corresponding to the projected 2024/25 dengue vaccine distribution in Brazil. We considered two sets of vaccine effectiveness (VE) estimates, as reported by Daniels and Shen [27,28]. Combining these VE assumptions with the projected future reproduction numbers, we computed the BoD averted over the next five years. **Fig. S13** displays the age-specific BoD averted per 1,000 vaccinated individuals. In terms of cases averted, the Daniels VE assumption yielded a modestly higher effect (approximately 12% greater in total) than the Shen assumption, whereas the two VE settings produced comparable averting effects for the remaining outcomes (hospitalizations, fatalities, and DALYs).

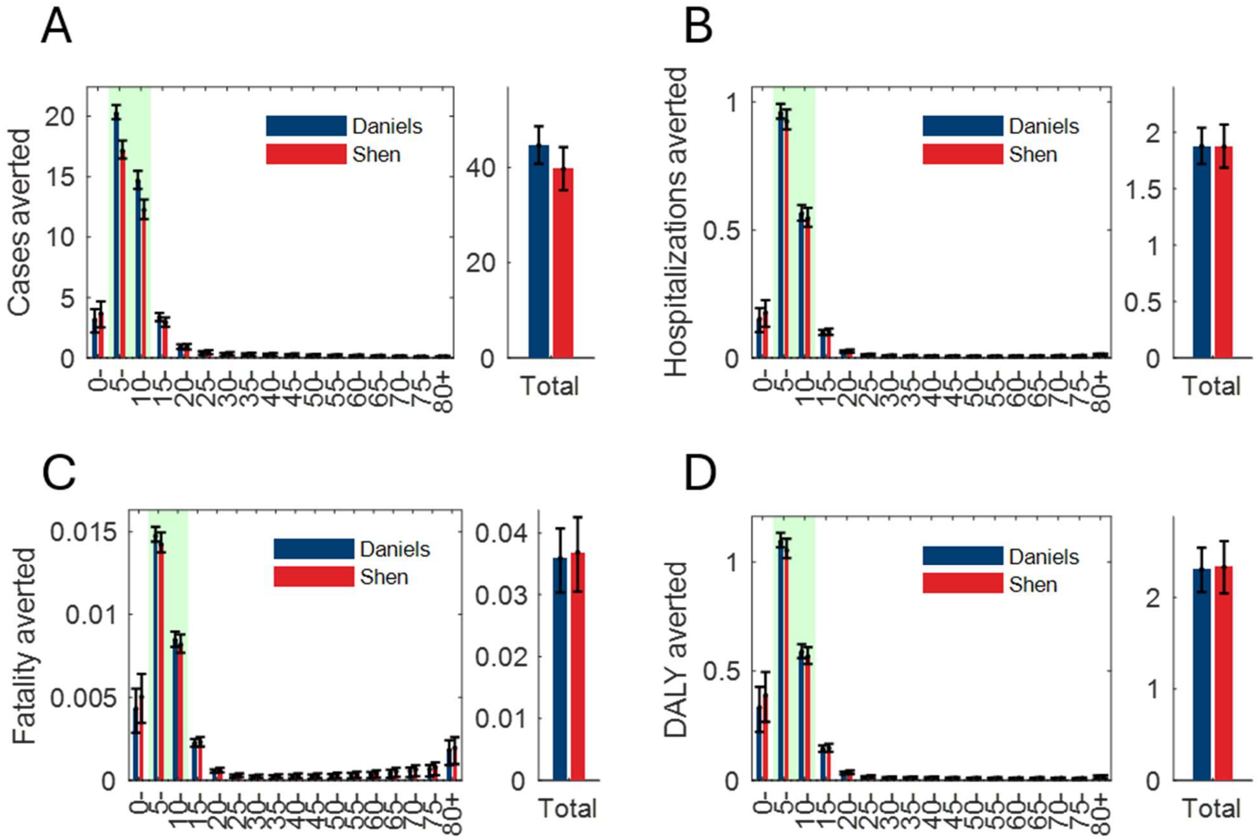


**Fig. S13**: Burden of disease (BoD) averted per 1,000 vaccinated individuals under the projected 2024/25 dengue vaccine distribution in Brazil, by age group (**A**: cases; **B**: hospitalizations; **C**: fatalities; **D**: DALYs). Blue and red bars represent simulations using vaccine effectiveness (VE) assumptions from the Daniels and Shen studies, respectively; error bars denote the interquartile range across the R0 = Q25–Q75 scenarios. Shaded green regions indicate the age groups (5–14 years) prioritized under the current allocation. Side panels show the corresponding totals summed across all age groups.

**S5-3. Optimal state-level vaccine allocation targeting children (or elderly)**

We first describe the spatial patterns of the optimized vaccine allocation strategies targeting either children (5–14 years) or the elderly (≥60 years), and then evaluate their expected public health impact. The vaccine distribution listed in **Table S10** reflects allocation criteria used by the Brazilian Ministry of Health, one of which incorporates recent incidence [29–36]. Figs. S14 and S15 show the optimized allocation strategies obtained under two alternative VE assumptions, with panels corresponding to different projected reproduction numbers (Q25, Q50, and Q75 scenarios) and target groups, presented as scatter plots against past incidence.

For the children-targeting strategy, cumulative incidence over the past ten years was negatively correlated with optimized allocation across all settings. Under the Shen VE assumption, correlations were r = −0.66 (p = 0.00), −0.66 (p = 0.00), and −0.84 (p = 0.00) for the Q25, Q50, and Q75 scenarios, respectively; under the Daniels VE assumption, the corresponding correlations were r = −0.46 (p = 0.02), −0.64 (p = 0.00), and −0.63 (p = 0.00). For the elderly-targeting strategy, correlations were positive across all settings but were not statistically significant: under the Shen VE assumption, r = 0.15 (p = 0.46), 0.14 (p = 0.49), and 0.26 (p = 0.18) for the Q25, Q50, and Q75 scenarios, respectively; under the Daniels VE assumption, the corresponding correlations were r = 0.14 (p = 0.49), 0.28 (p = 0.15), and 0.13 (p = 0.52).

We further compared the optimized allocations with cumulative incidence over the past ten years, alongside the actual Brazilian Ministry of Health distribution. Red markers represent the children-targeting optimized allocation by state, blue markers represent the elderly-targeting optimized allocation, and green markers denote the Ministry of Health distribution. The correlation coefficients were r = −0.74 (p = 0.00) for the children-targeting allocation, r = 0.18 (p = 0.36) for the elderly-targeting allocation, and r = 0.47 (p = 0.01) for the Brazilian distribution. Below the scatter plots, choropleth maps (**Figs. S16** and **S17**) illustrate the allocation weights across states, with darker shades indicating higher weight. Notably, while the scatter plots alone do not reveal a strong pattern for the elderly-targeting scenario, the maps highlight a pronounced concentration of vaccine allocation in the southern states under this strategy.

**Fig. S18** compares the optimized allocation vectors derived under the Shen and Daniels VE assumptions directly, with vertical dotted lines connecting the two VE-specific allocations for each state at matching R0 quantiles. The Pearson correlation between the Daniels- and Shen-derived allocation vectors was high for the elderly-targeting strategy at Q25 and Q75 (r = 0.97–0.99) and for the children-targeting strategy at Q75 (r = 0.81), but lower in the remaining scenarios (r = 0.21–0.46). These results are based on numerical optimization and may therefore differ slightly in magnitude across runs, but the overall allocation trends were consistent.

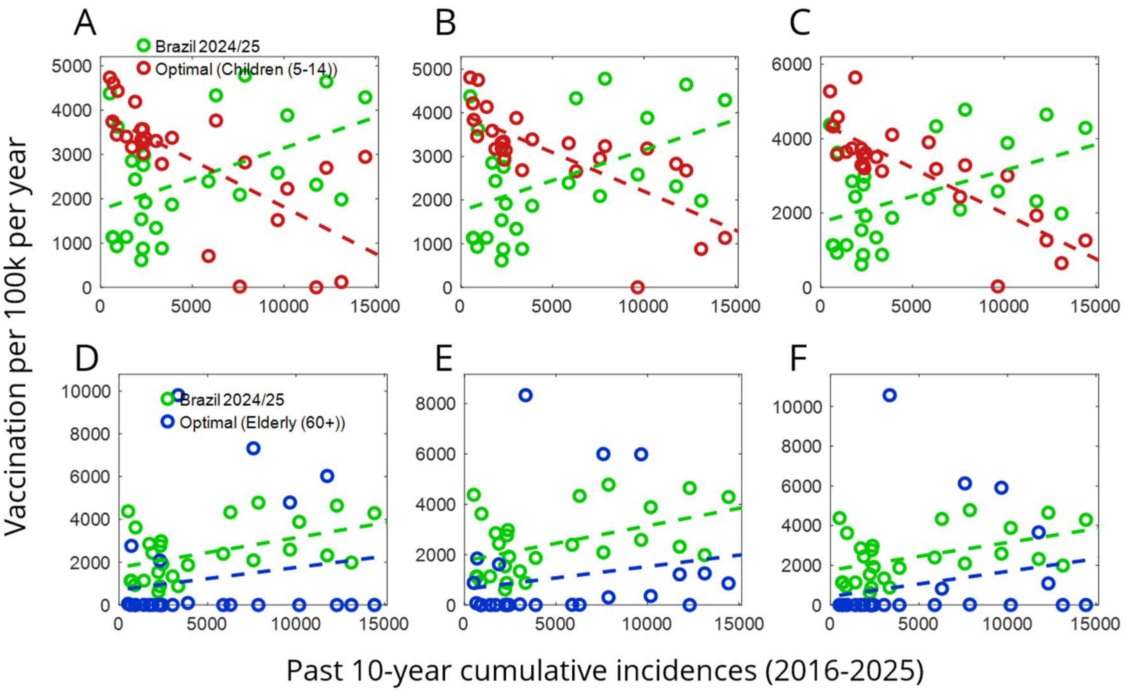


**Fig. S14**: Optimized vaccine allocation results under Shen's vaccine effectiveness assumptions, computed using the 25th, 50th, and 75th percentile (Q25, Q50, Q75) reproductive number scenarios, for children-targeting and elderly-targeting strategies. Panels A, B, and C show the children-targeting allocation at Q25, Q50, and Q75, respectively; panels D, E, and F show the elderly-targeting allocation at Q25, Q50, and Q75, respectively

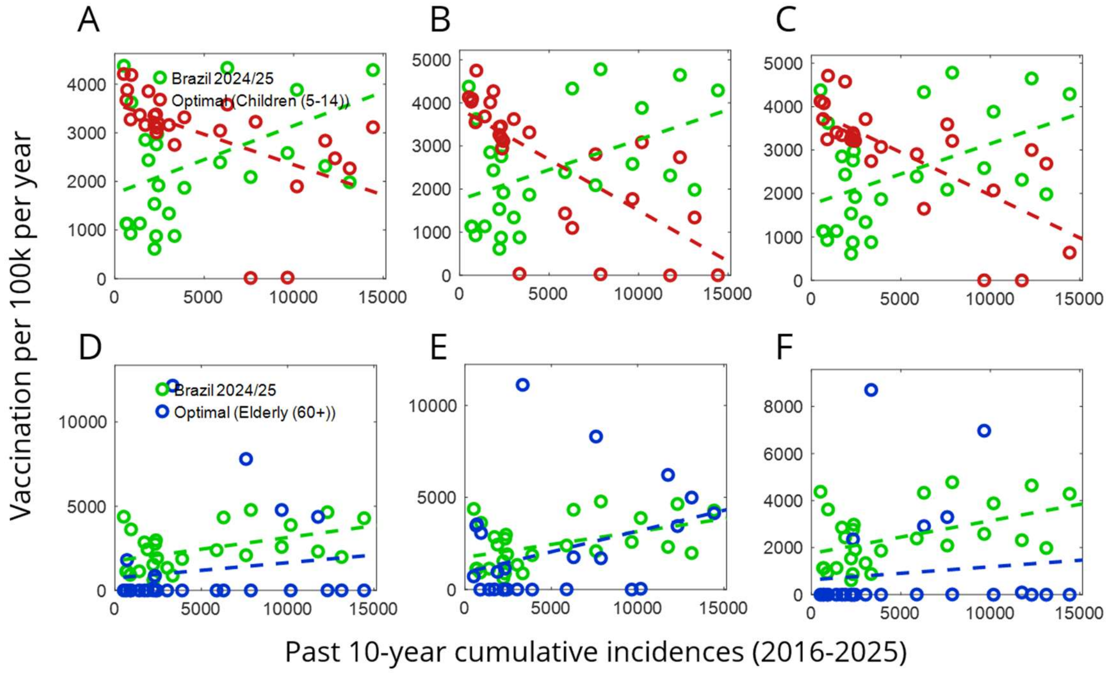


**Fig. S15**: Optimized vaccine allocation results under Daniels' vaccine effectiveness assumptions, computed using the 25th, 50th, and 75th percentile (Q25, Q50, Q75) reproductive number scenarios, for children-targeting and elderly-targeting strategies. Panels A, B, and C show the children-targeting allocation at Q25, Q50, and Q75, respectively; panels D, E, and F show the elderly-targeting allocation at Q25, Q50, and Q75, respectively

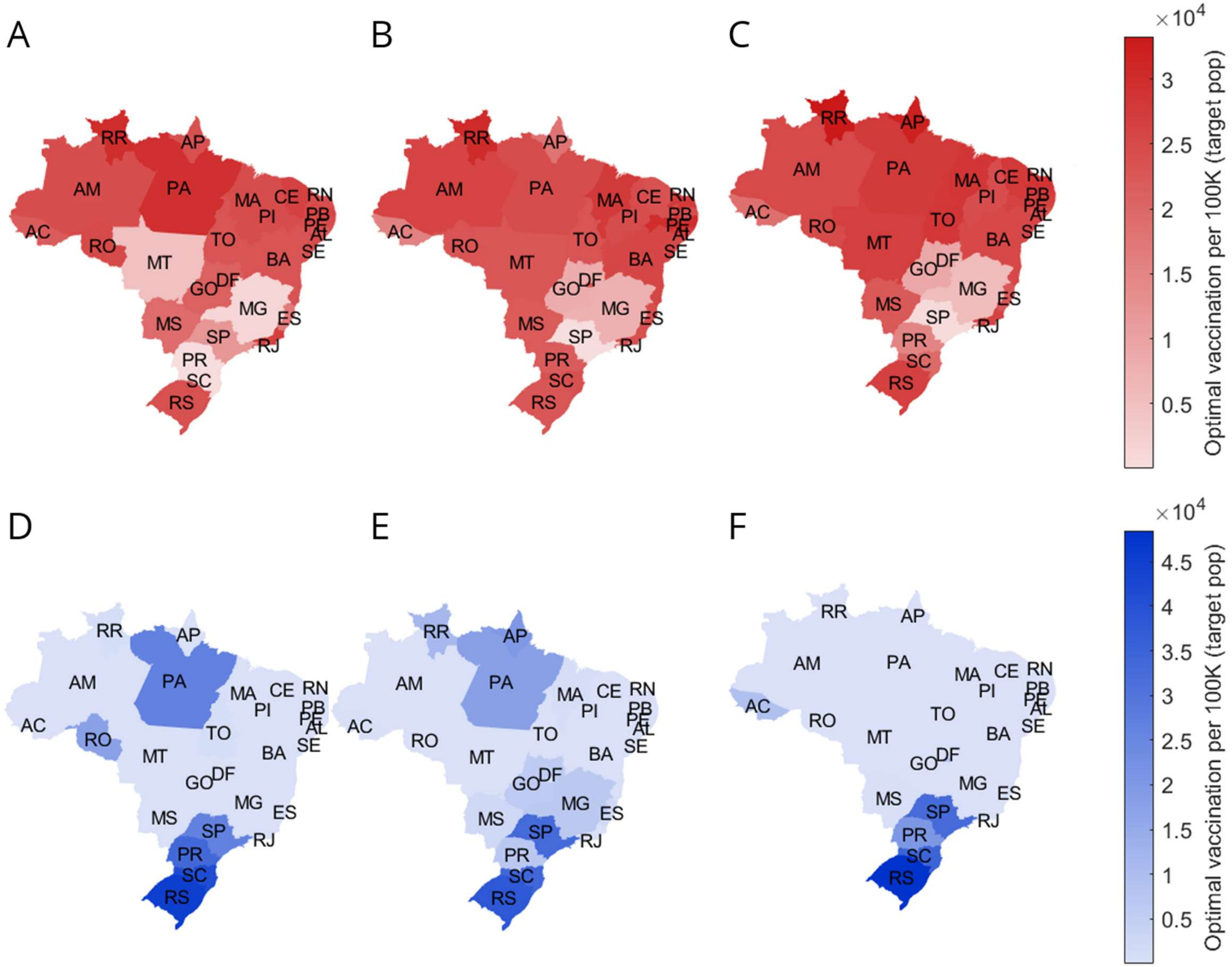


**Fig. S16**: Choropleth maps of optimized vaccine allocation under Shen's vaccine effectiveness assumptions, computed using the 25th, 50th, and 75th percentile (Q25, Q50, Q75) reproductive number scenarios, for children-targeting and elderly-targeting strategies. Panels A, B, and C show the children-targeting allocation at Q25, Q50, and Q75, respectively; panels D, E, and F show the elderly-targeting allocation at Q25, Q50, and Q75, respectively

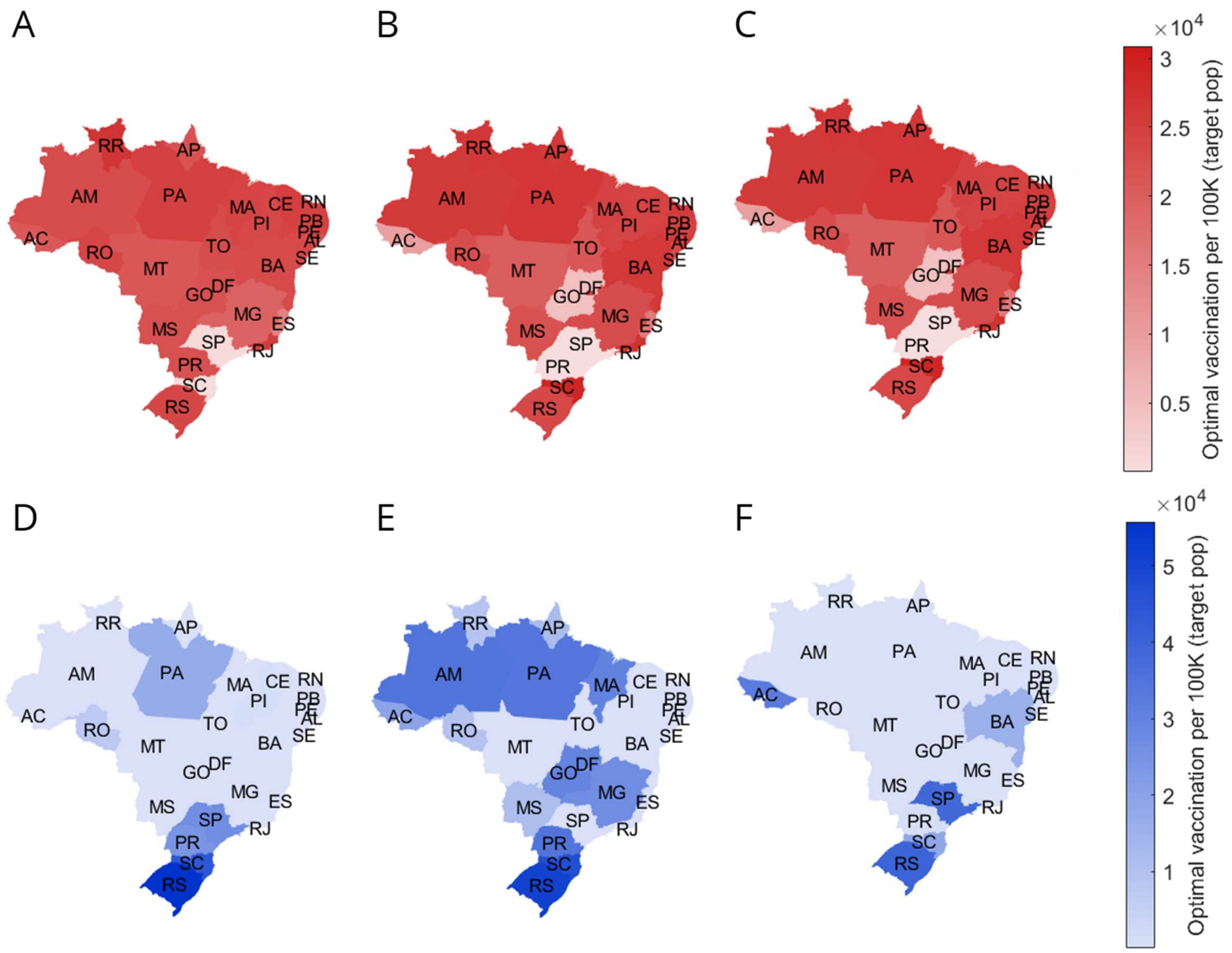


**Fig. S17**: Choropleth maps of optimized vaccine allocation under Daniels' vaccine effectiveness assumptions, computed using the 25th, 50th, and 75th percentile (Q25, Q50, Q75) reproductive number scenarios, for children-targeting and elderly-targeting strategies. Panels A, B, and C show the children-targeting allocation at Q25, Q50, and Q75, respectively; panels D, E, and F show the elderly-targeting allocation at Q25, Q50, and Q75, respectively

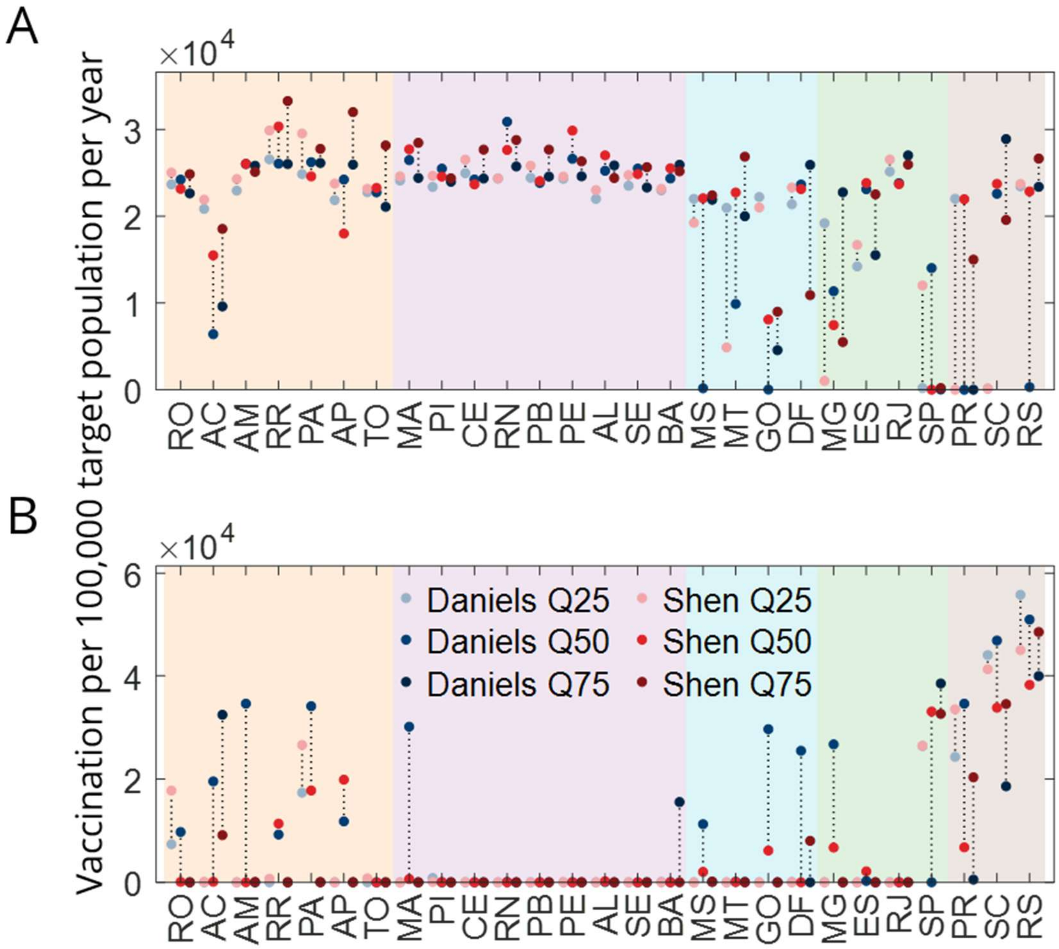


**Fig. S18**: Optimal vaccine allocation under Daniels- and Shen-derived vaccine efficacy (VE) assumptions, by state, across three basic reproduction number (R0) scenarios (Q25, Q50, Q75). Background shading indicates geographic region (orange: North; purple: Northeast; cyan: Central-west; green: Southeast; brown: South). **(A)** Target population size per state (children aged 5–14 or elderly aged 60+, depending on target group), shown for reference. **(B)** Optimal annual vaccine allocation per 100,000 target-age population, by state. Each point represents one R0 scenario–VE assumption combination; blue tones denote allocations optimized under Daniels VE estimates, red tones denote allocations optimized under Shen VE estimates, with color intensity increasing from Q25 (lightest) to Q75 (darkest). Dotted vertical lines connect the Daniels and Shen allocations at the same R0 quantile for each state, illustrating the degree of divergence between the two VE assumptions.
